# Distinct trajectories of childhood atopic dermatitis are associated with differences in long-term inflammatory and cardiometabolic disease risks

**DOI:** 10.1101/2025.08.18.25333898

**Authors:** Florian Thaçi, Philip Curman, Katja Bieber, Henning Olbrich, Diamant Thaçi, Ralf J. Ludwig

**Affiliations:** Lübeck Institute of Experimental Dermatology, University of Lübeck, Lübeck, Germany; Dermato-Venereology Clinic, Karolinska University Hospital, Stockholm, Sweden; Division of Dermatology and Venereology, Department of Medicine (Solna), Karolinska Institutet, Stockholm, Sweden; Department of Medical Epidemiology and Biostatistics, Karolinska Institutet, Stockholm, Sweden; Department of Dermatology, University-Hospital Schleswig-Holstein (UKSH), Lübeck, Germany; Institute and Comprehensive Center for Inflammation Medicine, University of Lübeck, Lübeck, Germany

**Keywords:** <u>Keywords</u>: Atopic dermatitis, childhood, persistence, type 2 inflammation, autoimmune disease, trajectory, real-world-data, TriNetX

## Abstract

**Importance:** Atopic dermatitis (AD) is a common childhood inflammatory skin disease which occurs frequently in the early childhood and is later linked to type 2 diseases but also other systemic comorbidities. The role of different AD-disease trajectories on long-term health outcomes remains unclear, as prior studies have rarely conducted direct comparisons between distinct AD trajectories.

**Objective:** To assess how different childhood AD trajectories - persistent, transient - or none, are associated with long-term risks of other type 2 inflammatory diseases (T2IDs), autoimmune diseases, and cardiometabolic disorders by performing all pairwise comparisons to identify trajectory-specific risk patterns.

**Design:** Retrospective cohort study using US TriNetX Analyses included propensity-score matching, sensitivity analyses, and subgroup (sex and ancestry) stratification.

**Setting:** Multicenter, population-based network of electronic health records from diverse US healthcare organizations.

**Participants:** Children with AD onset before age of two were grouped by disease trajectory (persistent or transient) and compared with matched non-AD controls.

**Exposures:** AD trajectory phenotype (persistent vs transient) or no AD.

**Main outcomes and measures:** Hazard ratios (HRs) for T2IDs, autoimmune diseases, cardiovascular risk factors, venous thromboembolism (VTE), and major adverse cardiovascular events (MACE).

**Results:** Persistent AD was associated with increased risk of T2IDs (HR 2.11, 95%-CI 2.01– 2.21), autoimmune diseases (HR 1.68, 1.60–1.76), and cardiovascular risk factors (HR 1.38, 1.22–1.56), but not VTE or MACE. Compared with transient, persistent AD conferred higher risk across most outcomes. EoE risk was especially elevated in females and children with Black or African American ancestry; other sex- or racial disparities were limited.

**Conclusions and relevance:** Persistent childhood AD is associated with a substantial long-term inflammatory and cardiometabolic burden, whereas the risk conferred by transient AD appears minimal. These findings underscore the prognostic importance of disease trajectory and support timely disease activity-adapted interventions in children.

## Introduction

Atopic dermatitis (AD) is among the most prevalent chronic inflammatory skin conditions in childhood, affecting up to 20% of children worldwide. It often emerges within the first two years of life and imposes a considerable burden on affected children and their families, both in terms of symptom severity and healthcare utilization (1–4). Despite its frequently early onset, the disease does not follow a uniform clinical course. Some infants and children experience spontaneous resolution by early childhood, while others suffer from persistent or relapsing symptoms that extend into adolescence or adulthood. This heterogeneity in disease trajectories could be a critical factor in determining long-term health outcomes (5–8).

Yet, whilst studies addressing disease trajectories in AD have well defined the different AD subgroups, i.e., defined by onset and resolution, (5,9), few studies have investigated the long-term impact of these different AD disease trajectories (6). In the latter study, the odds of developing other type 2 inflammatory diseases (T2IDs), such as asthma, food allergy, and allergic rhinitis, were assessed for different AD disease trajectories, using non-AD individuals as the reference group. However, due to relatively small sample sizes, direct comparisons between the individual AD trajectories were not feasible. In addition, a recent cohort study that pooled longitudinal data from 12 observational US birth cohorts spanning several decades found that, compared with minimal or no AD, early-onset AD phenotypes were associated with increased risk of food allergy, while later-onset AD phenotypes were linked to allergic rhinitis. Again, these differences were based on comparisons with a shared reference group rather than direct statistical comparisons between AD disease trajectories (10).

In addition to T2IDs, there is accumulating evidence that AD is associated with an increased risk of the subsequent diagnosis of autoimmune diseases (11,12). Notably, the risk for the occurrence of of autoimmune diseases corelated with disease severity. As in studies assessing the risk of subsequent T2IDs, this statement is based on comparing the magnitude of risk between different AD severity levels relative to non-AD controls, rather than on direct comparisons between AD severity groups (11). Relating the risks for cardiovascular diseases, i.e., MACE, VTE, as well as cardiovascular risk factors the current evidence does not support a clinically relevant increase (13). One recent longitudinal cohort study suggests that early-onset and severe AD may be associated with modest increases in cardiovascular risk factors by young adulthood in males but not females (14).

Like disease trajectories, sex- and racial disparities in AD are well documented (15,16). For example, male children are more likely than females to develop early-onset and persistent AD (10). In the US, Black or African American and Hispanic or Latino children have a higher prevalence of AD, along with greater disease severity and increased persistence compared to White children (17). While a few studies have begun to explore sex- and ancestry-related differences in AD endotypes, the impact of these factors on long-term disease trajectories remains unknown (17).

To address these knowledge gaps, namely, the limited understanding of differential long-term comorbidity risks across AD trajectories and the potential influence of sex and ancestry, we conducted retrospective cohort analyses using a large-scale electronic health record (EHR) database.

## Methods

### Ethics statement

The data analyzed is a secondary analysis of existing data, does not involve intervention or interaction with human subjects, and is de-identified per the de-identification standard defined in Section §164.514(a) of the HIPAA Privacy Rule. TriNetX has a security and governance model that provides a federated platform of real-world-data which is Health Insurance Portability and Accountability Act (HIPAA), General Data Protection Regulation (GDPR), and Lei Geral de Proteção de Dados (LGPD)-compliant. Thus, our study does not require Institutional Review Board approval.

### Study design and database

A propensity-score matched (PSM) retrospective cohort study was conducted using the US Collaborative Network from the federated electronic EHR database TriNetX, based on previously published studies (18–20). This network was selected due to the large number of documented EHRs and the high degree of covariate documentation (21). We aimed to contrast the risks for a subsequent diagnosis of (**i**) additional T2IDs, (**ii**) autoimmune diseases, (**iii**) cardiovascular and metabolic disease in children with (**i**) persistent childhood AD, (**ii**) transient childhood AD, and (**iii**) children with no AD (**Figure 1**). The study was performed according to the STROBE guidelines (22).

**Figure 1.**
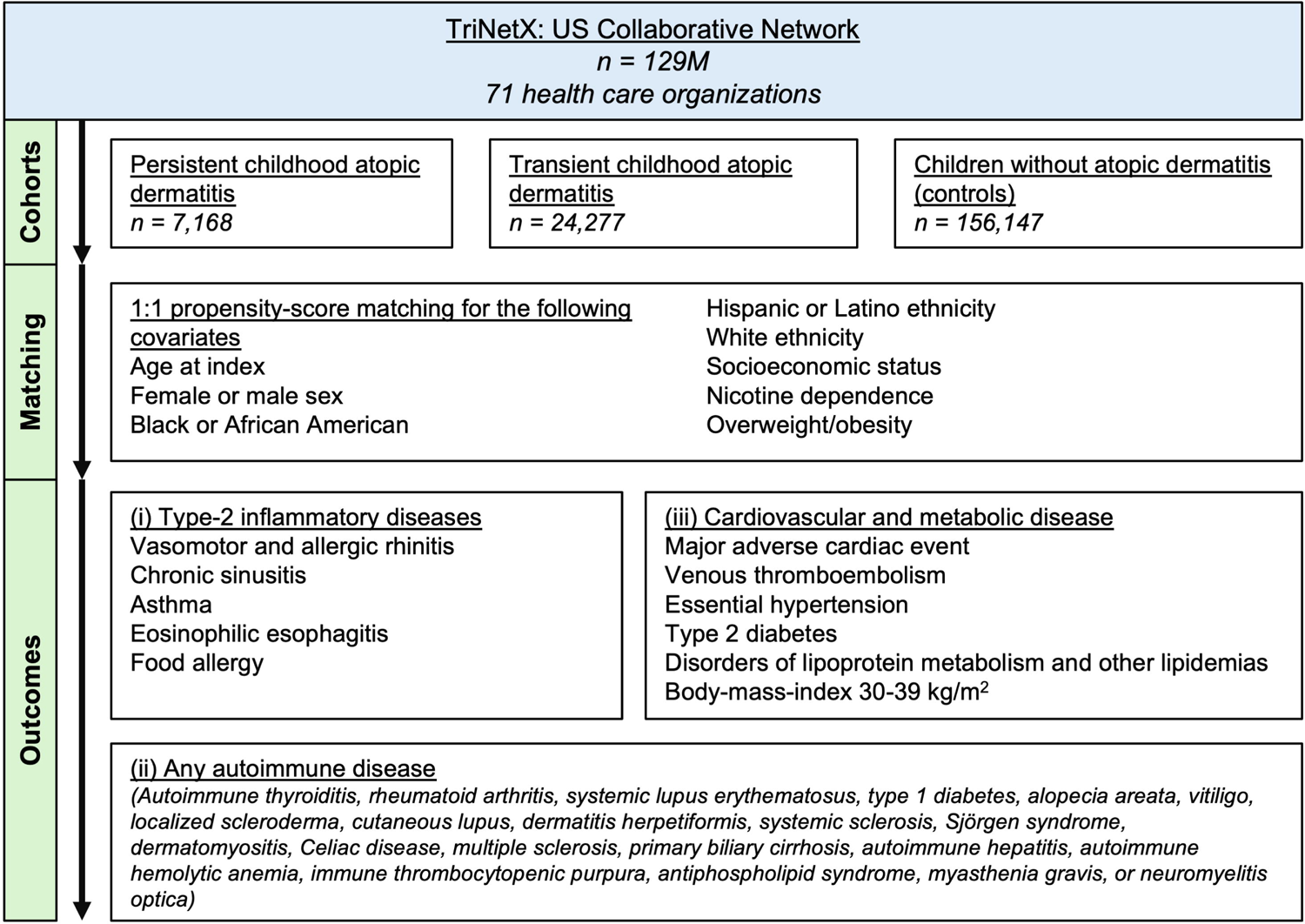
Study flow-chart.

### Study population

The study was conducted from April to May 2025. At the time of analysis, the database provided access to EHRs from close to 130 million patients across 71 healthcare organizations (HCOs) in the United States. We retrieved three distinct cohorts: (**i**) Persistent childhood AD, defined by documentation of the International Classification of Diseases, 10^th^ edition, Clinical Modification (ICD-10-CM) code L20 in patients aged 2 years or younger, excluding the following T2IDs: Vasomotor and allergic rhinitis (J30), chronic sinusitis (J32), asthma (J45), eosinophilic esophagitis (EoE, K20.0), anaphylactic reaction due to food (T78.0), or food allergy status (Z91.01). Persistence was defined as a second documentation of ICD-10-CM code L20 at or after the age of 12 years. (**ii**) Transient childhood AD was defined using the same criteria as persistent AD with transient AD defined as a healthcare encounter at age 12 years or older and no subsequent documentation of L20 after the age of 3 years. (**iii**) The non-AD cohort was defined by an encounter for general examination without complaint, suspected or reported diagnosis (Z00) in patients aged 2 years or younger, excluding all aforementioned T2IDs (including AD). In addition, a second instance of Z00 had to be recorded at age 12 years or older, again excluding all EHRs with a documentation of L20. Note that the index event was defined as the initial AD diagnosis (for both AD cohorts) or the healthcare encounter for the non-AD control group.

### Outcomes

Outcomes were categorized into (**i**) other T2IDs, specifically: vasomotor and allergic rhinitis (ICD-10-CM:J20, VMR/AR), chronic sinusitis (J32, CS), asthma (J45), eosinophilic esophagitis (K20.0, EoE), food allergy (Z91.01 or T78.9), or any of these T2IDs, (**ii**) any autoimmune disease, including a diagnosis of autoimmune thyroids (E06.3), rheumatoid arthritis (RA) with rheumatoid factor (M05)m other RA (06), systemic lupus erythematosus (M32, SLE), type 1 diabetes (E10), alopecia reata (L63) vitiligo (L80), localized scleroderma (L94.0), cutaneous lupus (L93), dermatitis herpetiformis (L13.9), systemic sclerosis (M34, SSc), Sjögren syndrome (M35.0), Dermatomyositis (M33), Celiac disease (K90.0), multiple sclerosis (G35), primary biliary cirrhosis (K74.3), autoimmune hepatitis 8K75.4), autoimmune hemolytic anemia (D59.10), immune thrombocytopenic purpura 8D69.3), antiphospholipid syndrome (IDC-10-CM:D68.61), myasthenia gravis (G70), or neuromyelitis optica (G36.0), (**iii**) major adverse cardiac event (MACE: I21, I22, I46, I50, I61-63, R57.0), venous thromboembolism (VTE: I26, I82.4, I80.0), and cardiovascular risk factors: Essential hypertension (I10), type 2 diabetes (E11), disorders of lipoprotein metabolism and other lipidemias (E78), or body-mass-index (BMI) 30-39 kg/m^2^ (Z68.3).

### Covariates

To minimize bias from potentially confounding variables, PSM was performed by establishing a covariate matrix that included relevant concomitant diagnoses, laboratory findings, and demographic information. Matching was performed 1:1 using propensity scores. Baseline characteristics were reported before and after matching. The following covariates were included in PSM for all analyses (binary unless otherwise stated): Age at index (continuous), female or male sex, self-reported ancestry (Black or African American, Hispanic or Latino, and White, ICD-10-CM codes Z55-Z65 (representing individuals with potential health hazards related to socioeconomic and psychosocial circumstances, used as a proxy for socioeconomic status), nicotine dependence (F17), and overweight or obesity (E66). In addition to the covariates listed above, the following variables were included for subgroup-specific matching: (**i**) for other T2IDs) the following covariates were considered: Family history of asthma and other chronic lower respiratory diseases (Z82.5), and family history of diseases of the skin and subcutaneous tissue (Z84.0); (**ii**) for autoimmune diseases, the following covariates were considered: Family history of other endocrine, nutritional and metabolic diseases (Z83.4), and family history of arthritis and other diseases of the musculoskeletal system and connective tissue (Z82.6); and (**iii**) for cardiovascular and metabolic diseases, the following covariates were considered: Family history of ischemic heart disease and other diseases of the circulatory system (Z82.4), diabetes mellitus (E08-E13), and disorders of lipoprotein metabolism and other lipidemias (E78).

### Primary, sensitivity and subgroup analyses

In the **primary analysis**, any outcomes occurring from one day after the index event were considered. In sensitivity analysis **S1**, to exclude already present but not diagnosed or coded outcomes at the index event, any outcomes occurring three months after the index event were considered. In **S2**, to account for changes in clinical practice, only EHRs documented after January 1^st^, 2004, were considered. For subgroup analyses, EHRs were stratified by sex (female, male) or by self-reported ancestry (Black or African American, Hispanic or Latino, or White). For all subgroup analyses EHRs documented after January 1^st^, 2004, were considered.

### Statistical analysis

A propensity score for each patient was generated by logistic regression analysis (with exposure as the dependent variable) using the Python package scikit-learn. Matching was performed 1:1 using the greedy nearest neighbor approach with a cut-off distance of 0.1 pooled standard deviations of the logit of the propensity score. Baseline characteristics were re-evaluated and reported after matching, differences were compared by t-test for continuous and z-test for binary or categorical variables. Relative risks and risk difference (RD) were calculated. Survival analysis was performed using the Kaplan–Meier method in Survival package v3.2–3 in R (R Foundation for Statistical Computing, Vienna, Austria) and validated by comparison with the outputs of SAS version 9.4 (SAS, Cary, NC). The proportionality assumption was tested by the coxph function in R’s Survival package. Kaplan-Meier-curves were compared using the Log-rank test. A univariate Cox proportional hazards regression was used to express hazard ratios (HR)s with 95%-confidence intervals (CI)s. Outcomes prior to index were excluded. To counter the bias introduced by multiple testing, a Bonferroni correction was applied, adjusting the significance threshold based on the number of outcomes tested within each disease category: type-2 inflammatory diseases (T2IDs, n=6; adjusted α=0.0083), autoimmune diseases (n=1; α=0.05), and cardiovascular outcomes (n=3; adjusted α=0.0167).

## Results

### Baseline characteristics

Depending on the specific comparison and outcome examined, between 4,867 and 18,099 EHRs were included in the analysis. **Table 1** lists full details on the comparisons between (**i**) persistent versus transient childhood AD, (**ii**) persistent versus no AD, and (**iii**) transient versus no AD for the primary analyses, relating to the data retrieved to compare risks of subsequent T2IDs among the groups. Baseline characteristics for all other analysis including sensitivity and subgroup analyses, as well as analyses with PSM tailored to analyze the risks of autoimmune and cardiovascular diseases are detailed in **Supplement Tables 1-3**. All cohorts were followed for a minimum of 12 years.

**Table 1.**
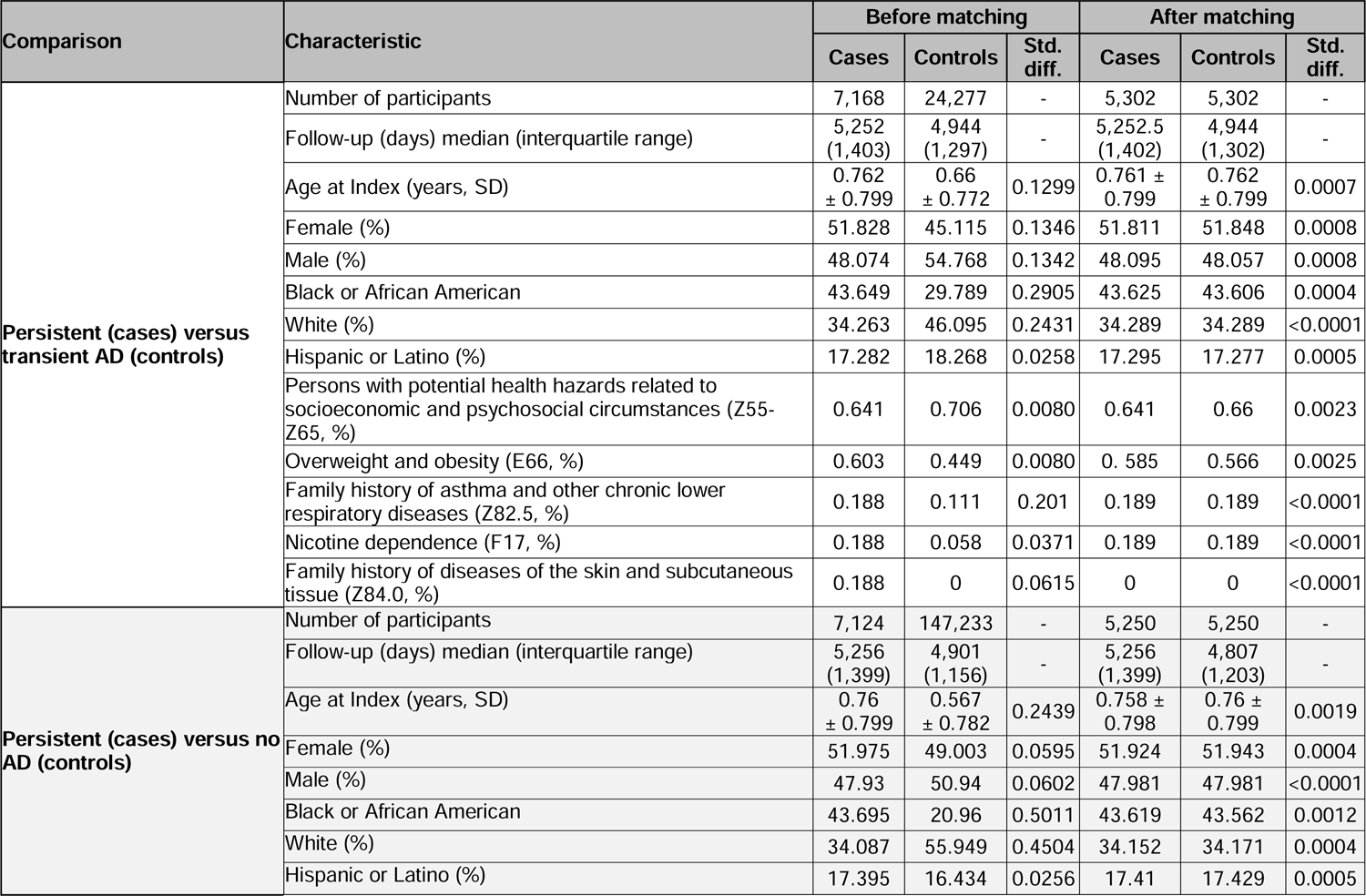
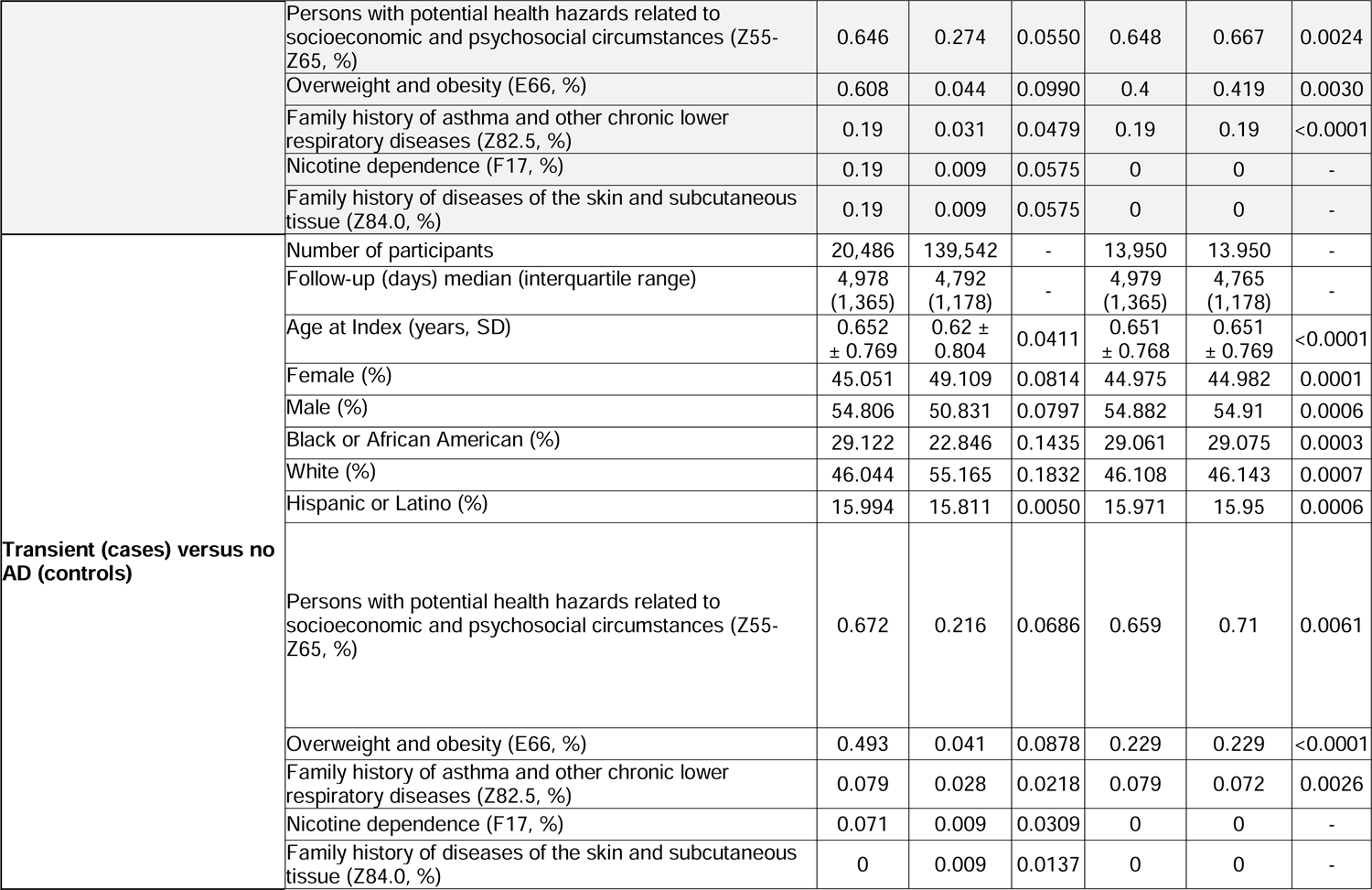
Baseline characteristics before and after propensity-score matching for the primary and S1 analyses relating to the risk of type 2 inflammatory diseases (T2IDs) in patients with persistent, transient of no childhood atopic dermatitis (AD).

### Persistent AD substantially increases comorbidity risks, while transient AD only marginally impacts on AD comorbidity patterns

Contrasting persistent and no childhood AD trajectories revealed that persistent AD is associated with a markedly increased comorbidity risk. Specifically, in the primary analysis, 80.46% of individuals with persistent AD developed at least one T2ID, compared to 50.91% of those without childhood AD, translating into a hazard ratio (HR) of 2.11 (95% confidence interval [CI], 2.01–2.21; p<0.0001). The highest individual risk increases were observed for food allergy (HR 9.18, 7.95–10.60) and EoE (HR 8.72, 4.68–16.25). Likewise, the risks for asthma or AR/VMR were significantly elevated, while no increased risk was observed for chronic sinusitis (**Figure 2**, **Supplement Tables 4–6**).

**Figure 2.**
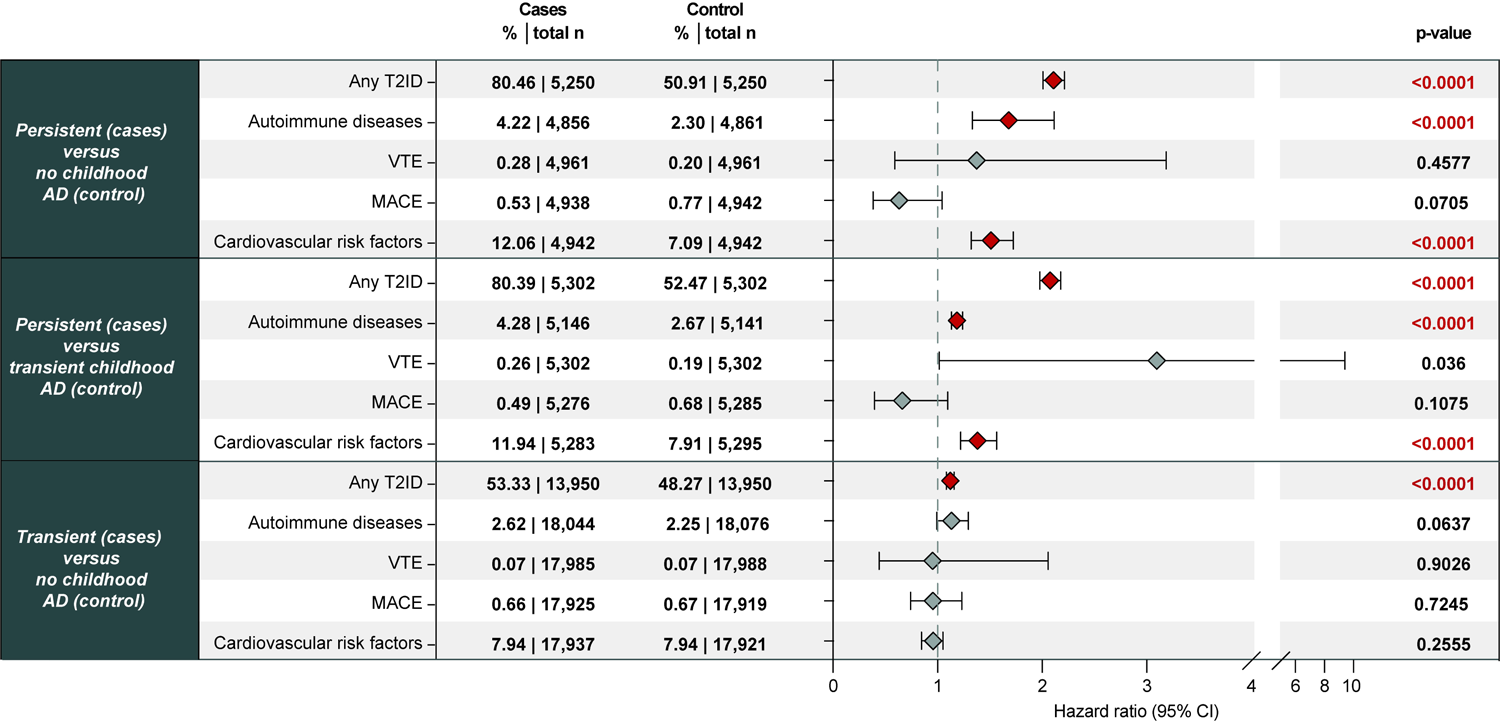
Risks of comorbid outcomes in children with persisting, transient, or no atopic dermatitis (AD). The figure presents hazard ratios (HRs) and 95% confidence intervals (CIs) for the indicated outcomes in children with persisting, transient AD, or without AD. Outcomes are grouped into three categories: Type-2 inflammatory diseases (T2IDs), autoimmune diseases, and cardiovascular outcomes. To counter the bias introduced by multiple testing, a Bonferroni correction was applied, adjusting the significance threshold based on the number of outcomes tested within each category: T2IDs (n=6; adjusted α=0.0083), autoimmune diseases (n=1; α=0.05), and cardiovascular outcomes (n=3; adjusted α=0.0167).

Similar risk patterns emerged when comparing T2ID trajectories between patients with persistent versus transient childhood AD. The overall risk of receiving a subsequent T2ID diagnosis was 80.39% in individuals with persistent AD, compared to 52.47% in those with transient AD (HR 2.08, 1.98-2.18). As in the prior analysis, the highest individual risks were again documented for food allergy (HR 5.45, 4.86-6.11) and EoE (HR 4.23, 2.69-6.65). Furthermore, the risks for asthma, AR/VMR, and chronic sinusitis were also significantly higher in patients with persistent AD compared to those with transient disease (**Figure 2**, **Supplement Tables 4–6**).

Notably, when comparing T2ID risk between patients with transient AD and those without AD, a statistically significant, but overall marginal, increase in risk was observed for individuals diagnosed with transient AD. These risks to be diagnosed a T2ID following the respective index events were consistent with the patterns observed in the previous comparisons: The overall risk of developing any T2ID was 53.33% in the transient AD group compared to 48.27% in the non-AD group, (HR 1.12, 1.09-1.16). Among individual T2IDs, the highest relative risk increases, though descriptively lower than those seen in persistent AD, were observed for food allergy and EoE. Asthma and AR/VMR also showed a marginal, but again, significant increase in risk associated with transient childhood AD, while no increased risk was observed for chronic sinusitis (**Figure 2**, **Supplement Tables 4–6**).

Taken together, these analyses indicate that the long-term risk of developing T2IDs is primarily driven by persistent childhood AD. While persistent AD was associated with a markedly elevated risk compared to both transient and no AD, the difference in T2ID risk between transient and no AD was marginal. The near-identical hazard ratios observed when comparing persistent AD to either transient or no AD further support the assumption that transient AD contributes minimally to long-term, overall T2ID burden.

In addition to T2IDs, AD has also been proposed as a potential risk factor for autoimmune diseases (23). Thus, we next analyzed the risk of developing autoimmune diseases across different childhood AD trajectories. In the primary analysis, individuals with persistent childhood AD had a markedly increased risk compared to those without AD (HR 1.68, 1.60–1.76). Similarly, persistent AD conferred a significantly elevated risk when compared to transient AD (HR 1.62, 1.55-1.70). In contrast, transient AD was associated with only a modest increase in autoimmune disease risk compared to no AD (HR 1.23 1.19–1.27). These results were consistent across all three sensitivity analyses. These findings indicate that persistent childhood AD is a key driver of long-term autoimmune comorbidity, while the contribution of transient childhood AD appears limited (**Figure 2**, **Supplement Tables 4–6**).

Furthermore, AD has also been implicated as a risk factor for cardiovascular disease and related risk factors (24). Following correction for multiple testing, no differences in MACE or VTE risks were noted among any of the three comparisons across all analyses. Regarding the cardiovascular risk factors, slightly, but significantly increased were noted when comparing persistent childhood AD with no AD (HR 1.38, 1.22-1.56) or with no AD (HR 1.51, 1.32.1.73). Notably, no difference regarding to subsequent cardiovascular risk factor documentation was noted between transient and no AD. Again, all results remained robust across all sensitivity analyses, with comparable effect sizes (**Figure 2**, **Supplement Tables 4–6**).

### Sex-stratified analyses reveal greater EoE risks in females with AD

The overall risk of developing any T2ID in the subgroup analyses stratified for female of male sex mirrored the findings of the primary and S1-3 analyses (**Supplement Tables 4-6**). At the individual T2ID level, EoE risk was markedly higher in females, particularly when comparing persistent or transient childhood AD to no AD. For example, in the sex-stratified analysis, persistent childhood AD was associated with a substantially increased risk of EoE in females (HR 37.70, 5.18-274.21), considerably higher than the corresponding risk in males (HR 6.01, 3.18-11.35, **Figure 3**). The risk of autoimmune diseases was consistent with the findings from the non-stratified analyses across both sexes. For VTE, MACE, and cardiovascular risk factors, overall patterns were similar to the main analyses, although some sex-specific differences were observed. These were, however, not consistent across outcomes or AD trajectories (**Supplement Tables 4-6**).

**Figure 3.**
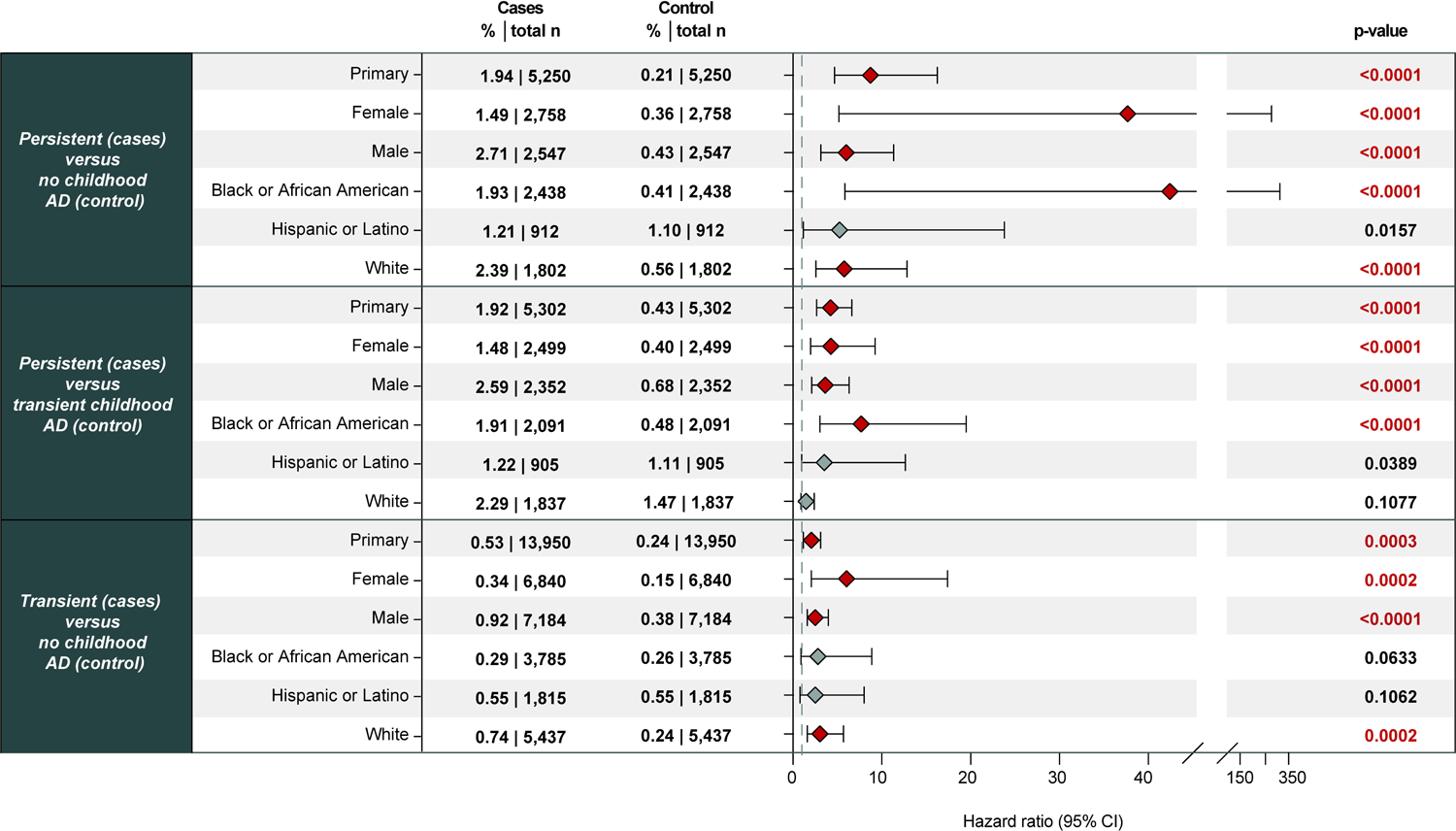
Risk of eosinophilic esophagitis (EoE) in children with persisting, transient, or no atopic dermatitis (AD), stratified by sex and ancestry. The figure presents hazard ratios (HRs) and 95% confidence intervals (CIs) for eosinophilic esophagitis (EoE) in children with persisting or transient AD compared to children without AD. Analyses were stratified by sex (male, female) and self-reported ancestry (Black or African American, Hispanic or Latino, White). Across strata, children with persisting AD consistently exhibited the highest risk of developing EoE. Notably, the risk of EoE was markedly elevated in females and Black or African American children with persisting AD, suggesting potential sex- and race-related disparities in this comorbidity pattern. A Bonferroni correction was applied to adjust for multiple testing (n=1 outcome; α=0.05). HRs should be interpreted with caution where proportional hazards assumptions may not have been met.

### Ancestry-specific differences for EoE risk

The overall risk of developing any T2ID in ancestry-stratified analyses was generally consistent with the findings of the primary and sensitivity analyses. Except for the comparison between transient childhood AD and no AD, EoE risk showed a striking racial disparity. For example, among children with persistent AD, the risk of EoE was approximately fivefold higher in those with Hispanic or Latino or White ancestry compared to those without AD. In contrast, the risk was more than 42-fold higher in children with Black or African American ancestry (**Figure 3**, **Supplement Tables 4–6**). Line in the sex-stratified analyses, For VTE, MACE, and cardiovascular risk factors, ancestry-stratified risk patterns were generally consistent with the main analyses, although some ancestry-specific differences were observed. These, however, were not consistent across outcomes or AD trajectories (**Supplement Tables 4-6**).

## Conclusions and Relevance

In this large-scale, retrospective cohort study using real-world data from a nationally representative US EHR network, we demonstrate that persistent childhood AD is associated with a substantially elevated long-term risk of developing T2IDs, autoimmune diseases, and cardiometabolic risk factors. By contrast, children with transient AD showed only modest, almost negligible increases in risk relative to non-AD controls. Importantly, and in contrast to prior studies, our design allowed for direct head-to-head comparisons between persistent and transient AD trajectories, revealing consistent dose-response patterns across disease categories. These findings underscore that disease trajectory, not merely early presence or absence of AD in infancy and childhood, is a critical determinant of long-term systemic disease burden.

Our results build on and extend prior findings in several ways: Earlier work has established that AD, particularly when severe, is associated with comorbid allergic and autoimmune diseases (25). However, most studies assessed risk in relation to disease presence or severity, using non-AD individuals as a comparator, without directly comparing different AD trajectories. This approach limited insight into whether persistent disease independently confers additional risk. Moreover, previous studies often lacked sufficient sample sizes or follow-up durations to perform stratified analyses. By conducting all pairwise comparisons between persistent, transient, and no AD, we address a major gap in the literature. Our data confirm that persistent AD conveys markedly higher comorbidity risk than transient AD, including for asthma, food allergy, autoimmune disease, and cardiovascular risk factors. This aligns with and extends birth cohort studies (6,10), which identified early-life AD phenotypes predictive of later allergic diseases but were underpowered for head-to-head comparisons. Our findings reinforce the concept that longitudinal disease expression matters, with persistent immune activation potentially shaping systemic disease development.

Further, we contribute new evidence regarding cardiovascular risk. While we found no association between early childhood AD and MACE or VTE, we did observe small but significant increases in cardiometabolic risk factors, specifically obesity, hypertension, and dyslipidemia, among individuals with persistent AD. These findings are in line with recent birth cohort data (Lundin et al.) showing elevated BMI and blood pressure in young adults with longstanding AD. Though the effect sizes were modest, our data suggest that persistent AD may subtly shape metabolic risk trajectories, particularly in males.

We also observed notable disparities in EoE risk. Female patients with persistent or transient AD had significantly higher EoE risk than male patients. Furthermore, children with Black or African American ancestry had disproportionately elevated EoE risk following persistent AD, with hazard ratios exceeding those observed in White or Hispanic or Latino children. These findings suggest that both sex and ancestry may influence downstream inflammatory trajectories in select conditions, though such disparities were not observed consistently across other endpoints.

A major strength of this study is the use of a large, population-based EHR dataset, enabling robust comparisons between distinct childhood AD trajectories over long-term follow-up. The extensive PS matching and inclusion of diverse covariates (e.g., ancestry, socioeconomic status, family history) enhance validity and generalizability. However, several limitations must be acknowledged. First, the retrospective design and use of real-world data introduce potential bias and unmeasured confounding, including missing information on disease severity, treatment history, and environmental exposures. Although we adjusted for many variables, causal inferences cannot be drawn. Second, reliance on ICD-10 codes for exposures and outcomes may lead to misclassification due to inconsistent coding. Third, our trajectory definitions, persistent and transient, do not capture more nuanced disease patterns such as flares or recurrence. Fourth, differences in healthcare access may affect diagnostic coding and mask socioeconomic disparities. Lastly, some subgroup analyses, particularly EoE risk in ancestry-stratified groups, had wide confidence intervals and should be interpreted with caution. These limitations underscore the need for prospective studies integrating clinical and biological data to better define AD trajectories and their long-term health consequences. However, such comprehensive data remain challenging to collect at scale (26).

In the interim, our findings nonetheless have important implications for the risk stratification and management of children with AD: Our data suggest that persistent childhood AD is not only a chronic skin condition but a marker of broader systemic risk, including allergic, autoimmune, and cardiometabolic comorbidities. Conversely, children with transient AD appear to have minimal long-term systemic burden, supporting a more conservative management approach in this group. Yet, distinguishing between children likely to experience transient versus persistent disease is hardly possible. However, the striking differences in risk trajectories emphasize the need for early identification of persistent disease patterns and potentially earlier initiation of systemic or targeted therapy in children unresponsive to topical treatment.

## Supporting information

SROBE

## Data Availability

All data produced in the present work are contained in the manuscript

## Acknowledgements

We express our gratitude for the exceptional and ongoing support provided by Friederike Uebing (Justiziariat und Stabsstelle Vernetzung und Strategische Kooperation, UKSH, Campus Kiel) in facilitating access to the TriNetX platform at the UKSH.

