## Supplement Tables for "Distinct trajectories of childhood atopic dermatitis are associated with differences in long-term inflammatory and cardiometabolic disease risks"

| **Comparison** | **Outcome** | **Analysis** | **Characteristic** | **Before matching** | | | **After matching** | | |
| --- | --- | --- | --- | --- | --- | --- | --- | --- | --- |
|  |  |  |  | **Cases** | **Controls** | **Std. diff.** | **Cases** | **Controls** | **Std. diff.** |
| **Persistent (cases) versus transient AD (controls)** | **T2IDs** | **Primary** | Number of participants | 7,168 | 24,277 | - | 5,302 | 5,302 | - |
|  |  |  | Follow-up (days) median (interquartile range) | 5,252 (1,403) | 4,944 (1,297) | - | 5,252.5 (1,402) | 4,944 (1,302) | - |
|  |  |  | Age at Index (years, SD) | 0.762 ± 0.799 | 0.66 ± 0.772 | 0.1299 | 0.761 ± 0.799 | 0.762 ± 0.799 | 0.0007 |
|  |  |  | Female (%) | 51.828 | 45.115 | 0.1346 | 51.811 | 51.848 | 0.0008 |
|  |  |  | Male (%) | 48.074 | 54.768 | 0.1342 | 48.095 | 48.057 | 0.0008 |
|  |  |  | Black or African American | 43.649 | 29.789 | 0.2905 | 43.625 | 43.606 | 0.0004 |
|  |  |  | White (%) | 34.263 | 46.095 | 0.2431 | 34.289 | 34.289 | <0.0001 |
|  |  |  | Hispanic or Latino (%) | 17.282 | 18.268 | 0.0258 | 17.295 | 17.277 | 0.0005 |
|  |  |  | Persons with potential health hazards related to socioeconomic and psychosocial circumstances (Z55-Z65, %) | 0.641 | 0.706 | 0.0080 | 0.641 | 0.66 | 0.0023 |
|  |  |  | Overweight and obesity (E66, %) | 0.603 | 0.449 | 0.0080 | 0. 585 | 0.566 | 0.0025 |
|  |  |  | Family history of asthma and other chronic lower respiratory diseases (Z82.5, %) | 0.188 | 0.111 | 0.201 | 0.189 | 0.189 | <0.0001 |
|  |  |  | Nicotine dependence (F17, %) | 0.188 | 0.058 | 0.0371 | 0.189 | 0.189 | <0.0001 |
|  |  |  | Family history of diseases of the skin and subcutaneous tissue (Z84.0, %) | 0.188 | 0 | 0.0615 | 0 | 0 | <0.0001 |
|  |  | **S1** | Number of participants | 6,561 | 21,426 | - | 4,846 | 4,846 | - |
|  |  |  | Follow-up (days) median (interquartile range) | 5,345 (1,398) | 5,005 (1,340) | - | 5,245 (1,397) | 4,996.5 (1,360) | - |
|  |  |  | Age at Index (years, SD) | 0.767 ± 0.797 | 0.649 ± 0.77 | 0.1497 | 0.766 ± 0.797 | 0.766 ± 0.797 | 0.0003 |
|  |  |  | Female (%) | 51.659 | 44.535 | 0.1430 | 51.671 | 51.692 | 0.0004 |
|  |  |  | Male (%) | 48.32 | 55.459 | 0.1432 | 48.329 | 48.308 | 0.0004 |
|  |  |  | Black or African American | 42.301 | 30.883 | 0.2387 | 42.282 | 42.303 | 0.0004 |
|  |  |  | White (%) | 36.302 | 44.675 | 0.1712 | 36.339 | 36.319 | 0.0004 |
|  |  |  | Hispanic or Latino (%) | 17.254 | 18.604 | 0.0352 | 17.272 | 17.293 | 0.0005 |
|  |  |  | Persons with potential health hazards related to socioeconomic and psychosocial circumstances (Z55-Z65, %) | 0.639 | 0.68 | 0.0051 | 0.64 | 0.64 | <0.0001 |
|  |  |  | Overweight and obesity (E66, %) | 0.515 | 0.371 | 0.0218 | 0.516 | 0.475 | 0.0059 |
|  |  |  | Family history of asthma and other chronic lower respiratory diseases (Z82.5, %) | 0.206 | 0.115 | 0.0226 | 0.206 | 0.206 | <0.0001 |
|  |  |  | Nicotine dependence (F17, %) | 0.206 | 0.061 | 0.0398 | 0.206 | 0.206 | <0.0001 |
|  |  |  | Family history of diseases of the skin and subcutaneous tissue (Z84.0, %) | 0.206 | 0.061 | 0.0398 | 0 | 0 | - |
|  |  | **S2** | Number of participants | 5,552 | 18,338 | - | 4,989 | 4,989 | - |
|  |  |  | Follow-up (days) median (interquartile range) | 5,225.5 (1,385) | 4,985.5 (1,317) | - | 5,226 (1,383) | 4,971 (1,345) | - |
|  |  |  | Age at Index (years, SD) | 0.772 ± 0.799 | 0.649 ± 0.77 | 0.1570 | 0.772 ±  0.799 | 0.772 ± 0.799 | 0.0003 |
|  |  |  | Female (%) | 51.882 | 44.637 | 0.1454 | 51.894 | 51.934 | 0.0008 |
|  |  |  | Male (%) | 48.098 | 55.357 | 0.1457 | 48.106 | 48.066 | 0.0008 |
|  |  |  | Black or African American | 43.873 | 31.81 | 0.2507 | 43.856 | 43.856 | <0.0001 |
|  |  |  | White (%) | 34.992 | 43.945 | 0.1854 | 34.957 | 34.997 | 0.0008 |
|  |  |  | Hispanic or Latino (%) | 16.88 | 17.814 | 0.0247 | 16.897 | 16.837 | 0.0016 |
|  |  |  | Persons with potential health hazards related to socioeconomic and psychosocial circumstances (Z55-Z65, %) | 0.601 | 0.668 | 0.0085 | 0.601 | 0.581 | 0.0026 |
|  |  |  | Overweight and obesity (E66, %) | 0.501 | 0.358 | 0.0218 | 0.501 | 0.481 | 0.0029 |
|  |  |  | Family history of asthma and other chronic lower respiratory diseases (Z82.5, %) | 0.2 | 0.113 | 0.0220 | 0.2 | 0.2 | <0.0001 |
|  |  |  | Nicotine dependence (F17, %) | 0.2 | 0.06 | 0.0390 | 0.2 | 0.2 | <0.0001 |
|  |  |  | Family history of diseases of the skin and subcutaneous tissue (Z84.0, %) | 0.2 | 0.06 | 0.0390 | 0 | 0 | - |
|  |  | **Female** | Number of participants | 2,848 | 9,709 | - | 2,499 | 2,499 | - |
|  |  |  | Follow-up (days) median (interquartile range) | 5,325 (1,416) | 4,985 (1,335) | - | 5,327 (1,413) | 4,984 (1,331) | - |
|  |  |  | Age at Index (years, SD) | 0.814 ± 0.815 | 0.719 ± 0.788 | 0.1189 | 0.814± 0.815 | 0.814 ± 0.816 | 0.0005 |
|  |  |  | Female (%) | 100 | 100 | - | 100 | 100 | - |
|  |  |  | Male (%) | 0 | 0 | - | 0 | 0 | - |
|  |  |  | Black or African American | 46.781 | 31.027 | 0.3275 | 46.739 | 46.779 | 0.0008 |
|  |  |  | White (%) | 33.307 | 45.615 | 0.2539 | 33.333 | 33.333 | <0.0001 |
|  |  |  | Hispanic or Latino (%) | 16.753 | 18.037 | 0.0339 | 16.767 | 16.687 | 0.0021 |
|  |  |  | Persons with potential health hazards related to socioeconomic and psychosocial circumstances (Z55-Z65, %) | 0.44 | 0.59 | 0.0210 | 0.44 | 0.4 | 0.0062 |
|  |  |  | Overweight and obesity (E66, %) | 0.72 | 0.658 | 0.0075 | 0.72 | 0.76 | 0.0047 |
|  |  |  | Family history of asthma and other chronic lower respiratory diseases (Z82.5, %) | 0.4 | 0.216 | 0.0333 | 0.4 | 0.4 | <0.0001 |
|  |  |  | Nicotine dependence (F17, %) | 0 | 0.113 | 0.0477 | 0 | 0 | - |
|  |  |  | Family history of diseases of the skin and subcutaneous tissue (Z84.0, %) | 0.4 | 0.113 | 0.0566 | 0 | 0 | - |
|  |  | **Male** | Number of participants | 2,633 | 11,655 | - | 2,352 | 2,352 | - |
|  |  |  | Follow-up (days) median (interquartile range) | 5,230 (1,368) | 4,948 (1,292) | - | 5,230 (1,366) | 4,923.5 (1,258.5) | - |
|  |  |  | Age at Index (years, SD) | 0.69 ± 0.772 | 0.618 ± 0.758 | 0.0945 | 0.69 ± 0.772 | 0.69 ± 0.772 | 0.0011 |
|  |  |  | Female (%) | 0 | 0 | - | 0 | 0 | - |
|  |  |  | Male (%) | 100 | 100 | - | 100 | 100 | - |
|  |  |  | Black or African American | 42.523 | 29.643 | 0.2707 | 42.571 | 42.56 | 0.0009 |
|  |  |  | White (%) | 38.658 | 49.938 | 0.2286 | 38.69 | 38.69 | <0.0001 |
|  |  |  | Hispanic or Latino (%) | 16.058 | 19.453 | 0.0889 | 16.071 | 16.114 | 0.0012 |
|  |  |  | Persons with potential health hazards related to socioeconomic and psychosocial circumstances (Z55-Z65, %) | 0.807 | 0.625 | 0.0216 | 0.808 | 0.808 | <0.0001 |
|  |  |  | Overweight and obesity (E66, %) | 0.51 | 0.256 | 0.0412 | 0.51 | 0.51 | <0.0001 |
|  |  |  | Family history of asthma and other chronic lower respiratory diseases (Z82.5, %) | 0.425 | 0.256 | 0.0290 | 0.425 | 0.425 | <0.0001 |
|  |  |  | Nicotine dependence (F17, %) | 0.425 | 0.095 | 0.0649 | 0.425 | 0 | 0.0924 |
|  |  |  | Family history of diseases of the skin and subcutaneous tissue (Z84.0, %) | 0.425 | 0.095 | 0.0649 | 0 | 0 | - |
|  |  | **Black or African American** | Number of participants | 2,360 | 6,111 | - | 2,091 | 2,091 | - |
|  |  |  | Follow-up (days) median (interquartile range) | 5,345 (1,338) | 5,103 (1,311) | - | 5,346 (1,341) | 5,092 (1,318) | - |
|  |  |  | Age at Index (years, SD) | 0.707 ± 0.801 | 0.549 ± 0.742 | 0.2048 | 0.706 ± 0.801 | 0.706 ± 0.801 | <0.0001 |
|  |  |  | Female (%) | 55.18 | 47.82 | 0.1478 | 55.141 | 55.141 | <0.0001 |
|  |  |  | Male (%) | 44.816 | 52.184 | 0.1478 | 44.859 | 44.859 | <0.0001 |
|  |  |  | Black or African American | 100 | 100 | - | 100 | 100 | - |
|  |  |  | White (%) | 0 | 0 | - | 0 | 0 | - |
|  |  |  | Hispanic or Latino (%) | 2.962 | 3.056 | 0.0055 | 2.965 | 2.965 | <0.0001 |
|  |  |  | Persons with potential health hazards related to socioeconomic and psychosocial circumstances (Z55-Z65, %) | 1.051 | 1.204 | 0.0145 | 1.052 | 1.052 | <0.0001 |
|  |  |  | Overweight and obesity (E66, %) | 0.86 | 0.575 | 0.0337 | 0.861 | 0.861 | <0.0001 |
|  |  |  | Family history of asthma and other chronic lower respiratory diseases (Z82.5, %) | 0.478 | 0.306 | 0.0276 | 0.478 | 0.478 | <0.0001 |
|  |  |  | Nicotine dependence (F17, %) | 0.478 | 0.18 | 0.0521 | 0.478 | 0.0980 | 0.0924 |
|  |  |  | Family history of diseases of the skin and subcutaneous tissue (Z84.0, %) | 0.478 | 0.18 | 0.0521 | 0 | 0 | - |
|  |  | **Hispanic** | Number of participants | 987 | 3,548 | - | 905 | 905 | - |
|  |  |  | Follow-up (days) median (interquartile range) | 5,229 (1,428) | 4,750 (1,141) | - | 5,229 (1,427) | 4,688 (1,110) | - |
|  |  |  | Age at Index (years, SD) | 0.837 ± 0.806 | 0.731 ± 0.791 | 0.1320 | 0.836 ± 0.806 | 0.836 ± 0.805 | <0.0001 |
|  |  |  | Female (%) | 53.091 | 44.036 | 0.1819 | 53.039 | 53.149 | 0.0022 |
|  |  |  | Male (%) | 46.909 | 55.964 | 0.1819 | 46.961 | 46.851 | 0.0022 |
|  |  |  | Black or African American | 7.616 | 3.348 | 0.1883 | 7.514 | 7.514 | <0.0001 |
|  |  |  | White (%) | 49.117 | 55.725 | 0.1326 | 49.171 | 49.392 | 0.0044 |
|  |  |  | Hispanic or Latino (%) | 100 | 100 | - | 100 | 100 | - |
|  |  |  | Persons with potential health hazards related to socioeconomic and psychosocial circumstances (Z55-Z65, %) | 1.104 | 0.598 | 0.0551 | 1.105 | 1.105 | <0.0001 |
|  |  |  | Overweight and obesity (E66, %) | 1.104 | 0.658 | 0.0478 | 1.105 | 1.105 | <0.0001 |
|  |  |  | Family history of asthma and other chronic lower respiratory diseases (Z82.5, %) | 0 | 0.229 | 0.0774 | 0 | 0 | - |
|  |  |  | Nicotine dependence (F17, %) | 0 | 0.299 | 0.0774 | 0 | 0 | - |
|  |  |  | Family history of diseases of the skin and subcutaneous tissue (Z84.0, %) | 0 | 0.329 | 0.0812 | 0 | 0 | - |
|  |  | **White** | Number of participants | 2,054 | 8,415 | - | 1,837 | 1,837 | - |
|  |  |  | Follow-up (days) median (interquartile range) | 5,185 (1,837) | 4,831 (1,275) | - | 5,185 (1,445) | 4,887 (1,318) | - |
|  |  |  | Age at Index (years, SD) | 0.833 ± 0.789 | 0.771 ± 0.788 | 0.0782 | 0.833 ± 0.789 | 0.832 ± 0.789 | 0.0014 |
|  |  |  | Female (%) | 47.414 | 41.84 | 0.1123 | 47.414 | 47.469 | 0.0011 |
|  |  |  | Male (%) | 52.586 | 58.148 | 0.1121 | 52.586 | 52.531 | 0.0011 |
|  |  |  | Black or African American | 0 | 0 | - | 0 | 0 | - |
|  |  |  | White (%) | 100 | 100 | - | 100 | 100 | - |
|  |  |  | Hispanic or Latino (%) | 24.496 | 24.436 | 0.0014 | 24.496 | 24.66 | 0.0038 |
|  |  |  | Persons with potential health hazards related to socioeconomic and psychosocial circumstances (Z55-Z65, %) | 0.544 | 0.433 | 0.0159 | 0.544 | 0.544 | <0.0001 |
|  |  |  | Overweight and obesity (E66, %) | 0.544 | 0.268 | 0.0435 | 0.544 | 0.544 | <0.0001 |
|  |  |  | Family history of asthma and other chronic lower respiratory diseases (Z82.5, %) | 0.544 | 0.293 | 0.0389 | 0.544 | 0.544 | <0.0001 |
|  |  |  | Nicotine dependence (F17, %) | 0.544 | 0.127 | 0.0721 | 0.544 | 0.544 | <0.0001 |
|  |  |  | Family history of diseases of the skin and subcutaneous tissue (Z84.0, %) | 0 | 0.127 | 0.0505 | 0 | 0 | - |
| **Persistent (cases) versus no AD (controls)** | **T2IDs** | **Primary** | Number of participants | 7,124 | 147,233 | - | 5,250 | 5,250 | - |
|  |  |  | Follow-up (days) median (interquartile range) | 5,256  (1,399) | 4,901  (1,156) | - | 5,256  (1,399) | 4,807  (1,203) | - |
|  |  |  | Age at Index (years, SD) | 0.76 ± 0.799 | 0.567 ± 0.782 | 0.2439 | 0.758 ± 0.798 | 0.76 ± 0.799 | 0.0019 |
|  |  |  | Female (%) | 51.975 | 49.003 | 0.0595 | 51.924 | 51.943 | 0.0004 |
|  |  |  | Male (%) | 47.93 | 50.94 | 0.0602 | 47.981 | 47.981 | <0.0001 |
|  |  |  | Black or African American | 43.695 | 20.96 | 0.5011 | 43.619 | 43.562 | 0.0012 |
|  |  |  | White (%) | 34.087 | 55.949 | 0.4504 | 34.152 | 34.171 | 0.0004 |
|  |  |  | Hispanic or Latino (%) | 17.395 | 16.434 | 0.0256 | 17.41 | 17.429 | 0.0005 |
|  |  |  | Persons with potential health hazards related to socioeconomic and psychosocial circumstances (Z55-Z65, %) | 0.646 | 0.274 | 0.0550 | 0.648 | 0.667 | 0.0024 |
|  |  |  | Overweight and obesity (E66, %) | 0.608 | 0.044 | 0.0990 | 0.4 | 0.419 | 0.0030 |
|  |  |  | Family history of asthma and other chronic lower respiratory diseases (Z82.5, %) | 0.19 | 0.031 | 0.0479 | 0.19 | 0.19 | <0.0001 |
|  |  |  | Nicotine dependence (F17, %) | 0.19 | 0.009 | 0.0575 | 0 | 0 | - |
|  |  |  | Family history of diseases of the skin and subcutaneous tissue (Z84.0, %) | 0.19 | 0.009 | 0.0575 | 0 | 0 | - |
|  |  | **S1** | Number of participants | 7,100 | 153,281 | - | 5,305 | 5,305 | - |
|  |  |  | Follow-up (days) median (interquartile range) | 5,210 (1,365) | 4,890 (1,095) | - | 5,210 (1,107) | 4,798 (1,107) | - |
|  |  |  | Age at Index (years, SD) | 0.775 ± 0.803 | 0.61 ± 0.795 | 0.2056 | 0.775 ± 0.803 | 0.775 ± 0.803 | 0.0005 |
|  |  |  | Female (%) | 51.946 | 48.939 | 0.0602 | 51.932 | 51.913 | 0.0004 |
|  |  |  | Male (%) | 47.96 | 51.016 | 0.0612 | 47.974 | 47.992 | 0.0004 |
|  |  |  | Black or African American | 46.042 | 16.064 | 0.6848 | 45.919 | 45.957 | 0.0008 |
|  |  |  | White (%) | 33.258 | 59.698 | 0.5498 | 33.327 | 33.308 | 0.0004 |
|  |  |  | Hispanic or Latino (%) | 17.09 | 27.156 | 0.2443 | 17.135 | 17.097 | 0.0010 |
|  |  |  | Persons with potential health hazards related to socioeconomic and psychosocial circumstances (Z55-Z65, %) | 0.639 | 0.326 | 0.0452 | 0.641 | 0.641 | <0.0001 |
|  |  |  | Overweight and obesity (E66, %) | 0.583 | 0.052 | 0.0944 | 0.377 | 0.358 | 0.0031 |
|  |  |  | Family history of asthma and other chronic lower respiratory diseases (Z82.5, %) | 0.188 | 0.129 | 0.0149 | 0.189 | 0.189 | <0.0001 |
|  |  |  | Nicotine dependence (F17, %) | 0.188 | 0.008 | 0.0576 | 0 | 0 | - |
|  |  |  | Family history of diseases of the skin and subcutaneous tissue (Z84.0, %) | 0.188 | 0.012 | 0.0555 | 0.189 | 0 .189 | <0.0001 |
|  |  | **S2** | Number of participants | 5,894 | 134,318 | - | 5,310 | 5,310 | - |
|  |  |  | Follow-up (days) median (interquartile range) | 5,213 (1,366.5) | 4,891 (1,096) | - | 5,323.5 (1,366) | 4,784 (1,082) | - |
|  |  |  | Age at Index (years, SD) | 0.776 ± 0.803 | 0.611 ± 0.795 | 0.2064 | 0.776 ± 0.804 | 0.776 ± 0.804 | 0.0005 |
|  |  |  | Female (%) | 51.953 | 48.942 | 0.0602 | 51.94 | 51.921 | 0.0004 |
|  |  |  | Male (%) | 47.953 | 51.013 | 0.0612 | 47.966 | 47.985 | 0.0004 |
|  |  |  | Black or African American | 46.037 | 16.067 | 0.6846 | 45.913 | 45.951 | 0.0008 |
|  |  |  | White (%) | 33.264 | 59.699 | 0.5496 | 33.333 | 33.315 | 0.0004 |
|  |  |  | Hispanic or Latino (%) | 17.092 | 27.152 | 0.2442 | 17.137 | 17.1 | 0.0010 |
|  |  |  | Persons with potential health hazards related to socioeconomic and psychosocial circumstances (Z55-Z65, %) | 0.639 | 0.326 | 0.0451 | 0.64 | 0.64 | <0.0001 |
|  |  |  | Overweight and obesity (E66, %) | 0.582 | 0.052 | 0.0943 | 0.377 | 0.358 | 0.0031 |
|  |  |  | Family history of asthma and other chronic lower respiratory diseases (Z82.5, %) | 0.188 | 0.129 | 0.0149 | 0.188 | 0.188 | <0.0001 |
|  |  |  | Nicotine dependence (F17, %) | 0.188 | 0.008 | 0.0576 | 0 | 0 | - |
|  |  |  | Family history of diseases of the skin and subcutaneous tissue (Z84.0, %) | 0.188 | 0.012 | 0.0555 | 0.188 | 0.188 | <0.0001 |
|  |  | **Female** | Number of participants | 3,076 | 65,750 | - | 2,758 | 2,758 | - |
|  |  |  | Follow-up (days) median (interquartile range) | 5,236 (1,406) | 4,911 (1,116) | - | 5,236 (1,405) | 4,808 (1,113) | - |
|  |  |  | Age at Index (years, SD) | 0.84 ± 0.82 | 0.612 ± 0.797 | 0.2819 | 0.84 ± 0.821 | 0.84 ± 0.821 | 0.0004 |
|  |  |  | Female (%) | 100 | 100 | - | 100 | 100 | - |
|  |  |  | Male (%) | 0 | 0 | - | 0 | 0 | - |
|  |  |  | Black or African American | 49.024 | 16.093 | 0.7506 | 48.876 | 48.912 | 0.0007 |
|  |  |  | White (%) | 30.513 | 59.738 | 0.6144 | 30.602 | 30.566 | 0.0008 |
|  |  |  | Hispanic or Latino (%) | 17.462 | 27.116 | 0.2335 | 17.513 | 17.44 | 0.0019 |
|  |  |  | Persons with potential health hazards related to socioeconomic and psychosocial circumstances (Z55-Z65, %) | 0.47 | 0.298 | 0.0278 | 0.471 | 0.471 | <0.0001 |
|  |  |  | Overweight and obesity (E66, %) | 0.687 | 0.048 | 0.1058 | 0.435 | 0.399 | 0.0056 |
|  |  |  | Family history of asthma and other chronic lower respiratory diseases (Z82.5, %) | 0.362 | 0.128 | 0.0474 | 0.363 | 0.363 | <0.0001 |
|  |  |  | Nicotine dependence (F17, %) | 0 | 0 | - | 0 | 0 | - |
|  |  |  | Family history of diseases of the skin and subcutaneous tissue (Z84.0, %) | 0.362 | 0.016 | 0.0797 | 0.363 | 0.363 | <0.0001 |
|  |  | **Male** | Number of participants | 2,813 | 68,511 | - | 2,547 | 2,547 | - |
|  |  |  | Follow-up (days) median (interquartile range) | 5,181 (1,333) | 4,872 (1,076) | - | 5,181 (1,327) | 4,809 (1,047) | - |
|  |  |  | Age at Index (years, SD) | 0.707 ± 0.779 | 0.609 ± 0.794 | 0.1244 | 0.707 ± 0.779 | 0.707 ± 0.779 | <0.0001 |
|  |  |  | Female (%) | 0 | 0 | - | 0 | 0 | - |
|  |  |  | Male (%) | 100 | 100 | - | 100 | 100 | - |
|  |  |  | Black or African American | 42.891 | 16.038 | 0.6164 | 42.795 | 42.835 | 0.0008 |
|  |  |  | White (%) | 36.31 | 59.678 | 0.4811 | 36.356 | 36.356 | <0.0001 |
|  |  |  | Hispanic or Latino (%) | 16.725 | 27.207 | 0.2552 | 16.765 | 16.804 | 0.0011 |
|  |  |  | Persons with potential health hazards related to socioeconomic and psychosocial circumstances (Z55-Z65, %) | 0.823 | 0.353 | 0.0614 | 0.824 | 0.824 | <0.0001 |
|  |  |  | Overweight and obesity (E66, %) | 0.47 | 0.057 | 0.0807 | 0.393 | 0.393 | <0.0001 |
|  |  |  | Family history of asthma and other chronic lower respiratory diseases (Z82.5, %) | 0.392 | 0.13 | 0.0513 | 0.393 | 0.393 | <0.0001 |
|  |  |  | Nicotine dependence (F17, %) | 0.392 | 0.015 | 0.0836 | 0 | 0 | - |
|  |  |  | Family history of diseases of the skin and subcutaneous tissue (Z84.0, %) | 0.392 | 0.13 | 0.0513 | 0.393 | 0.393 | <0.0001 |
|  |  | **Black or African American** | Number of participants | 2,734 | 21,361 | - | 2,438 | 2,438 | - |
|  |  |  | Follow-up (days) median (interquartile range) | 5,277 (1,354) | 4,723 (1,001) | - | 5,277.5 (1,353) | 4,688 (1,041) | - |
|  |  |  | Age at Index (years, SD) | 0.734 ± 0.805 | 0.569 ± 0.827 | 0.0540 | 0.734 ± 0.805 | 0.734 ± 0.805 | <0.0001 |
|  |  |  | Female (%) | 55.324 | 49.024 | 0.1264 | 55.291 | 55.25 | 0.0008 |
|  |  |  | Male (%) | 44.676 | 50.923 | 0.1253 | 44.709 | 44.75 | 0.0008 |
|  |  |  | Black or African American | 100 | 100 | - | 100 | 100 | - |
|  |  |  | White (%) | 0 | 0 | - | 0 | 0 | - |
|  |  |  | Hispanic or Latino (%) | 2.774 | 3.608 | 0.0474 | 2.789 | 2.748 | 0.0025 |
|  |  |  | Persons with potential health hazards related to socioeconomic and psychosocial circumstances (Z55-Z65, %) | 0.979 | 0.592 | 0.0438 | 0.984 | 0.984 | <0.0001 |
|  |  |  | Overweight and obesity (E66, %) | 0.734 | 0.053 | 0.1089 | 0.41 | 0.41 | <0.0001 |
|  |  |  | Family history of asthma and other chronic lower respiratory diseases (Z82.5, %) | 0.408 | 0.325 | 0.0137 | 0.41 | 0.41 | <0.0001 |
|  |  |  | Nicotine dependence (F17, %) | 0.408 | 0 | 0.0905 | 0 | 0 | - |
|  |  |  | Family history of diseases of the skin and subcutaneous tissue (Z84.0, %) | 0.408 | 0.049 | 0.0754 | 0.41 | 0.41 | <0.0001 |
|  |  | **Hispanic** | Number of participants | 993 | 35,910 | - | 912 | 912 | - |
|  |  |  | Follow-up (days) median (interquartile range) | 5,226.5 (1,433) | 4,836 (1,015) | - | 5,226.5 (1,433) | 4,761.5 (1,013) | - |
|  |  |  | Age at Index (years, SD) | 0.834 ± 0.806 | 0.581 ± 0.79 | 0.3179 | 0.834 ± 0.806 | 0.836 ± 0.807 | 0.0014 |
|  |  |  | Female (%) | 53.18 | 48.913 | 0.0854 | 53.18 | 52.961 | 0.0044 |
|  |  |  | Male (%) | 46.82 | 51.082 | 0.0853 | 46.82 | 47.039 | 0.0044 |
|  |  |  | Black or African American | 7.566 | 2.237 | 0.2487 | 7.566 | 7.566 | <0.0001 |
|  |  |  | White (%) | 49.013 | 64.337 | 0.3130 | 49.013 | 49.013 | <0.0001 |
|  |  |  | Hispanic or Latino (%) | 100 | 100 | - | 100 | 100 | - |
|  |  |  | Persons with potential health hazards related to socioeconomic and psychosocial circumstances (Z55-Z65, %) | 1.096 | 0.313 | 0.0938 | 1.096 | 1.096 | <0.0001 |
|  |  |  | Overweight and obesity (E66, %) | 1.096 | 0.117 | 0.1265 | 1.096 | 1.096 | <0.0001 |
|  |  |  | Family history of asthma and other chronic lower respiratory diseases (Z82.5, %) | 0 | 0.179 | 0.0599 | 0 | 0 | - |
|  |  |  | Nicotine dependence (F17, %) | 0 | 0 | - | 0 | 0 | - |
|  |  |  | Family history of diseases of the skin and subcutaneous tissue (Z84.0, %) | 0 | 0.028 | 0.0238 | 0 | 0 | - |
|  |  | **White** | Number of participants | 1,997 | 85,608 | - | 1,802 | 1,802 | - |
|  |  |  | Follow-up (days) median (interquartile range) | 5,158 (1,427) | 4,873 (1,128) | - | 5,158 (1,426) | 4,759 (1,187) | - |
|  |  |  | Age at Index (years, SD) | 0.835 ± 0.789 | 0.624 ± 0.799 | 0.2661 | 0.836 ± 0.789 | 0.836 ± 0.789 | <0.0001 |
|  |  |  | Female (%) | 47.284 | 49.021 | 0.0348 | 47.218 | 47.281 | <0.0001 |
|  |  |  | Male (%) | 52.716 | 50.951 | 0.0353 | 52.719 | 52.719 | <0.0001 |
|  |  |  | Black or African American | 0 | 0 | - | 0 | 0 | - |
|  |  |  | White (%) | 100 | 100 | - | 100 | 100 | - |
|  |  |  | Hispanic or Latino (%) | 24.778 | 24.654 | 0.0654 | 24.75 | 24.75 | <0.0001 |
|  |  |  | Persons with potential health hazards related to socioeconomic and psychosocial circumstances (Z55-Z65, %) | 0.554 | 0.265 | 0.0453 | 0.555 | 0.555 | <0.0001 |
|  |  |  | Overweight and obesity (E66, %) | 0.554 | 0.042 | 0.0942 | 0.555 | 0.555 | <0.0001 |
|  |  |  | Family history of asthma and other chronic lower respiratory diseases (Z82.5, %) | 0.554 | 0.095 | 0.0807 | 0.555 | 0.555 | <0.0001 |
|  |  |  | Nicotine dependence (F17, %) | 0.554 | 0.012 | 0.1021 | 0 | 0 | - |
|  |  |  | Family history of diseases of the skin and subcutaneous tissue (Z84.0, %) | 0 | 0.012 | 0.0156 | 0 | 0 | - |
| **Transient (cases) versus no AD (controls)** | **T2IDs** | **Primary** | Number of participants | 20,486 | 139,542 | - | 13,950 | 13,950 | - |
|  |  |  | Follow-up (days) median (interquartile range) | 4,978 (1,365) | 4,792 (1,178) | - | 4,979 (1,365) | 4,765 (1,178) | - |
|  |  |  | Age at Index (years, SD) | 0.652 ± 0.769 | 0.62 ± 0.804 | 0.0411 | 0.651 ± 0.768 | 0.651 ± 0.769 | <0.0001 |
|  |  |  | Female (%) | 45.051 | 49.109 | 0.0814 | 44.975 | 44.982 | 0.0001 |
|  |  |  | Male (%) | 54.806 | 50.831 | 0.0797 | 54.882 | 54.91 | 0.0006 |
|  |  |  | Black or African American | 29.122 | 22.846 | 0.1435 | 29.061 | 29.075 | 0.0003 |
|  |  |  | White (%) | 46.044 | 55.165 | 0.1832 | 46.108 | 46.143 | 0.0007 |
|  |  |  | Hispanic or Latino (%) | 15.994 | 15.811 | 0.0050 | 15.971 | 15.95 | 0.0006 |
|  |  |  | Persons with potential health hazards related to socioeconomic and psychosocial circumstances (Z55-Z65, %) | 0.672 | 0.216 | 0.0686 | 0.659 | 0.71 | 0.0061 |
|  |  |  | Overweight and obesity (E66, %) | 0.493 | 0.041 | 0.0878 | 0.229 | 0.229 | <0.0001 |
|  |  |  | Family history of asthma and other chronic lower respiratory diseases (Z82.5, %) | 0.079 | 0.028 | 0.0218 | 0.079 | 0.072 | 0.0026 |
|  |  |  | Nicotine dependence (F17, %) | 0.071 | 0.009 | 0.0309 | 0 | 0 | - |
|  |  |  | Family history of diseases of the skin and subcutaneous tissue (Z84.0, %) | 0 | 0.009 | 0.0137 | 0 | 0 | - |
|  |  | **S1** | Number of participants | 20,062 | 166,664 | - | 16,229 | 16,229 | - |
|  |  |  | Follow-up (days) median (interquartile range) | 4,840 (1,173) | 4,937 (1,172) | - | 4,840 (1,172) | 4,680 (1,138) | - |
|  |  |  | Age at Index (years, SD) | 0.688 ± 0.78 | 0.602 ± 0.792 | 0.1088 | 0.688 ± 0.78 | 0.688 ± 0.78 | 0.0006 |
|  |  |  | Female (%) | 44.222 | 48.887 | 0.0936 | 44.199 | 44.162 | 0.0007 |
|  |  |  | Male (%) | 55.655 | 51.067 | 0.0921 | 55.678 | 55.703 | 0.0005 |
|  |  |  | Black or African American | 31.644 | 18.229 | 0.3139 | 31.592 | 31.653 | 0.0013 |
|  |  |  | White (%) | 44.197 | 58.88 | 0.2970 | 44.236 | 44.192 | 0.0009 |
|  |  |  | Hispanic or Latino (%) | 21.311 | 25.377 | 0.0962 | 21.314 | 21.301 | 0.0003 |
|  |  |  | Persons with potential health hazards related to socioeconomic and psychosocial circumstances (Z55-Z65, %) | 0.584 | 0.307 | 0.0417 | 0.567 | 0.561 | 0.0008 |
|  |  |  | Overweight and obesity (E66, %) | 0.387 | 0.051 | 0.0720 | 0.222 | 0.203 | 0.0040 |
|  |  |  | Family history of asthma and other chronic lower respiratory diseases (Z82.5, %) | 0.264 | 0.139 | 0.0280 | 0.246 | 0.24 | 0.0013 |
|  |  |  | Nicotine dependence (F17, %) | 0.062 | 0.007 | 0.0293 | 0.062 | 0.062 | <0.0001 |
|  |  |  | Family history of diseases of the skin and subcutaneous tissue (Z84.0, %) | 0.062 | 0.012 | 0.0262 | 0.062 | 0 | 0.0351 |
|  |  | **S2** | Number of participants | 16,303 | 142,711 | - | 15,325 | 15,325 | - |
|  |  |  | Follow-up (days) median (interquartile range) | 4,831 (1,163) | 4,947 (1,173) | - | 4,831 (1,162) | 4,894 (1,162) | - |
|  |  |  | Age at Index (years, SD) | 0.695 ± 0.782 | 0.597 ± 0.79 | 0.1247 | 0.695 ± 0.782 | 0.696 ± 0.782 | 0.0017 |
|  |  |  | Female (%) | 44.219 | 48.885 | 0.0937 | 44.176 | 44.15 | 0.0005 |
|  |  |  | Male (%) | 55.651 | 51.072 | 0.0919 | 55.693 | 55.7 | 0.0001 |
|  |  |  | Black or African American | 30.389 | 18.369 | 0.2827 | 30.369 | 30.369 | <0.0001 |
|  |  |  | White (%) | 45.639 | 58.802 | 0.2658 | 45.657 | 45.644 | 0.0003 |
|  |  |  | Hispanic or Latino (%) | 21.262 | 25.531 | 0.1010 | 21.246 | 21.253 | 0.0002 |
|  |  |  | Persons with potential health hazards related to socioeconomic and psychosocial circumstances (Z55-Z65, %) | 0.554 | 0.312 | 0.0368 | 0.542 | 0.522 | 0.0027 |
|  |  |  | Overweight and obesity (E66, %) | 0.358 | 0.051 | 0.0681 | 0.189 | 0.183 | 0.0015 |
|  |  |  | Family history of asthma and other chronic lower respiratory diseases (Z82.5, %) | 0.28 | 0.14 | 0.0305 | 0.268 | 0.235 | 0.0065 |
|  |  |  | Nicotine dependence (F17, %) | 0.065 | 0.007 | 0.0304 | 0.065 | 0 | 0.0361 |
|  |  |  | Family history of diseases of the skin and subcutaneous tissue (Z84.0, %) | 0.065 | 0.012 | 0.0272 | 0.065 | 0.065 | <0.0001 |
|  |  | **Female** | Number of participants | 7,291 | 71,482 | - | 6,840 | 6,840 | - |
|  |  |  | Follow-up (days) median (interquartile range) | 4,830 (1,181) | 4,959 (1,200) | - | 4,830.5 (1,183.5) | 4,905 (1,204) | - |
|  |  |  | Age at Index (years, SD) | 0.76 ± 0.798 | 0.606 ± 0.794 | 0.1938 | 0.76 ± 0.798 | 0.76 ± 0.797 | 0.0002 |
|  |  |  | Female (%) | 100 | 100 | - | 100 | 100 | - |
|  |  |  | Male (%) | 0 | 0 | - | 0 | 0 | - |
|  |  |  | Black or African American | 32.406 | 18.284 | 0.3290 | 32.383 | 32.368 | 0.0003 |
|  |  |  | White (%) | 43.596 | 58.869 | 0.3092 | 43.626 | 43.64 | 0.0003 |
|  |  |  | Hispanic or Latino (%) | 21.011 | 25.301 | 0.1018 | 20.98 | 20.98 | <0.0001 |
|  |  |  | Persons with potential health hazards related to socioeconomic and psychosocial circumstances (Z55-Z65, %) | 0.481 | 0.282 | 0.0322 | 0.453 | 0.439 | 0.0022 |
|  |  |  | Overweight and obesity (E66, %) | 0.568 | 0.047 | 0.0942 | 0.278 | 0.278 | <0.0001 |
|  |  |  | Family history of asthma and other chronic lower respiratory diseases (Z82.5, %) | 0.248 | 0.141 | 0.0242 | 0.175 | 0.19 | 0.0034 |
|  |  |  | Nicotine dependence (F17, %) | 0 | 0 | - | 0 | 0 | - |
|  |  |  | Family history of diseases of the skin and subcutaneous tissue (Z84.0, %) | 0.146 | 0.015 | 0.0463 | 0 | 0.146 | 0.541 |
|  |  | **Male** | Number of participants | 7,596 | 64,854 | - | 7,184 | 7,184 | - |
|  |  |  | Follow-up (days) median (interquartile range) | 4,836 (1,146) | 4,992 (1,205) | - | 4,836 (1.145.5) | 4,963 (1,215) | - |
|  |  |  | Age at Index (years, SD) | 0.635 ± 0.762 | 0.599 ± 0.788 | 0.0462 | 0.635 ± 0.762 | 0.635 ± 0.762 | <0.0001 |
|  |  |  | Female (%) | 0 | 0 | - | 0 | 0 | - |
|  |  |  | Male (%) | 100 | 100 | - | 100 | 100 | - |
|  |  |  | Black or African American | 27.771 | 16.423 | 0.2761 | 27.742 | 27.742 | <0.0001 |
|  |  |  | White (%) | 48.06 | 61.197 | 0.2662 | 48.065 | 48.079 | 0.0003 |
|  |  |  | Hispanic or Latino (%) | 16.187 | 15.017 | 0.0322 | 16.203 | 16.189 | 0.0004 |
|  |  |  | Persons with potential health hazards related to socioeconomic and psychosocial circumstances (Z55-Z65, %) | 0.528 | 0.268 | 0.0413 | 0.501 | 0.501 | <0.0001 |
|  |  |  | Overweight and obesity (E66, %) | 0.139 | 0.049 | 0.0295 | 0.139 | 0.139 | <0.0001 |
|  |  |  | Family history of asthma and other chronic lower respiratory diseases (Z82.5, %) | 0.139 | 0.052 | 0.0282 | 0.139 | 0.139 | <0.0001 |
|  |  |  | Nicotine dependence (F17, %) | 0.139 | 0.016 | 0.0441 | 0 | 0 | - |
|  |  |  | Family history of diseases of the skin and subcutaneous tissue (Z84.0, %) | 0 | 0.016 | 0.0180 | 0 | 0 | - |
|  |  | **Black or African American** | Number of participants | 4,020 | 20,815 | - | 3,785 | 3,785 | - |
|  |  |  | Follow-up (days) median (interquartile range) | 4,901 (1,176) | 4,876 (1,218) | - | 4,901 (1,177) | 4,852 (1,174) | - |
|  |  |  | Age at Index (years, SD) | 0.589 ± 0.759 | 0.631 ± 0.81 | 0.0527 | 0.589 ± 0.759 | 0.589 ± 0.759 | 0.0007 |
|  |  |  | Female (%) | 47.35 | 49.087 | 0.0348 | 47.318 | 47.345 | 0.0005 |
|  |  |  | Male (%) | 52.65 | 50.858 | 0.0359 | 52.682 | 52.655 | 0.0005 |
|  |  |  | Black or African American | 100 | 100 | - | 100 | 100 | - |
|  |  |  | White (%) | 0 | 0 | - | 0 | 0 | - |
|  |  |  | Hispanic or Latino (%) | 2.426 | 4.544 | 0.1157 | 2.431 | 2.404 | 0.0017 |
|  |  |  | Persons with potential health hazards related to socioeconomic and psychosocial circumstances (Z55-Z65, %) | 0.923 | 0.458 | 0.0562 | 0.872 | 0.845 | 0.0029 |
|  |  |  | Overweight and obesity (E66, %) | 0.369 | 0.055 | 0.0682 | 0.264 | 0.264 | <0.0001 |
|  |  |  | Family history of asthma and other chronic lower respiratory diseases (Z82.5, %) | 0.264 | 0.111 | 0.0354 | 0.264 | 0.264 | <0.0001 |
|  |  |  | Nicotine dependence (F17, %) | 0 | 0 | - | 0 | 0 | - |
|  |  |  | Family history of diseases of the skin and subcutaneous tissue (Z84.0, %) | 0 | 0.05 | 0.0317 | 0 | 0 | - |
|  |  | **Hispanic** | Number of participants | 2,038 | 21,075 | - | 1,815 | 1,815 | - |
|  |  |  | Follow-up (days) median (interquartile range) | 4,940 (1,256) | 4,987 (986) | - | 4,945 (1,258) | 4,954 (1,065) | - |
|  |  |  | Age at Index (years, SD) | 0.708 ± 0.788 | 0.435 ± 0.728 | 0.33579 | 0.704 ± 0.786 | 0.705 ± 0.786 | 0.0014 |
|  |  |  | Female (%) | 45.37 | 48.832 | 0.0694 | 45.234 | 45.289 | 0.0011 |
|  |  |  | Male (%) | 54.63 | 51.168 | 0.0694 | 54.766 | 54.711 | 0.0011 |
|  |  |  | Black or African American | 5.918 | 2.778 | 0.1544 | 5.785 | 5.785 | <0.0001 |
|  |  |  | White (%) | 45.753 | 64.76 | 0.3894 | 45.73 | 45.785 | 0.0011 |
|  |  |  | Hispanic or Latino (%) | 100 | 100 | - | 100 | 100 | - |
|  |  |  | Persons with potential health hazards related to socioeconomic and psychosocial circumstances (Z55-Z65, %) | 0.548 | 0.3 | 0.0381 | 0.551 | 0.551 | <0.0001 |
|  |  |  | Overweight and obesity (E66, %) | 0.712 | 0.061 | 0.1051 | 0.551 | 0.551 | <0.0001 |
|  |  |  | Family history of asthma and other chronic lower respiratory diseases (Z82.5, %) | 0.548 | 0.051 | 0.0911 | 0.551 | 0.551 | <0.0001 |
|  |  |  | Nicotine dependence (F17, %) | 0.548 | 0 | 0.1050 | 0 | 0 | - |
|  |  |  | Family history of diseases of the skin and subcutaneous tissue (Z84.0, %) | 0 | 0.051 | 0.0319 | 0 | 0 | - |
|  |  | **White** | Number of participants | 6,103 | 61,167 | - | 5,437 | 5,437 | - |
|  |  |  | Follow-up (days) median (interquartile range) | 5,068.5 (1,366) | 5,088.5 (1,019) | - | 5,072 (1,367) | 5,049 (1,031) | - |
|  |  |  | Age at Index (years, SD) | 0.678 ± 0.766 | 0.448 ± 0.728 | 0.3075 | 0.676 ± 0.765 | 0.676 ± 0.765 | <0.0001 |
|  |  |  | Female (%) | 43.766 | 49.013 | 0.1054 | 43.664 | 43.682 | 0.0004 |
|  |  |  | Male (%) | 56.216 | 50.982 | 0.1051 | 56.318 | 56.299 | 0.0004 |
|  |  |  | Black or African American | 0 | 0 | - | 0 | 0 | - |
|  |  |  | White (%) | 100 | 100 | - | 100 | 100 | - |
|  |  |  | Hispanic or Latino (%) | 15.31 | 22.471 | 0.1827 | 15.247 | 15.266 | 0.0005 |
|  |  |  | Persons with potential health hazards related to socioeconomic and psychosocial circumstances (Z55-Z65, %) | 0.532 | 0.272 | 0.0411 | 0.533 | 0.533 | <0.0001 |
|  |  |  | Overweight and obesity (E66, %) | 0.367 | 0.03 | 0.0757 | 0.184 | 0.184 | <0.0001 |
|  |  |  | Family history of asthma and other chronic lower respiratory diseases (Z82.5, %) | 0.183 | 0.018 | 0.0523 | 0.184 | 0.184 | <0.0001 |
|  |  |  | Nicotine dependence (F17, %) | 0.183 | 0 | 0.0606 | 0 | 0 | - |
|  |  |  | Family history of diseases of the skin and subcutaneous tissue (Z84.0, %) | 0 | 0.018 | 0.0188 | 0 | 0 | - |

**Supplement Table 1**. Baseline characteristics before and after propensity-score matching for all analyses relating to the risk of type 2 inflammatory diseases (T2IDs) in patients with persistent, transient or no childhood atopic dermatitis (AD).

| **Comparison** | **Outcome** | **Analysis** | **Characteristic** | **Before matching** | | | **After matching** | | |
| --- | --- | --- | --- | --- | --- | --- | --- | --- | --- |
|  |  |  |  | **Cases** | **Controls** | **Std. diff.** | **Cases** | **Controls** | **Std. diff.** |
| **Persistent (cases) versus transient AD (controls)** | **Autoimmune diseases** | **Primary** | Number of participants | 7,013 | 24,812 | - | 5,158 | 5,158 | - |
|  |  |  | Follow-up (days) median (interquartile range) | 5,263 (1,414) | 4,946 (1,316) | - | 5,263 (1,414) | 4,948 (1,343) | - |
|  |  |  | Age at Index (years, SD) | 0.76 ± 0.798 | 0.666 ± 0.774 | 0.1198 | 0.76 ± 0.798 | 0.76 ± 0.799 | <0.0001 |
|  |  |  | Female (%) | 51.919 | 44.692 | 0.1450 | 51.919 | 51.881 | 0.0001 |
|  |  |  | Male (%) | 47.984 | 55.302 | 0.1468 | 47.984 | 48.119 | 0.0006 |
|  |  |  | Black or African American | 44.203 | 31.446 | 0.2654 | 44.203 | 44.184 | 0.0003 |
|  |  |  | White (%) | 33.501 | 42.011 | 0.1762 | 33.501 | 33.501 | 0.0007 |
|  |  |  | Hispanic or Latino (%) | 17.429 | 18.726 | 0.0337 | 17.429 | 17.429 | 0.0006 |
|  |  |  | Persons with potential health hazards related to socioeconomic and psychosocial circumstances (Z55-Z66, %) | 0.62 | 0.758 | 0.0166 | 0.62 | 0.62 | 0.0061 |
|  |  |  | Overweight and obesity (E66, %) | 0.62 | 0.486 | 0.0181 | 0.62 | 0.601 | <0.0001 |
|  |  |  | Family history of other endocrine, nutritional and metabolic diseases (Z83.4, %) | 0.194 | 0.057 | 0.0388 | 0.194 | 0 | 0.0026 |
|  |  |  | Nicotine dependence (F17, %) | 0.194 | 0.057 | 0.0388 | 0.194 | 0.194 | - |
|  |  |  | Family history of arthritis and other diseases of the musculoskeletal sytem and connective tissue (Z82.6, %) | 0.194 | 0.057 | 0.0388 | 0.194 | 0.194 | - |
|  |  | **S1** | Number of participants | 7,336 | 17,483 | - | 5,357 | 5,357 | - |
|  |  |  | Follow-up (days) median (interquartile range) | 5,212 (1,402) | 5,051 (1,376) | - | 5,222 (1,402) | 5,042 (1,407) | - |
|  |  |  | Age at Index (years, SD) | 0.77 ± 0.8 | 0.65 ± 0.767 | 0.1538 | 0.754 ± 0.792 | 0.754 ± 0.793 | 0.0007 |
|  |  |  | Female (%) | 51.906 | 45.074 | 0.1370 | 51.279 | 51.279 | 0.0004 |
|  |  |  | Male (%) | 48.002 | 54.917 | 0.1387 | 48.628 | 48.684 | 0.0011 |
|  |  |  | Black or African American | 45.185 | 29.79 | 0.3221 | 44.465 | 44.428 | 0.0008 |
|  |  |  | White (%) | 34.414 | 48.647 | 0.2919 | 34.852 | 34.926 | 0.0016 |
|  |  |  | Hispanic or Latino (%) | 15.743 | 16.302 | 0.0152 | 15.96 | 15.886 | 0.0020 |
|  |  |  | Persons with potential health hazards related to socioeconomic and psychosocial circumstances (Z55-Z66, %) | 0.571 | 0.67 | 0.0126 | 0.579 | 0.541 | 0.0050 |
|  |  |  | Overweight and obesity (E66, %) | 0.589 | 0.473 | 0.0159 | 0.541 | 0.485 | 0.0078 |
|  |  |  | Family history of other endocrine, nutritional and metabolic diseases (Z83.4, %) | 0.184 | 0.089 | 0.0257 | 0.187 | 0 | 0.0612 |
|  |  |  | Nicotine dependence (F17, %) | 0.184 | 0.089 | 0.0257 | 0.187 | 0 | 0.0612 |
|  |  |  | Family history of arthritis and other diseases of the musculoskeletal sytem and connective tissue (Z82.6, %) | 0.184 | 0.089 | 0.0257 | 0 | 0 | - |
|  |  | **S2** | Number of participants | 5,995 | 12,213 | - | 5,266 | 5,266 | - |
|  |  |  | Follow-up (days) median (interquartile range) | 5,224 (1,416) | 5,069 (1,389) | - | 5,227 (1,414) | 5,073 (1,424) | - |
|  |  |  | Age at Index (years, SD) | 0.769 ± 0.8 | 0.647 ± 0.766 | 0.1553 | 0.749 ± 0.791 | 0.75 ± 0.792 | 0.0012 |
|  |  |  | Female (%) | 51.821 | 45.027 | 0.1363 | 51.044 | 51.139 | 0.0019 |
|  |  |  | Male (%) | 48.085 | 54.964 | 0.1380 | 48.861 | 48.842 | 0.0004 |
|  |  |  | Black or African American | 44.76 | 29.038 | 0.3302 | 43.866 | 43.828 | 0.0008 |
|  |  |  | White (%) | 34.766 | 49.472 | 0.3012 | 35.321 | 35.378 | 0.0012 |
|  |  |  | Hispanic or Latino (%) | 15.991 | 16.732 | 0.0200 | 16.255 | 16.236 | 0.0005 |
|  |  |  | Persons with potential health hazards related to socioeconomic and psychosocial circumstances (Z55-Z66, %) | 0.579 | 0.689 | 0.0138 | 0.589 | 0.57 | 0.0025 |
|  |  |  | Overweight and obesity (E66, %) | 0.598 | 0.487 | 0.0151 | 0.57 | 0.494 | 0.0104 |
|  |  |  | Family history of other endocrine, nutritional and metabolic diseases (Z83.4, %) | 0.187 | 0.092 | 0.0255 | 0.19 | 0 | 0.0617 |
|  |  |  | Nicotine dependence (F17, %) | 0.187 | 0.092 | 0.0255 | 0.19 | 0.19 | <0.0001 |
|  |  |  | Family history of arthritis and other diseases of the musculoskeletal sytem and connective tissue (Z82.6, %) | 0.187 | 0.092 | 0.0255 | 0.19 | 0 | 0.0617 |
|  |  | **Female** | Number of participants | 3,173 | 5,672 | - | 2,753 | 2,753 | - |
|  |  |  | Follow-up (days) median (interquartile range) | 5,245 (1,448) | 5,064.5 (1,404.5) | - | 5,253 (1,451) | 5,026 (1,419) | - |
|  |  |  | Age at Index (years, SD) | 0.833 ± 0.816 | 0.711 ± 0.786 | 0.1520 | 0.806 ± 0.806 | 0.804 ± 0.805 | 0.0023 |
|  |  |  | Female (%) | 100 | 100 | - | 100 | 100 | - |
|  |  |  | Male (%) | 0 | 0 | - | 0 | 0 | - |
|  |  |  | Black or African American | 48.033 | 31.131 | 0.3509 | 46.785 | 46.713 | 0.0015 |
|  |  |  | White (%) | 31.514 | 47.35 | 0.3284 | 32.256 | 32.219 | 0.0008 |
|  |  |  | Hispanic or Latino (%) | 16.058 | 16.357 | 0.0081 | 16.309 | 16,527 | 0.0059 |
|  |  |  | Persons with potential health hazards related to socioeconomic and psychosocial circumstances (Z55-Z66, %) | 0.425 | 0.692 | 0.0358 | 0.436 | 0.436 | <0.0001 |
|  |  |  | Overweight and obesity (E66, %) | 0.674 | 0.771 | 0.0116 | 0.654 | 0.618 | 0.0046 |
|  |  |  | Family history of other endocrine, nutritional and metabolic diseases (Z83.4, %) | 0.354 | 0.198 | 0.0299 | 0.363 | 0 | 0.0854 |
|  |  |  | Nicotine dependence (F17, %) | 0 | 0,198 | 0.0630 | 0 | 0 | - |
|  |  |  | Family history of arthritis and other diseases of the musculoskeletal sytem and connective tissue (Z82.6, %) | 0 | 0 | - | 0 | 0 | - |
|  |  | **Male** | Number of participants | 2,949 | 7,293 | - | 2,654 | 2,654 | - |
|  |  |  | Follow-up (days) median (interquartile range) | 5,187.5 (1,357) | 5,041 (1,342) | - | 5,189.5 (1.357) | 5,027.5 (1,341) | - |
|  |  |  | Age at Index (years, SD) | 0.703 ± 0.777 | 0.584 ± 0.743 | 0.1563 | 0.702 ± 0.776 | 0.702 ± 0.777 | 0.0005 |
|  |  |  | Female (%) | 0 | 0 | - | 0 | 0 | - |
|  |  |  | Male (%) | 100 | 100 | - | 100 | 100 | - |
|  |  |  | Black or African American | 41.385 | 26.996 | 0.3069 | 41.334 | 41.409 | 0.0015 |
|  |  |  | White (%) | 36.983 | 47.133 | 0.2067 | 37.001 | 37.001 | <0.0001 |
|  |  |  | Hispanic or Latino (%) | 15.275 | 15.3 | 0.0007 | 15.222 | 15.185 | 0.0010 |
|  |  |  | Persons with potential health hazards related to socioeconomic and psychosocial circumstances (Z55-Z66, %) | 0.715 | 0.669 | 0.0055 | 0.716 | 0.716 | <0.0001 |
|  |  |  | Overweight and obesity (E66, %) | 0.489 | 0.259 | 0.0378 | 0.414 | 0.377 | 0.0060 |
|  |  |  | Family history of other endocrine, nutritional and metabolic diseases (Z83.4, %) | 0 | 0.152 | 0.0552 | 0 | 0 | - |
|  |  |  | Nicotine dependence (F17, %) | 0.376 | 0.152 | 0.0437 | 0.377 | 0 | 0.0870 |
|  |  |  | Family history of arthritis and other diseases of the musculoskeletal sytem and connective tissue (Z82.6, %) | 0.376 | 0.152 | 0.0437 | 0 | 0 | - |
|  |  | **Black or African American** | Number of participants | 2,739 | 3,758 | - | 2,382 | 2,382 | - |
|  |  |  | Follow-up (days) median (interquartile range) | 5,280 (1,382) | 5,155.5 (1,455) | - | 5,293.5 (1,369) | 5,158.5 (1,493) | - |
|  |  |  | Age at Index (years, SD) | 0.729 ± 0.803 | 0.581 ± 0.753 | 0.1909 | 0.692 ± 0.785 | 0.691 ± 0.784 | 0.0016 |
|  |  |  | Female (%) | 55.193 | 47.047 | 0.1635 | 53.904 | 53.946 | 0.0008 |
|  |  |  | Male (%) | 44.807 | 52.953 | 0.1635 | 46.096 | 46.054 | 0.0008 |
|  |  |  | Black or African American | 100 | 100 | - | 100 | 100 | - |
|  |  |  | White (%) | 0 | 0 | - | 0 | 0 | - |
|  |  |  | Hispanic or Latino (%) | 2.525 | 3.222 | 0.0417 | 2.603 | 2.519 | 0.0053 |
|  |  |  | Persons with potential health hazards related to socioeconomic and psychosocial circumstances (Z55-Z66, %) | 0.896 | 1.104 | 0.0209 | 0.882 | 0.882 | <0.0001 |
|  |  |  | Overweight and obesity (E66, %) | 0.733 | 0.567 | 0.0207 | 0.672 | 0.63 | 0.0052 |
|  |  |  | Family history of other endocrine, nutritional and metabolic diseases (Z83.4, %) | 0 | 0.298 | 0.0774 | 0 | 0 | - |
|  |  |  | Nicotine dependence (F17, %) | 0.407 | 0.298 | 0.0184 | 0 | 0 | - |
|  |  |  | Family history of arthritis and other diseases of the musculoskeletal sytem and connective tissue (Z82.6, %) | 0 | 0 | - | 0 | 0 | - |
|  |  | **Hispanic** | Number of participants | 940 | 1,905 | - | 852 | 852 | - |
|  |  |  | Follow-up (days) median (interquartile range) | 5,194 (1,395) | 4,978 (1,327) | - | 5,199 (1,387.5) | 4,886 (1,294.5) | - |
|  |  |  | Age at Index (years, SD) | 0.84 ± 0.807 | 0.707 ± 0.787 | 0.1660 | 0.829 ± 0.803 | 0.831 ± 0.805 | 0.0029 |
|  |  |  | Female (%) | 52.846 | 45.567 | 0.1460 | 52.347 | 52.582 | 0.0047 |
|  |  |  | Male (%) | 47.154 | 54.433 | 0.1460 | 47.653 | 47.418 | 0.0047 |
|  |  |  | Black or African American | 7.201 | 5.674 | 0.0623 | 7.277 | 7.042 | 0.0091 |
|  |  |  | White (%) | 44.483 | 47.813 | 0.0668 | 44.836 | 45.07 | 0.0047 |
|  |  |  | Hispanic or Latino (%) | 100 | 100 | - | 100 | 100 | - |
|  |  |  | Persons with potential health hazards related to socioeconomic and psychosocial circumstances (Z55-Z66, %) | 1.161 | 0.591 | 0.0612 | 1.174 | 1.174 | <0.0001 |
|  |  |  | Overweight and obesity (E66, %) | 1.161 | 0.65 | 0.0540 | 1.174 | 1.174 | <0.0001 |
|  |  |  | Family history of other endocrine, nutritional and metabolic diseases (Z83.4, %) | 1.161 | 0.591 | 0.061 | 0 | 0.0 | - |
|  |  |  | Nicotine dependence (F17, %) | 0 | 0.591 | 0.1090 | 0 | 0 | - |
|  |  |  | Family history of arthritis and other diseases of the musculoskeletal sytem and connective tissue (Z82.6, %) | 0 | 0.051 | 0.0319 | 0 | 0 | - |
|  |  | **White** | Number of participants | 2,093 | 5,831 | - | 1,873 | 1,873 | - |
|  |  |  | Follow-up (days) median (interquartile range) | 5,181 (1,441) | 5,125 (1,395) | - | 5,181 (1,439) | 5,084 (1,450) | - |
|  |  |  | Age at Index (years, SD) | 0.835 ± 0.791 | 0.666 ± 0.763 | 0.2180 | 0.835 ± 0.791 | 0.836 ± 0.791 | 0.0013 |
|  |  |  | Female (%) | 47.545 | 43.9 | 0.0732 | 47.571 | 47.571 | <0.0001 |
|  |  |  | Male (%) | 52.455 | 56.081 | 0.0728 | 52.429 | 52.429 | <0.0001 |
|  |  |  | Black or African American | 0 | 0 | - | 0 | 0 | - |
|  |  |  | White (%) | 100 | 100 | - | 100 | 100 | - |
|  |  |  | Hispanic or Latino (%) | 20.438 | 15.618 | 0.1256 | 20.448 | 20.342 | 0.0027 |
|  |  |  | Persons with potential health hazards related to socioeconomic and psychosocial circumstances (Z55-Z66, %) | 0.534 | 0.56 | 0.0036 | 0.534 | 0.534 | <0.0001 |
|  |  |  | Overweight and obesity (E66, %) | 0.534 | 0.386 | 0.0218 | 0.534 | 0.534 | <0.0001 |
|  |  |  | Family history of other endocrine, nutritional and metabolic diseases (Z83.4, %) | 0.534 | 0.193 | 0.0566 | 0.534 | 0 | 0.1036 |
|  |  |  | Nicotine dependence (F17, %) | 0.534 | 0.193 | 0.0566 | 0.534 | 0.534 | <0.0001 |
|  |  |  | Family history of arthritis and other diseases of the musculoskeletal sytem and connective tissue (Z82.6, %) | 0.534 | 0.193 | 0.0566 | 0 | 0 | - |
| **Persistent (cases) versus no AD (controls)** | **Autoimmune diseases** | **Primary** | Number of participants | 6,738 | 188,340 | - | 4,867 | 4,867 | - |
|  |  |  | Follow-up (days) median (interquartile range) | 5,302.5 (1,427.5) | 4,844 (1,145) | - | 5,301 (1,427) | 4,770 (1,149) | - |
|  |  |  | Age at Index (years, SD) | 0.753 ± 0.798 | 0.585 ± 0.774 | 0.2118 | 0.752 ± 0.798 | 0.753 ± 0.798 | 0.0005 |
|  |  |  | Female (%) | 51.537 | 48.934 | 0.0521 | 51.531 | 51.531 | <0.0001 |
|  |  |  | Male (%) | 48.361 | 51.025 | 0.0533 | 48.367 | 48.428 | 0.0012 |
|  |  |  | Black or African American | 43.422 | 21.498 | 0.4816 | 43.353 | 43.374 | 0.0004 |
|  |  |  | White (%) | 35.02 | 55.618 | 0.4229 | 35.073 | 35.073 | <0.0001 |
|  |  |  | Hispanic or Latino (%) | 16.967 | 20.854 | 0.0994 | 16.951 | 16.93 | 0.0005 |
|  |  |  | Persons with potential health hazards related to socioeconomic and psychosocial circumstances (Z55-Z66, %) | 0.656 | 0.287 | 0.0538 | 0.657 | 0.657 | <0.0001 |
|  |  |  | Overweight and obesity (E66, %) | 0.635 | 0.047 | 0.1010 | 0.39 | 0.411 | 0.0033 |
|  |  |  | Family history of other endocrine, nutritional and metabolic diseases (Z83.4, %) | 0.205 | 0.017 | 0.0566 | 0.205 | 0.205 | <0.0001 |
|  |  |  | Nicotine dependence (F17, %) | 0.205 | 0.007 | 0.0610 | 0.205 | 0 | 0.0642 |
|  |  |  | Family history of arthritis and other diseases of the musculoskeletal sytem and connective tissue (Z82.6, %) | 0.205 | 0.007 | 0.0610 | 0 | 0 | - |
|  |  | **S1** | Number of participants | 7,402 | 133,049 | - | 5,485 | 5,485 | - |
|  |  |  | Follow-up (days) median (interquartile range) | 5,223 (1,401) | 5,095 (1,225) | - | 5,222 (1,399) | 4,934 (1,289) | - |
|  |  |  | Age at Index (years, SD) | 0.768 ± 0.801 | 0.536 ± 0.774 | 0.2953 | 0.767 ± 0.801 | 0.768 ± 0.801 | 0.0007 |
|  |  |  | Female (%) | 51.937 | 49.065 | 0.0575 | 51.905 | 51.869 | 0.0007 |
|  |  |  | Male (%) | 47.972 | 50.885 | 0.0583 | 48.004 | 48.077 | 0.0015 |
|  |  |  | Black or African American | 44.572 | 19.68 | 0.5531 | 44.467 | 44.521 | 0.0011 |
|  |  |  | White (%) | 33.752 | 58.32 | 0.5086 | 33.82 | 33.801 | 0.0004 |
|  |  |  | Hispanic or Latino (%) | 15.476 | 10.897 | 0.1356 | 15.497 | 15.46 | 0.0010 |
|  |  |  | Persons with potential health hazards related to socioeconomic and psychosocial circumstances (Z55-Z66, %) | 0.582 | 0.235 | 0.0543 | 0.583 | 0.565 | 0.0024 |
|  |  |  | Overweight and obesity (E66, %) | 0.582 | 0.041 | 0.0972 | 0.346 | 0.365 | 0.0031 |
|  |  |  | Family history of other endocrine, nutritional and metabolic diseases (Z83.4, %) | 0.182 | 0.01 | 0.0556 | 0.182 | 0 | 0.0604 |
|  |  |  | Nicotine dependence (F17, %) | 0.182 | 0.01 | 0.0556 | 0.182 | 0 | 0.0604 |
|  |  |  | Family history of arthritis and other diseases of the musculoskeletal sytem and connective tissue (Z82.6, %) | 0.182 | 0.01 | 0.0556 | 0 | 0 | - |
|  |  | **S2** | Number of participants | 6,150 | 112,077 | - | 5,488 | 5,488 | - |
|  |  |  | Follow-up (days) median (interquartile range) | 5,223 (1,403) | 5,095 (1,227) | - | 5,223 (1,400) | 4,962 (1,303.5) | - |
|  |  |  | Age at Index (years, SD) | 0.769 ± 0.801 | 0.537 ± 0.775 | 0.2947 | 0.768 ± 0.801 | 0.769 ± 0.802 | 0.0002 |
|  |  |  | Female (%) | 51.935 | 49.067 | 0.0574 | 51.913 | 51.877 | 0.0007 |
|  |  |  | Male (%) | 47.974 | 50.883 | 0.0582 | 47.996 | 48.05 | 0.0011 |
|  |  |  | Black or African American | 44.558 | 19.695 | 0.5524 | 44.461 | 44.515 | 0.0011 |
|  |  |  | White (%) | 33.763 | 58.308 | 0.5081 | 33.819 | 33.819 | <0.0001 |
|  |  |  | Hispanic or Latino (%) | 15.482 | 10.901 | 0.1357 | 15.507 | 15.434 | 0.0020 |
|  |  |  | Persons with potential health hazards related to socioeconomic and psychosocial circumstances (Z55-Z66, %) | 0.582 | 0.236 | 0.0541 | 0.583 | 0.583 | <0.0001 |
|  |  |  | Overweight and obesity (E66, %) | 0.582 | 0.042 | 0.0969 | 0.346 | 0.346 | <0.0001 |
|  |  |  | Family history of other endocrine, nutritional and metabolic diseases (Z83.4, %) | 0.182 | 0.01 | 0.0556 | 0.182 | 0 | 0.0604 |
|  |  |  | Nicotine dependence (F17, %) | 0.182 | 0.01 | 0.0556 | 0 | 0 | - |
|  |  |  | Family history of arthritis and other diseases of the musculoskeletal sytem and connective tissue (Z82.6, %) | 0.182 | 0.01 | 0.0556 | 0 | 0 | - |
|  |  | **Female** | Number of participants | 3,212 | 55,076 | - | 2.846 | 2,846 | - |
|  |  |  | Follow-up (days) median (interquartile range) | 5,249.5 (1,454) | 5,117 (1,255) | - | 5,245.5 (1,454) | 4,964 (1,322) | - |
|  |  |  | Age at Index (years, SD) | 0.833 ± 0.818 | 0.539 ± 0.776 | 0.3686 | 0.831 ± 0.818 | 0.832 ± 0.819 | 0.0004 |
|  |  |  | Female (%) | 100 | 100 | - | 100 | 100 | - |
|  |  |  | Male (%) | 0 | 0 | - | 0 | 0 | - |
|  |  |  | Black or African American | 47.376 | 19.752 | 0.6117 | 47.259 | 47.259 | <0.0001 |
|  |  |  | White (%) | 30.931 | 58.355 | 0.5735 | 30.991 | 31.026 | 0.0008 |
|  |  |  | Hispanic or Latino (%) | 15.78 | 10.881 | 0.1445 | 15.741 | 15.741 | <0.0001 |
|  |  |  | Persons with potential health hazards related to socioeconomic and psychosocial circumstances (Z55-Z66, %) | 0.455 | 0.215 | 0.0416 | 0.422 | 0.422 | <0.0001 |
|  |  |  | Overweight and obesity (E66, %) | 0.665 | 0.038 | 0.1061 | 0.351 | 0.351 | <0.0001 |
|  |  |  | Family history of other endocrine, nutritional and metabolic diseases (Z83.4, %) | 0.35 | 0.02 | 0.0769 | 0 | 0 | - |
|  |  |  | Nicotine dependence (F17, %) | 0 | 0.02 | 0.0199 | 0 | 0 | - |
|  |  |  | Family history of arthritis and other diseases of the musculoskeletal sytem and connective tissue (Z82.6, %) | 0 | 0.02 | 0.0199 | 0 | 0 | - |
|  |  | **Male** | Number of participants | 2,933 | 56,949 | - | 2,634 | 2,634 | - |
|  |  |  | Follow-up (days) median (interquartile range) | 5,192.5 (1,355) | 5,077 (1,206) | - | 5,190.5 (1.354) | 4,914.5 (1,262) | - |
|  |  |  | Age at Index (years, SD) | 0.701 ± 0.777 | 0.534 ± 0.773 | 0.2150 | 0.701 ± 0.778 | 0.701 ± 0.778 | 0.0005 |
|  |  |  | Female (%) | 0 | 0 | - | 0 | 0 | - |
|  |  |  | Male (%) | 100 | 100 | - | 100 | 100 | - |
|  |  |  | Black or African American | 41.591 | 19.64 | 0.4904 | 41.572 | 41.572 | <0.0001 |
|  |  |  | White (%) | 36.894 | 58.291 | 0.4386 | 36.902 | 36.902 | <0.0001 |
|  |  |  | Hispanic or Latino (%) | 15.189 | 10.93 | 0.1267 | 15.186 | 15.148 | 0.0011 |
|  |  |  | Persons with potential health hazards related to socioeconomic and psychosocial circumstances (Z55-Z66, %) | 0.72 | 0.257 | 0.0664 | 0.721 | 0.683 | 0.0045 |
|  |  |  | Overweight and obesity (E66, %) | 0.492 | 0.046 | 0.0862 | 0.38 | 0.38 | <0.0001 |
|  |  |  | Family history of other endocrine, nutritional and metabolic diseases (Z83.4, %) | 0 | 0.019 | 0.0196 | 0 | 0 | - |
|  |  |  | Nicotine dependence (F17, %) | 0.379 | 0 | 0.0872 | 0 | 0 | - |
|  |  |  | Family history of arthritis and other diseases of the musculoskeletal sytem and connective tissue (Z82.6, %) | 0.379 | 0.019 | 0.0808 | 0 | 0 | - |
|  |  | **Black or African American** | Number of participants | 2,737 | 23,248 | - | 2,440 | 2,440 | - |
|  |  |  | Follow-up (days) median (interquartile range) | 5,280 (1,381) | 4,990 (1,404) | - | 5,278 (1,380) | 4,882 (1,319) | - |
|  |  |  | Age at Index (years, SD) | 0.729 ± 0.803 | 0.536 ± 0.789 | 0.2429 | 0.727 ± 0.803 | 0.727 ± 0.803 | <0.0001 |
|  |  |  | Female (%) | 55.22 | 49.207 | 0.1206 | 55.123 | 55.123 | <0.0001 |
|  |  |  | Male (%) | 44.78 | 50.738 | 0.1195 | 44.877 | 44.877 | <0.0001 |
|  |  |  | Black or African American | 100 | 100 | - | 100 | 100 | - |
|  |  |  | White (%) | 0 | 0 | - | 0 | 0 | - |
|  |  |  | Hispanic or Latino (%) | 2.488 | 3.205 | 0.0432 | 2.5 | 2.5 | <0.0001 |
|  |  |  | Persons with potential health hazards related to socioeconomic and psychosocial circumstances (Z55-Z66, %) | 0.897 | 0.431 | 0.0574 | 0.902 | 0.902 | <0.0001 |
|  |  |  | Overweight and obesity (E66, %) | 0.734 | 0.054 | 0.1086 | 0.41 | 0.41 | <0.0001 |
|  |  |  | Family history of other endocrine, nutritional and metabolic diseases (Z83.4, %) | 0 | 0.05 | 0.0315 | 0 | 0 | - |
|  |  |  | Nicotine dependence (F17, %) | 0.408 | 0 | 0.0905 | 0 | 0 | - |
|  |  |  | Family history of arthritis and other diseases of the musculoskeletal sytem and connective tissue (Z82.6, %) | 0 | 0.05 | 0.315 | 0 | 0 | - |
|  |  | **Hispanic** | Number of participants | 931 | 12,317 | - | 848 | 848 | - |
|  |  |  | Follow-up (days) median (interquartile range) | 5,199 (1,398.5) | 4,977 (1,257) | - | 5,196 (1,402.5) | 5,005 (1,221.5) | - |
|  |  |  | Age at Index (years, SD) | 0.843 ± 0.806 | 0.611 ± 0.8 | 0.2887 | 0.842 ± 0.808 | 0.844 ± 0.808 | 0.0029 |
|  |  |  | Female (%) | 52.934 | 48.975 | 0.0793 | 52.83 | 52.712 | 0.0024 |
|  |  |  | Male (%) | 47.066 | 51.016 | 0.0791 | 47.17 | 47.288 | 0.0024 |
|  |  |  | Black or African American | 7.16 | 5,791 | 0.0566 | 7.193 | 7.311 | 0.0045 |
|  |  |  | White (%) | 44.366 | 33.116 | 0.2325 | 44.458 | 44.34 | 0.0024 |
|  |  |  | Hispanic or Latino (%) | 100 | 100 | - | 100 | 100 | - |
|  |  |  | Persons with potential health hazards related to socioeconomic and psychosocial circumstances (Z55-Z66, %) | 1.174 | 0.107 | 0.1340 | 1.179 | 1.179 | <0.0001 |
|  |  |  | Overweight and obesity (E66, %) | 1.174 | 0.098 | 0.1356 | 1.179 | 1.179 | <0.0001 |
|  |  |  | Family history of other endocrine, nutritional and metabolic diseases (Z83.4, %) | 1.174 | 0.09 | 0.1372 | 0 | 0 | - |
|  |  |  | Nicotine dependence (F17, %) | 0 | 0 | - | 0 | 0 | - |
|  |  |  | Family history of arthritis and other diseases of the musculoskeletal sytem and connective tissue (Z82.6, %) | 0 | 0 | - | 0 | 0 | - |
|  |  | **White** | Number of participants | 2,079 | 64,282 | - | 1,855 | 1,855 | - |
|  |  |  | Follow-up (days) median (interquartile range) | 5,181 (1,444) | 5,140.5 (1,143) | - | 5,181 (1,444) | 5,066 (1,161) | - |
|  |  |  | Age at Index (years, SD) | 0.835 ± 0.791 | 0.527 ± 0.763 | 0.3963 | 0.836 ± 0.791 | 0.837 ± 0.792 | 0.0014 |
|  |  |  | Female (%) | 47.578 | 49.09 | 0.0303 | 47.601 | 47.601 | <0.0001 |
|  |  |  | Male (%) | 52.422 | 50.868 | 0.0311 | 52.399 | 52.399 | <0.0001 |
|  |  |  | Black or African American | 0 | 0 | - | 0 | 0 | - |
|  |  |  | White (%) | 100 | 100 | - | 100 | 100 | - |
|  |  |  | Hispanic or Latino (%) | 20.344 | 6.191 | 0.4266 | 20.323 | 20.27 | 0.0013 |
|  |  |  | Persons with potential health hazards related to socioeconomic and psychosocial circumstances (Z55-Z66, %) | 0.538 | 0.187 | 0.0584 | 0.539 | 0.539 | <0.0001 |
|  |  |  | Overweight and obesity (E66, %) | 0.538 | 0.025 | 0.0969 | 0.539 | 0.539 | <0.0001 |
|  |  |  | Family history of other endocrine, nutritional and metabolic diseases (Z83.4, %) | 0.538 | 0.017 | 0.0993 | 0 | 0 | - |
|  |  |  | Nicotine dependence (F17, %) | 0.538 | 0.017 | 0.0993 | 0 | 0 | - |
|  |  |  | Family history of arthritis and other diseases of the musculoskeletal sytem and connective tissue (Z82.6, %) | 0.538 | 0.017 | 0.0993 | 0 | 0 | - |
| **Transient (cases) versus no AD (controls)** | **Autoimmune diseases** | **Primary** | Number of participants | 25,313 | 188,323 | - | 18,099 | 18,099 | - |
|  |  |  | Follow-up (days) median (interquartile range) | 4,949.5 (1,302.5) | 4,836 (1,153) | - | 4,949 (1,303) | 4,778 (1,151) | - |
|  |  |  | Age at Index (years, SD) | 0.661 ± 0.773 | 0.585 ± 0.791 | 0.0962 | 0.66 ± 0.773 | 0.66 ± 0.773 | <0.0001 |
|  |  |  | Female (%) | 44.665 | 48.925 | 0.0855 | 44.638 | 44.621 | 0.0003 |
|  |  |  | Male (%) | 55.33 | 51.034 | 0.0862 | 55.357 | 55.329 | 0.0006 |
|  |  |  | Black or African American | 31.493 | 22.554 | 0.2023 | 31.422 | 31.422 | <0.0001 |
|  |  |  | White (%) | 42.444 | 53.415 | 0.2209 | 42.477 | 42.472 | 0.0001 |
|  |  |  | Hispanic or Latino (%) | 18.32 | 21.755 | 0.0859 | 18.294 | 18.272 | 0.0006 |
|  |  |  | Persons with potential health hazards related to socioeconomic and psychosocial circumstances (Z55-Z66, %) | 0.755 | 0.298 | 0.0631 | 0.729 | 0.746 | 0.0019 |
|  |  |  | Overweight and obesity (E66, %) | 0.468 | 0.053 | 0.0817 | 0.265 | 0.254 | 0.0022 |
|  |  |  | Family history of other endocrine, nutritional and metabolic diseases (Z83.4, %) | 0.055 | 0.017 | 0.0203 | 0.055 | 0.055 | <0.0001 |
|  |  |  | Nicotine dependence (F17, %) | 0.055 | 0.007 | 0.0276 | 0.055 | 0.055 | <0.0001 |
|  |  |  | Family history of arthritis and other diseases of the musculoskeletal sytem and connective tissue (Z82.6, %) | 0.055 | 0.007 | 0.0271 | 0.055 | 0.055 | <0.0001 |
|  |  | **S1** | Number of participants | 18,452 | 148,738 | - | 12,072 | 12,072 | - |
|  |  |  | Follow-up (days) median (interquartile range) | 5,058.5 (1,393) | 4,976 (1,124.5) | - | 5,058.5 (1,393) | 4,949 (1,156.5) | - |
|  |  |  | Age at Index (years, SD) | 0.629 ± 0.762 | 0.514 ± 0.767 | 0.1503 | 0.628 ± 0.762 | 0.628 ± 0.761 | 0.0002 |
|  |  |  | Female (%) | 45.216 | 49.111 | 0.0781 | 45.154 | 45.154 | <0.0001 |
|  |  |  | Male (%) | 54.776 | 50.882 | 0.0781 | 54.838 | 54.838 | <0.0001 |
|  |  |  | Black or African American | 28.218 | 21.589 | 0.1538 | 28.173 | 28.181 | 0.0002 |
|  |  |  | White (%) | 48.744 | 60.001 | 0.2275 | 48.791 | 48.791 | <0.0001 |
|  |  |  | Hispanic or Latino (%) | 14.708 | 18.005 | 0.0892 | 14.695 | 14.695 | <0.0001 |
|  |  |  | Persons with potential health hazards related to socioeconomic and psychosocial circumstances (Z55-Z66, %) | 0.669 | 0.247 | 0.0626 | 0.663 | 0.671 | 0.0010 |
|  |  |  | Overweight and obesity (E66, %) | 0.454 | 0.037 | 0.0845 | 0.257 | 0.257 | <0.0001 |
|  |  |  | Family history of other endocrine, nutritional and metabolic diseases (Z83.4, %) | 0.083 | 0.012 | 0.0325 | 0.083 | 0.083 | <0.0001 |
|  |  |  | Nicotine dependence (F17, %) | 0.083 | 0.009 | 0.0347 | 0.083 | 0.083 | <0.0001 |
|  |  |  | Family history of arthritis and other diseases of the musculoskeletal sytem and connective tissue (Z82.6, %) | 0.083 | 0.009 | 0.0347 | 0.083 | 0 | 0.0407 |
|  |  | **S2** | Number of participants | 13,470 | 127,391 | - | 12,094 | 12,094 | - |
|  |  |  | Follow-up (days) median (interquartile range) | 5,057.5 (1,395) | 4,976 (1,126) | - | 5,058 (1,395) | 4,932 (1,174) | - |
|  |  |  | Age at Index (years, SD) | 0.631 ± 0.763 | 0.516 ± 0.768 | 0.1500 | 0.63 ± 0.763 | 0.63 ± 0.763 | 0.0002 |
|  |  |  | Female (%) | 45.224 | 49.114 | 0.0780 | 45.163 | 45.155 | 0.0002 |
|  |  |  | Male (%) | 54.768 | 50.879 | 0.0780 | 54.829 | 54.829 | <0.0001 |
|  |  |  | Black or African American | 28.221 | 21.568 | 0.1539 | 28.163 | 28.179 | 0.0004 |
|  |  |  | White (%) | 48.705 | 59.969 | 0.2276 | 48.751 | 48.735 | 0.0003 |
|  |  |  | Hispanic or Latino (%) | 14.684 | 17.999 | 0.0897 | 14.668 | 14.668 | <0.0001 |
|  |  |  | Persons with potential health hazards related to socioeconomic and psychosocial circumstances (Z55-Z66, %) | 0.668 | 0.248 | 0.0623 | 0.661 | 0.67 | 0.0010 |
|  |  |  | Overweight and obesity (E66, %) | 0.462 | 0.039 | 0.0847 | 0.281 | 0.273 | 0.0016 |
|  |  |  | Family history of other endocrine, nutritional and metabolic diseases (Z83.4, %) | 0.082 | 0.012 | 0.0325 | 0.083 | 0.083 | <0.0001 |
|  |  |  | Nicotine dependence (F17, %) | 0.082 | 0.009 | 0.0347 | 0.083 | 0.083 | <0.0001 |
|  |  |  | Family history of arthritis and other diseases of the musculoskeletal sytem and connective tissue (Z82.6, %) | 0.082 | 0.009 | 0.0347 | 0.083 | 0 | 0.0407 |
|  |  | **Female** | Number of participants | 5,516 | 58,170 | - | 4,941 | 4,941 | - |
|  |  |  | Follow-up (days) median (interquartile range) | 5,082 (1,452) | 5,192 (1,334) | - | 5,083 (1,458) | 5,125 (1,418) | - |
|  |  |  | Age at Index (years, SD) | 0.688 ± 0.781 | 0.497 ± 0.756 | 0.2483 | 0.685 ± 0.779 | 0.685 ± 0.78 | 0.0003 |
|  |  |  | Female (%) | 100 | 100 | - | 100 | 100 | - |
|  |  |  | Male (%) | 0 | 0 | - | 0 | 0 | - |
|  |  |  | Black or African American | 28.258 | 17.72 | 0.2524 | 28.253 | 28.274 | 0.0004 |
|  |  |  | White (%) | 46.928 | 62.05 | 0.3072 | 46.934 | 46.914 | 0.0004 |
|  |  |  | Hispanic or Latino (%) | 10.393 | 9.991 | 0.0133 | 10.342 | 10.342 | <0.0001 |
|  |  |  | Persons with potential health hazards related to socioeconomic and psychosocial circumstances (Z55-Z66, %) | 0.665 | 0.19 | 0.0729 | 0.668 | 0.648 | 0.0025 |
|  |  |  | Overweight and obesity (E66, %) | 0.745 | 0.034 | 0.1143 | 0.324 | 0.324 | <0.0001 |
|  |  |  | Family history of other endocrine, nutritional and metabolic diseases (Z83.4, %) | 0.201 | 0.019 | 0.0550 | 0.202 | 0.202 | <0.0001 |
|  |  |  | Nicotine dependence (F17, %) | 0.201 | 0.019 | 0.0550 | 0 | 0 | - |
|  |  |  | Family history of arthritis and other diseases of the musculoskeletal sytem and connective tissue (Z82.6, %) | 0 | 0.019 | 0.0195 | 0 | 0 | - |
|  |  | **Male** | Number of participants | 6,819 | 60,093 | - | 6,169 | 6,169 | - |
|  |  |  | Follow-up (days) median (interquartile range) | 5,073 (1,374) | 5,163 (1,279) | - | 5,073 (1.374) | 5,115 (1,308) | - |
|  |  |  | Age at Index (years, SD) | 0.576 ± 0.738 | 0.495 ± 0.753 | 0.1083 | 0.576 ± 0.739 | 0.576 ± 0.739 | <0.0001 |
|  |  |  | Female (%) | 0 | 0 | - | 0 | 0 | - |
|  |  |  | Male (%) | 100 | 100 | - | 100 | 100 | - |
|  |  |  | Black or African American | 25.659 | 17.678 | 0.1946 | 25.612 | 25.644 | 0.0007 |
|  |  |  | White (%) | 50.866 | 61.971 | 0.2254 | 50.916 | 50.883 | 0.0006 |
|  |  |  | Hispanic or Latino (%) | 10.225 | 9.966 | 0.0086 | 10.18 | 10.196 | 0.0005 |
|  |  |  | Persons with potential health hazards related to socioeconomic and psychosocial circumstances (Z55-Z66, %) | 0.663 | 0.232 | 0.0646 | 0.616 | 0.616 | <0.0001 |
|  |  |  | Overweight and obesity (E66, %) | 0.259 | 0.044 | 0.0553 | 0.162 | 0.162 | <0.0001 |
|  |  |  | Family history of other endocrine, nutritional and metabolic diseases (Z83.4, %) | 0.162 | 0.018 | 0.0479 | 0.162 | 0 | 0.0570 |
|  |  |  | Nicotine dependence (F17, %) | 0.162 | 0.018 | 0.0479 | 0.162 | 0.162 | <0.0001 |
|  |  |  | Family history of arthritis and other diseases of the musculoskeletal sytem and connective tissue (Z82.6, %) | 0.162 | 0.018 | 0.0479 | 0.162 | 0 | 0.0570 |
|  |  | **Black or African American** | Number of participants | 3,280 | 21,658 | - | 2,945 | 2,945 | - |
|  |  |  | Follow-up (days) median (interquartile range) | 5,116 (1,487) | 5,105 (1,469) | - | 5,115 (1,487) | 5,054 (1,455) | - |
|  |  |  | Age at Index (years, SD) | 0.578 ± 0.754 | 0.49 ± 0.768 | 0.1155 | 0.576 ± 0.753 | 0.576 ± 0.753 | 0.0005 |
|  |  |  | Female (%) | 46.739 | 49.148 | 0.0482 | 46.723 | 46.723 | <0.0001 |
|  |  |  | Male (%) | 53.261 | 50.847 | 0.0483 | 53.277 | 53.277 | <0.0001 |
|  |  |  | Black or African American | 100 | 100 | - | 100 | 100 | - |
|  |  |  | White (%) | 0 | 0 | - | 0 | 0 | - |
|  |  |  | Hispanic or Latino (%) | 3.143 | 3.478 | 0.0187 | 3.09 | 3.124 | 0.0020 |
|  |  |  | Persons with potential health hazards related to socioeconomic and psychosocial circumstances (Z55-Z66, %) | 1.149 | 0.417 | 0.0832 | 1.121 | 1.121 | <0.0001 |
|  |  |  | Overweight and obesity (E66, %) | 0.541 | 0.053 | 0.0869 | 0.34 | 0.34 | <0.0001 |
|  |  |  | Family history of other endocrine, nutritional and metabolic diseases (Z83.4, %) | 0.338 | 0.053 | 0.0644 | 0 | 0 | - |
|  |  |  | Nicotine dependence (F17, %) | 0.338 | 0 | 0.0824 | 0 | 0 | - |
|  |  |  | Family history of arthritis and other diseases of the musculoskeletal sytem and connective tissue (Z82.6, %) | 0 | 0.053 | 0.0327 | 0 | 0 | - |
|  |  | **Hispanic** | Number of participants | 1,295 | 11,862 | - | 1,161 | 1,161 | - |
|  |  |  | Follow-up (days) median (interquartile range) | 4,874.5 (1,366) | 5,014.5 (1,345) | - | 4,878 (1,363) | 4,964 (1,293) | - |
|  |  |  | Age at Index (years, SD) | 0.724 ± 0.784 | 0.589 ± 0.79 | 0.1717 | 0.719 ± 0.783 | 0.719 ± 0.783 | <0.0001 |
|  |  |  | Female (%) | 45.043 | 49.179 | 0.0829 | 45.047 | 45.875 | 0.0035 |
|  |  |  | Male (%) | 54.957 | 50.821 | 0.0829 | 54.953 | 55.125 | 0.0035 |
|  |  |  | Black or African American | 7.949 | 6.074 | 0.0735 | 7.838 | 7.838 | <0.0001 |
|  |  |  | White (%) | 31.282 | 36.257 | 0.1053 | 31.008 | 31.266 | 0.0056 |
|  |  |  | Hispanic or Latino (%) | 100 | 100 | - | 100 | 100 | - |
|  |  |  | Persons with potential health hazards related to socioeconomic and psychosocial circumstances (Z55-Z66, %) | 0.855 | 0.103 | 0.1091 | 0.861 | 0.861 | <0.0001 |
|  |  |  | Overweight and obesity (E66, %) | 0.94 | 0.093 | 0.1183 | 0.861 | 0.861 | <0.0001 |
|  |  |  | Family history of other endocrine, nutritional and metabolic diseases (Z83.4, %) | 0.855 | 0.093 | 0.1110 | 0 | 0.861 | 0.1318 |
|  |  |  | Nicotine dependence (F17, %) | 0.855 | 0 | 0.1313 | 0 | 0 | - |
|  |  |  | Family history of arthritis and other diseases of the musculoskeletal sytem and connective tissue (Z82.6, %) | 0 | 0 | - | 0 | 0 | - |
|  |  | **White** | Number of participants | 6,180 | 72,485 | - | 5,537 | 5,537 | - |
|  |  |  | Follow-up (days) median (interquartile range) | 5,109 (1,401) | 5,223 (1,236) | - | 5,113 (1,399) | 5,223 (1,280) | - |
|  |  |  | Age at Index (years, SD) | 0.667 ± 0.763 | 0.481 ± 0.74 | 0.2475 | 0.666 ± 0.762 | 0.666 ± 0.762 | <0.0001 |
|  |  |  | Female (%) | 42.618 | 49.147 | 0.1313 | 42.532 | 42.532 | <0.0001 |
|  |  |  | Male (%) | 57.364 | 50.848 | 0.1310 | 57.45 | 57.468 | 0.0004 |
|  |  |  | Black or African American | 0 | 0 | - | 0 | 0 | - |
|  |  |  | White (%) | 100 | 100 | - | 100 | 100 | - |
|  |  |  | Hispanic or Latino (%) | 6.59 | 5.834 | 0.0313 | 6.52 | 6.52 | <0.0001 |
|  |  |  | Persons with potential health hazards related to socioeconomic and psychosocial circumstances (Z55-Z66, %) | 0.486 | 0.168 | 0.0557 | 0.47 | 0.47 | <0.0001 |
|  |  |  | Overweight and obesity (E66, %) | 0.324 | 0.024 | 0.0720 | 0.181 | 0.181 | <0.0001 |
|  |  |  | Family history of other endocrine, nutritional and metabolic diseases (Z83.4, %) | 0.18 | 0.015 | 0.0529 | 0.181 | 0.181 | <0.0001 |
|  |  |  | Nicotine dependence (F17, %) | 0.18 | 0.015 | 0.0529 | 0.181 | 0.181 | <0.0001 |
|  |  |  | Family history of arthritis and other diseases of the musculoskeletal sytem and connective tissue (Z82.6, %) | 0.18 | 0.015 | 0.0529 | 0 | 0 | - |

**Supplement Table 2**. Baseline characteristics before and after propensity-score matching for all analyses relating to the risk of autoimmune diseases in patients with persistent, transient or no childhood atopic dermatitis (AD).

| **Comparison** | **Outcome** | **Analysis** | **Characteristic** | **Before matching** | | | **After matching** | | |
| --- | --- | --- | --- | --- | --- | --- | --- | --- | --- |
|  |  |  |  | **Cases** | **Controls** | **Std. diff.** | **Cases** | **Controls** | **Std. diff.** |
| **Persistent (cases) versus transient AD (controls)** | **Cardiovascular diseases** | **Primary** | Number of participants | 7,168 | 25,957 | - | 5,304 | 5,304 | - |
|  |  |  | Follow-up (days) median (interquartile range) | 5,256 (1,402) | 4,926 (1,283) | - | 5,256 (1,403) | 4,948 (1,296.5) | - |
|  |  |  | Age at Index (years, SD) | 0.762 ± 0.799 | 0.665 ± 0.775 | 0.1229 | 0.762 ± 0.799 | 0.763 ± 0.8 | 0.0014 |
|  |  |  | Female (%) | 51.857 | 44.614 | 0.1453 | 51.867 | 51.81 | 0.0011 |
|  |  |  | Male (%) | 48.124 | 55.38 | 0.1456 | 48.133 | 48.19 | 0.0011 |
|  |  |  | Black or African American | 43.695 | 30.664 | 0.2721 | 43.703 | 43.759 | 0.0004 |
|  |  |  | White (%) | 34.27 | 43.64 | 0.1930 | 34.276 | 34.238 | 0.0008 |
|  |  |  | Hispanic or Latino (%) | 17.286 | 18.024 | 0.0194 | 17.289 | 17.213 | 0.0020 |
|  |  |  | Persons with potential health hazards related to socioeconomic and psychosocial circumstances (Z55-Z66, %) | 0.641 | 0.74 | 0.0120 | 0.641 | 0.66 | 0.0023 |
|  |  |  | Overweight and obesity (E66, %) | 0.603 | 0.469 | 0.0184 | 0.603 | 0.6528 | 0.0101 |
|  |  |  | Family history of ischemic heart disease and other diseases of the circulatory system (Z82.4, %) | 0.189 | 0.197 | 0.0019 | 0.189 | 0.189 | <0.0001 |
|  |  |  | Nicotine dependence (F17, %) | 0.189 | 0.053 | 0.0389 | 0.189 | 0.189 | <0.0001 |
|  |  |  | Diabetes mellitus (E08-E13, %) | 0.189 | 0.053 | 0.0389 | 0.189 | 0.189 | <0.0001 |
|  |  |  | Disorders of lipoprotein metabolism and other lipidemias (E78, %) | 0.189 | 0.133 | 0.0138 | 0.189 | 0.189 | <0.0001 |
|  |  | **S1** | Number of participants | 7,406 | 18,040 | - | 5,448 | 5,448 | - |
|  |  |  | Follow-up (days) median (interquartile range) | 5,223 (1,402) | 4,958 (1,336) | - | 5,225.5 (1,400) | 4,914 (1,367.5) | - |
|  |  |  | Age at Index (years, SD) | 0.768 ± 0.801 | 0.651 ± 0.768 | 0.1483 | 0.758 ± 0.797 | 0.758 ± 0.795 | 0.0032 |
|  |  |  | Female (%) | 51.899 | 44.613 | 0.1462 | 51.597 | 51.523 | 0.0015 |
|  |  |  | Male (%) | 48.01 | 55.379 | 0.1479 | 48.403 | 48.477 | 0.0015 |
|  |  |  | Black or African American | 44.539 | 27.763 | 0.3564 | 44.181 | 44.64 | 0.0092 |
|  |  |  | White (%) | 33.709 | 47.357 | 0.2807 | 33.976 | 33.425 | 0.0116 |
|  |  |  | Hispanic or Latino (%) | 15.501 | 10.628 | 0.1450 | 15.565 | 15.162 | 0.0112 |
|  |  |  | Persons with potential health hazards related to socioeconomic and psychosocial circumstances (Z55-Z66, %) | 0.582 | 0.578 | 0.0004 | 0.587 | 0.0181 | 0.0181 |
|  |  |  | Overweight and obesity (E66, %) | 0.582 | 0.407 | 0.0249 | 0.587 | 0.569 | 0.0024 |
|  |  |  | Family history of ischemic heart disease and other diseases of the circulatory system (Z82.4, %) | 0.182 | 0.081 | 0.0277 | 0.184 | 0.184 | <0.0001 |
|  |  |  | Nicotine dependence (F17, %) | 0.182 | 0.081 | 0.0277 | 0.184 | 0.184 | <0.0001 |
|  |  |  | Diabetes mellitus (E08-E13, %) | 0.182 | 0.081 | 0.0277 | 0.184 | 0.184 | <0.0001 |
|  |  |  | Disorders of lipoprotein metabolism and other lipidemias (E78, %) | 0.182 | 0.114 | 0.0176 | 0.184 | 0.184 | <0.0001 |
|  |  | **S2** | Number of participants | 6,154 | 13,433 | - | 5,452 | 5,452 | - |
|  |  |  | Follow-up (days) median (interquartile range) | 5,223 (1,403) | 4,959 (1,337) | - | 5,226.5 (1,406) | 4,922.5 (1,377.5) | - |
|  |  |  | Age at Index (years, SD) | 0.769 ± 0.801 | 0.652 ± 0.769 | 0.1482 | 0.759 ± 0.797 | 0.758 ± 0.796 | 0.0012 |
|  |  |  | Female (%) | 51.898 | 44.608 | 0.1463 | 51.559 | 51.522 | 0.0007 |
|  |  |  | Male (%) | 48.012 | 55.384 | 0.1479 | 48.423 | 48.459 | 0.0007 |
|  |  |  | Black or African American | 44.525 | 27.785 | 0.3538 | 44.149 | 44.534 | 0.0078 |
|  |  |  | White (%) | 33.721 | 47.343 | 0.2802 | 33.988 | 33.529 | 0.0097 |
|  |  |  | Hispanic or Latino (%) | 15.508 | 10.621 | 0.1454 | 15.591 | 15.187 | 0.0112 |
|  |  |  | Persons with potential health hazards related to socioeconomic and psychosocial circumstances (Z55-Z66, %) | 0.581 | 0.578 | 0.0004 | 0.587 | 0.679 | 0.0116 |
|  |  |  | Overweight and obesity (E66, %) | 0.581 | 0.415 | 0.0236 | 0.532 | 0.624 | 0.0121 |
|  |  |  | Family history of ischemic heart disease and other diseases of the circulatory system (Z82.4, %) | 0.182 | 0.081 | 0.0277 | 0.183 | 0.183 | <0.0001 |
|  |  |  | Nicotine dependence (F17, %) | 0.182 | 0.081 | 0.0277 | 0.183 | 0.183 | <0.0001 |
|  |  |  | Diabetes mellitus (E08-E13, %) | 0.182 | 0.081 | 0.0277 | 0.183 | 0 | 0.0606 |
|  |  |  | Disorders of lipoprotein metabolism and other lipidemias (E78, %) | 0.182 | 0.114 | 0.0176 | 0.183 | 0.183 | <0.0001 |
|  |  | **Female** | Number of participants | 3,212 | 6,018 | - | 2.808 | 2,808 | - |
|  |  |  | Follow-up (days) median (interquartile range) | 5,251 (1,454) | 4,959 (1,374) | - | 5,260.5 (1,459) | 4,934.5 (1,380) | - |
|  |  |  | Age at Index (years, SD) | 0.833 ± 0.818 | 0.717 ± 0.789 | 0.1442 | 0.813 ± 0.811 | 0.809 ± 0.81 | 0.0057 |
|  |  |  | Female (%) | 100 | 100 | - | 100 | 100 | - |
|  |  |  | Male (%) | 0 | 0 | - | 0 | 0 | - |
|  |  |  | Black or African American | 47.341 | 29.137 | 0.3813 | 46.581 | 47.471 | 0.0178 |
|  |  |  | White (%) | 30.896 | 45.576 | 0.3056 | 31.303 | 30.449 | 0.0185 |
|  |  |  | Hispanic or Latino (%) | 15.78 | 10.619 | 0.1529 | 15.954 | 14.993 | 0.0266 |
|  |  |  | Persons with potential health hazards related to socioeconomic and psychosocial circumstances (Z55-Z66, %) | 0.455 | 0.584 | 0.0179 | 0.427 | 0.392 | 0.0056 |
|  |  |  | Overweight and obesity (E66, %) | 0.665 | 0.712 | 0.0057 | 0.677 | 0.783 | 0.0126 |
|  |  |  | Family history of ischemic heart disease and other diseases of the circulatory system (Z82.4, %) | 0.35 | 0.182 | 0.0325 | 0.356 | 0.356 | <0.0001 |
|  |  |  | Nicotine dependence (F17, %) | 0 | 0.182 | 0.0605 | 0 | 0 | - |
|  |  |  | Diabetes mellitus (E08-E13, %) | 0.35 | 0 | 0.0838 | 0 | 0 | - |
|  |  |  | Disorders of lipoprotein metabolism and other lipidemias (E78, %) | 0.35 | 0.182 | 0.0325 | 0.356 | 0.356 | <0.0001 |
|  |  | **Male** | Number of participants | 2,937 | 7,414 | - | 2,636 | 2,636 | - |
|  |  |  | Follow-up (days) median (interquartile range) | 5,189.5 (1,358) | 4,959 (1,311) | - | 5,189.5 (1.358) | 4,938 (1,316) | - |
|  |  |  | Age at Index (years, SD) | 0.7 ± 0.777 | 0.6 ± 0.747 | 0.1309 | 0.699 ± 0.777 | 0.699 ± 0.776 | 0.0005 |
|  |  |  | Female (%) | 0 | 0 | - | 0 | 0 | - |
|  |  |  | Male (%) | 100 | 100 | - | 100 | 100 | - |
|  |  |  | Black or African American | 41.566 | 26.701 | 0.3174 | 41.54 | 41.654 | 0.0023 |
|  |  |  | White (%) | 36.838 | 48.758 | 0.2427 | 36.836 | 36.76 | 0.0016 |
|  |  |  | Hispanic or Latino (%) | 15.242 | 10.625 | 0.1379 | 15.212 | 15.175 | 0.0011 |
|  |  |  | Persons with potential health hazards related to socioeconomic and psychosocial circumstances (Z55-Z66, %) | 0.719 | 0.573 | 0.0182 | 0.721 | 0.645 | 0.0092 |
|  |  |  | Overweight and obesity (E66, %) | 0.492 | 0.176 | 0.0547 | 0.379 | 0.379 | <0.0001 |
|  |  |  | Family history of ischemic heart disease and other diseases of the circulatory system (Z82.4, %) | 0.378 | 0 | 0.0871 | 0 | 0 | - |
|  |  |  | Nicotine dependence (F17, %) | 0.378 | 0.147 | 0.0452 | 0.379 | 0 | 0.0873 |
|  |  |  | Diabetes mellitus (E08-E13, %) | 0 | 0.147 | 0.0543 | 0 | 0 | - |
|  |  |  | Disorders of lipoprotein metabolism and other lipidemias (E78, %) | 0.378 | 0.147 | 0.0452 | 0.379 | 0.379 | <0.0001 |
|  |  | **Black or African American** | Number of participants | 2,740 | 3,716 | - | 2,405 | 2,405 | - |
|  |  |  | Follow-up (days) median (interquartile range) | 5,280 (1,384) | 5,010 (1,437) | - | 5,288 (1,371) | 5,003 (1,447) | - |
|  |  |  | Age at Index (years, SD) | 0.73 ± 0.803 | 0.594 ± 0.759 | 0.1731 | 0.704 ± 0.791 | 0.704 ± 0.79 | 0.0005 |
|  |  |  | Female (%) | 55.112 | 46.836 | 0.1661 | 54.262 | 54.345 | 0.0017 |
|  |  |  | Male (%) | 44.888 | 53.164 | 0.1661 | 45.738 | 45.655 | 0.0017 |
|  |  |  | Black or African American | 100 | 100 | - | 100 | 100 | - |
|  |  |  | White (%) | 0 | 0 | - | 0 | 0 | - |
|  |  |  | Hispanic or Latino (%) | 2.525 | 2.868 | 0.0212 | 2.578 | 2.495 | 0.0053 |
|  |  |  | Persons with potential health hazards related to socioeconomic and psychosocial circumstances (Z55-Z66, %) | 0.896 | 1.005 | 0.0113 | 0.873 | 0.915 | 0.0044 |
|  |  |  | Overweight and obesity (E66, %) | 0.733 | 0.562 | 0.0214 | 0.748 | 0.707 | 0.0049 |
|  |  |  | Family history of ischemic heart disease and other diseases of the circulatory system (Z82.4, %) | 0.407 | 0 | 0.0904 | 0 | 0 | - |
|  |  |  | Nicotine dependence (F17, %) | 0.407 | 0.296 | 0.0189 | 0 | 0 | - |
|  |  |  | Diabetes mellitus (E08-E13, %) | 0 | 0 | - | 0 | 0 | - |
|  |  |  | Disorders of lipoprotein metabolism and other lipidemias (E78, %) | 0.407 | 0.296 | 0.0189 | 0 | 0 | - |
|  |  | **Hispanic** | Number of participants | 942 | 1,436 | - | 834 | 834 | - |
|  |  |  | Follow-up (days) median (interquartile range) | 5,195 (1,404) | 4,867 (1,250) | - | 5,196 (1,395) | 4,836 (1,240) | - |
|  |  |  | Age at Index (years, SD) | 0.838 ± 0.807 | 0.728 ± 0.787 | 0.1382 | 0.825 ± 0.805 | 0.818 ± 0.807 | 0.0089 |
|  |  |  | Female (%) | 52.723 | 44.3 | 0.1691 | 51.319 | 51.439 | 0.0024 |
|  |  |  | Male (%) | 47.277 | 55.7 | 0.1691 | 48.681 | 48.561 | 0.0024 |
|  |  |  | Black or African American | 7.184 | 7.422 | 0.0091 | 7.434 | 7.314 | 0.0046 |
|  |  |  | White (%) | 44.38 | 29.916 | 0.3028 | 42.446 | 42.806 | 0.0073 |
|  |  |  | Hispanic or Latino (%) | 100 | 100 | - | 100 | 100 | - |
|  |  |  | Persons with potential health hazards related to socioeconomic and psychosocial circumstances (Z55-Z66, %) | 1.159 | 0.765 | 0.0403 | 1.199 | 1.199 | <0.0001 |
|  |  |  | Overweight and obesity (E66, %) | 1.159 | 0.765 | 0.0403 | 1.199 | 1.199 | <0.0001 |
|  |  |  | Family history of ischemic heart disease and other diseases of the circulatory system (Z82.4, %) | 1.159 | 0 | 0.1531 | 0 | 0 | - |
|  |  |  | Nicotine dependence (F17, %) | 0 | 0.765 | 0.1242 | 0 | 0 | - |
|  |  |  | Diabetes mellitus (E08-E13, %) | 0 | 0.765 | 0.1242 | 0 | 0 | - |
|  |  |  | Disorders of lipoprotein metabolism and other lipidemias (E78, %) | 1.159 | 0.765 | 0.0403 | 1.199 | 1.199 | <0.0001 |
|  |  | **White** | Number of participants | 2,094 | 6,484 | - | 1,839 | 1,839 | - |
|  |  |  | Follow-up (days) median (interquartile range) | 5,181 (1,442) | 4,954 (1,336) | - | 5,171 (1,442) | 4,880 (1,289) | - |
|  |  |  | Age at Index (years, SD) | 0.834 ± 0.791 | 0.699 ± 0.77 | 0.1732 | 0.823 ± 0.789 | 0.826 ± 0.789 | 0.0034 |
|  |  |  | Female (%) | 47.517 | 42.896 | 0.0930 | 46.656 | 46.765 | 0.0022 |
|  |  |  | Male (%) | 52.422 | 50.868 | 0.0311 | 52.399 | 52.399 | <0.0001 |
|  |  |  | Black or African American | 0 | 0 | - | 0 | 0 | - |
|  |  |  | White (%) | 100 | 100 | - | 100 | 100 | - |
|  |  |  | Hispanic or Latino (%) | 20.448 | 6.614 | 0.4130 | 19.141 | 19.141 | <0.0001 |
|  |  |  | Persons with potential health hazards related to socioeconomic and psychosocial circumstances (Z55-Z66, %) | 0.534 | 0.389 | 0.0214 | 0.544 | 0.544 | <0.0001 |
|  |  |  | Overweight and obesity (E66, %) | 0.534 | 0.237 | 0.0480 | 0.544 | 0.544 | <0.0001 |
|  |  |  | Family history of ischemic heart disease and other diseases of the circulatory system (Z82.4, %) | 0.534 | 0.169 | 0.0617 | 0.544 | 0.544 | <0.0001 |
|  |  |  | Nicotine dependence (F17, %) | 0.534 | 0.169 | 0.0617 | 0.544 | 0.544 | <0.0001 |
|  |  |  | Diabetes mellitus (E08-E13, %) | 0.534 | 0.169 | 0.0617 | 0 | 0.644 | 0.1046 |
|  |  |  | Disorders of lipoprotein metabolism and other lipidemias (E78, %) | 0.534 | 0.169 | 0.0617 | 0.544 | 0.544 | <0.0001 |
| **Persistent (cases) versus no AD (controls)** | **Cardiovascular diseases** | **Primary** | Number of participants | 6,828 | 182,928 | - | 4,963 | 4,963 | - |
|  |  |  | Follow-up (days) median (interquartile range) | 5,299 (1,412) | 4,913 (1,137) | - | 5,298 (1,413) | 4,829 (1,193) | - |
|  |  |  | Age at Index (years, SD) | 0.754 ± 0.797 | 0.549 ± 0.778 | 0.2596 | 0.753 ± 0.798 | 0.753 ± 0.798 | 0.0003 |
|  |  |  | Female (%) | 51.669 | 48.833 | 0.0567 | 51.662 | 51.662 | <0.0001 |
|  |  |  | Male (%) | 48.231 | 51.125 | 0.0579 | 48.237 | 48.217 | 0.0004 |
|  |  |  | Black or African American | 42.943 | 19.704 | 0.5176 | 42.857 | 42.918 | 0.0012 |
|  |  |  | White (%) | 35.283 | 56.438 | 0.4344 | 35.342 | 35.301 | 0.0008 |
|  |  |  | Hispanic or Latino (%) | 17.129 | 21.659 | 0.1148 | 17.147 | 17.107 | 0.0011 |
|  |  |  | Persons with potential health hazards related to socioeconomic and psychosocial circumstances (Z55-Z66, %) | 0.643 | 0.304 | 0.0120 | 0.645 | 0.645 | <0.0001 |
|  |  |  | Overweight and obesity (E66, %) | 0.623 | 0.05 | 0.0990 | 0.443 | 0.403 | 0.0062 |
|  |  |  | Family history of ischemic heart disease and other diseases of the circulatory system (Z82.4, %) | 0.201 | 0.113 | 0.0222 | 0.201 | 0.201 | <0.0001 |
|  |  |  | Nicotine dependence (F17, %) | 0.201 | 0.007 | 0.0603 | 0 | 0 | - |
|  |  |  | Diabetes mellitus (E08-E13, %) | 0.201 | 0.022 | 0.0536 | 0.201 | 0.201 | <0.0001 |
|  |  |  | Disorders of lipoprotein metabolism and other lipidemias (E78, %) | 0.201 | 0.09 | 0.0292 | 0.201 | 0.201 | <0.0001 |
|  |  | **S1** | Number of participants | 7,434 | 114,361 | - | 5,506 | 5,506 | - |
|  |  |  | Follow-up (days) median (interquartile range) | 5,222 (1,398) | 5,102 (1,444) | - | 5,222 (1,396) | 4,967 (1,462) | - |
|  |  |  | Age at Index (years, SD) | 0.766 ± 0.799 | 0.556 ± 0.783 | 0.2657 | 0.765 ± 0.8 | 0.766 ± 0.8 | 0.0011 |
|  |  |  | Female (%) | 51.829 | 49.111 | 0.0544 | 51.762 | 51.762 | <0.0001 |
|  |  |  | Male (%) | 48.08 | 50.824 | 0.0549 | 48.147 | 48.147 | <0.0001 |
|  |  |  | Black or African American | 44.422 | 22.45 | 0.4789 | 44.333 | 44.388 | 0.0011 |
|  |  |  | White (%) | 33.865 | 52.348 | 0.3799 | 33.927 | 33.927 | <0.0001 |
|  |  |  | Hispanic or Latino (%) | 15.556 | 12.677 | 0.0828 | 15.565 | 15.529 | 0.0010 |
|  |  |  | Persons with potential health hazards related to socioeconomic and psychosocial circumstances (Z55-Z66, %) | 0.58 | 0.243 | 0.0527 | 0.581 | 0.563 | 0.0024 |
|  |  |  | Overweight and obesity (E66, %) | 0.58 | 0.042 | 0.0967 | 0.345 | 0.363 | 0.0031 |
|  |  |  | Family history of ischemic heart disease and other diseases of the circulatory system (Z82.4, %) | 0.181 | 0.016 | 0.0528 | 0.182 | 0.182 | <0.0001 |
|  |  |  | Nicotine dependence (F17, %) | 0.181 | 0.012 | 0.545 | 0.182 | 0 | 0.0603 |
|  |  |  | Diabetes mellitus (E08-E13, %) | 0.181 | 0.018 | 0.0517 | 0.182 | 0.182 | <0.0001 |
|  |  |  | Disorders of lipoprotein metabolism and other lipidemias (E78, %) | 0.181 | 0.084 | 0.0267 | 0.182 | 0.182 | <0.0001 |
|  |  | **S2** | Number of participants | 6,174 | 92,761 | - | 5,512 | 5,512 | - |
|  |  |  | Follow-up (days) median (interquartile range) | 5,223 (1,399) | 5,103 (1,446) | - | 5,225.5 (1,397) | 4,931 (1,491) | - |
|  |  |  | Age at Index (years, SD) | 0.767 ± 0.8 | 0.557 ± 0.784 | 0.2648 | 0.766 ± 0.8 | 0.767 ± 0.8 | 0.0011 |
|  |  |  | Female (%) | 51.828 | 49.114 | 0.0543 | 51.778 | 51.742 | 0.0007 |
|  |  |  | Male (%) | 48.082 | 50.822 | 0.0558 | 48.131 | 48.168 | 0.0007 |
|  |  |  | Black or African American | 44.408 | 22.466 | 0.4782 | 44.303 | 44.376 | 0.0015 |
|  |  |  | White (%) | 33.876 | 52.339 | 0.3795 | 44.303 | 44.376 | 0.0015 |
|  |  |  | Hispanic or Latino (%) | 15.563 | 12.68 | 0.0828 | 15.584 | 15.53 | 0.0015 |
|  |  |  | Persons with potential health hazards related to socioeconomic and psychosocial circumstances (Z55-Z66, %) | 0.579 | 0.244 | 0.0524 | 0.581 | 0.544 | 0.0049 |
|  |  |  | Overweight and obesity (E66, %) | 0.0963 | 0.043 | 0.0963 | 0.363 | 0.399 | 0.0059 |
|  |  |  | Family history of ischemic heart disease and other diseases of the circulatory system (Z82.4, %) | 0.181 | 0.016 | 0.0528 | 0.181 | 0.181 | <0.0001 |
|  |  |  | Nicotine dependence (F17, %) | 0.181 | 0.012 | 0.0544 | 0.181 | 0 | 0.0603 |
|  |  |  | Diabetes mellitus (E08-E13, %) | 0.181 | 0.019 | 0.0512 | 0.181 | 0.181 | <0.0001 |
|  |  |  | Disorders of lipoprotein metabolism and other lipidemias (E78, %) | 0.181 | 0.084 | 0.0267 | 0.181 | 0.181 | <0.0001 |
|  |  | **Female** | Number of participants | 3,219 | 45,629 | - | 2,850 | 2,850 | - |
|  |  |  | Follow-up (days) median (interquartile range) | 5,249.5 (1,450.5) | 5,124 (1,478.5) | - | 5,248 (1,452) | 4,935.5 (1,476) | - |
|  |  |  | Age at Index (years, SD) | 0.831 ± 0.817 | 0.558 ± 0.785 | 0.3402 | 0.829 ± 0.818 | 0.829 ± 0.817 | 0.0004 |
|  |  |  | Female (%) | 100 | 100 | - | 100 | 100 | - |
|  |  |  | Male (%) | 0 | 0 | - | 0 | 0 | - |
|  |  |  | Black or African American | 47.207 | 22.502 | 0.5368 | 47.123 | 47.123 | <0.0001 |
|  |  |  | White (%) | 31.075 | 52.392 | 0.4428 | 31.123 | 31.158 | 0.0008 |
|  |  |  | Hispanic or Latino (%) | 15.852 | 12.627 | 0.0924 | 15.754 | 15.824 | 0.0019 |
|  |  |  | Persons with potential health hazards related to socioeconomic and psychosocial circumstances (Z55-Z66, %) | 0.454 | 0.22 | 0.0404 | 0.421 | 0.421 | <0.0001 |
|  |  |  | Overweight and obesity (E66, %) | 0.663 | 0.034 | 0.1069 | 0.351 | 0.351 | <0.0001 |
|  |  |  | Family history of ischemic heart disease and other diseases of the circulatory system (Z82.4, %) | 0.349 | 0.024 | 0.0753 | 0.351 | 0.351 | <0.0001 |
|  |  |  | Nicotine dependence (F17, %) | 0 | 0.024 | 0.0221 | 0 | 0 | - |
|  |  |  | Diabetes mellitus (E08-E13, %) | 0.349 | 0.078 | 0.0753 | 0.351 | 0.351 | <0.0001 |
|  |  |  | Disorders of lipoprotein metabolism and other lipidemias (E78, %) | 0.349 | 0.078 | 0.0587 | 0.351 | 0.351 | <0.0001 |
|  |  | **Male** | Number of participants | 2,950 | 47,076 | - | 2,651 | 2,651 | - |
|  |  |  | Follow-up (days) median (interquartile range) | 5,189 (1,356) | 5,086 (1,415) | - | 5,186 (1.355) | 4,909 (1,368) | - |
|  |  |  | Age at Index (years, SD) | 0.699 ± 0.775 | 0.555 ± 0.783 | 0.1840 | 0.699 ± 0.776 | 0.7 ± 0.776 | 0.0015 |
|  |  |  | Female (%) | 0 | 0 | - | 0 | 0 | - |
|  |  |  | Male (%) | 100 | 100 | - | 100 | 100 | - |
|  |  |  | Black or African American | 41.475 | 22.436 | 0.4171 | 41.456 | 41.456 | <0.0001 |
|  |  |  | White (%) | 36.959 | 52.285 | 0.3120 | 36.967 | 36.967 | <0.0001 |
|  |  |  | Hispanic or Latino (%) | 15.28 | 12.746 | 0.0731 | 15.315 | 15.24 | 0.0021 |
|  |  |  | Persons with potential health hazards related to socioeconomic and psychosocial circumstances (Z55-Z66, %) | 0.715 | 0.267 | 0.0642 | 0.717 | 0.679 | 0.0045 |
|  |  |  | Overweight and obesity (E66, %) | 0.489 | 0.052 | 0.0843 | 0.377 | 0.377 | <0.0001 |
|  |  |  | Family history of ischemic heart disease and other diseases of the circulatory system (Z82.4, %) | 0.376 | 0.024 | 0.790 | 0 | 0 | - |
|  |  |  | Nicotine dependence (F17, %) | 0.376 | 0 | 0.0869 | 0 | 0 | - |
|  |  |  | Diabetes mellitus (E08-E13, %) | 0 | 0.024 | 0.0217 | 0 | 0 | - |
|  |  |  | Disorders of lipoprotein metabolism and other lipidemias (E78, %) | 0.376 | 0.09 | 0.0595 | 0.377 | 0.377 | <0.0001 |
|  |  | **Black or African American** | Number of participants | 2,740 | 21,717 | - | 2,441 | 2,441 | - |
|  |  |  | Follow-up (days) median (interquartile range) | 5,280 (1,384) | 4,985 (1,480) | - | 5,278 (1,376) | 4,813 (1,441) | - |
|  |  |  | Age at Index (years, SD) | 0.73 ± 0.803 | 0.54 ± 0.792 | 0.2376 | 0.727 ± 0.803 | 0.727 ± 0.803 | <0.0001 |
|  |  |  | Female (%) | 55.094 | 49.193 | 0.1183 | 54.977 | 54.977 | <0.0001 |
|  |  |  | Male (%) | 44.906 | 50.753 | 0.1173 | 45.023 | 45.023 | <0.0001 |
|  |  |  | Black or African American | 100 | 100 | - | 100 | 100 | - |
|  |  |  | White (%) | 0 | 0 | - | 0 | 0 | - |
|  |  |  | Hispanic or Latino (%) | 2.526 | 3.505 | 0.572 | 2.54 | 2.54 | <0.0001 |
|  |  |  | Persons with potential health hazards related to socioeconomic and psychosocial circumstances (Z55-Z66, %) | 0.896 | 0.454 | 0.0540 | 0.901 | 0.901 | <0.0001 |
|  |  |  | Overweight and obesity (E66, %) | 0.733 | 0.059 | 0.1076 | 0.41 | 0.41 | <0.0001 |
|  |  |  | Family history of ischemic heart disease and other diseases of the circulatory system (Z82.4, %) | 0.407 | 0.053 | 0.0739 | 0 | 0 | - |
|  |  |  | Nicotine dependence (F17, %) | 0.407 | 0 | 0.0905 | 0 | 0 | - |
|  |  |  | Diabetes mellitus (E08-E13, %) | 0 | 0.053 | 0.0327 | 0 | 0 | - |
|  |  |  | Disorders of lipoprotein metabolism and other lipidemias (E78, %) | 6,828 | 182,928 | - | 4,963 | 4,963 | - |
|  |  | **Hispanic** | Number of participants | 939 | 11,749 | - | 855 | 855 | - |
|  |  |  | Follow-up (days) median (interquartile range) | 5,199 (1,396) | 4,950 (1,354) | - | 5,197 (1,397) | 4,881 (1,321) | - |
|  |  |  | Age at Index (years, SD) | 0.834 ± 0.805 | 0.636 ± 0.806 | 0.2454 | 0.834 ± 0.782 | 0.833 ± 0.808 | 0.0014 |
|  |  |  | Female (%) | 52.791 | 48.907 | 0.0777 | 52.632 | 52.632 | <0.0001 |
|  |  |  | Male (%) | 47.209 | 51.084 | 0.0776 | 47.368 | 47.368 | <0.0001 |
|  |  |  | Black or African American | 7.209 | 6.209 | 0.0400 | 7.251 | 7.485 | 0.0090 |
|  |  |  | White (%) | 44.419 | 34.018 | 0.2142 | 44.327 | 44.327 | <0.0001 |
|  |  |  | Hispanic or Latino (%) | 100 | 100 | - | 100 | 100 | - |
|  |  |  | Persons with potential health hazards related to socioeconomic and psychosocial circumstances (Z55-Z66, %) | 1.163 | 0.104 | 0.1337 | 1.17 | 1.17 | <0.0001 |
|  |  |  | Overweight and obesity (E66, %) | 1.163 | 0.095 | 0.1354 | 1.17 | 1.17 | <0.0001 |
|  |  |  | Family history of ischemic heart disease and other diseases of the circulatory system (Z82.4, %) | 1.163 | 0.095 | 0.1354 | 0 | 0 | - |
|  |  |  | Nicotine dependence (F17, %) | 0 | 0 | - | 0 | 0 | - |
|  |  |  | Diabetes mellitus (E08-E13, %) | 0 | 0.095 | 0.1050 | 0 | 0 | - |
|  |  |  | Disorders of lipoprotein metabolism and other lipidemias (E78, %) | 1.163 | 0.123 | 0.1304 | 1.17 | 1.17 | <0.0001 |
|  |  | **White** | Number of participants | 2,093 | 47,993 | - | 1,868 | 1,868 | - |
|  |  |  | Follow-up (days) median (interquartile range) | 5,181 (1,442.5) | 5,178 (1,445) | - | 5,181 (1,437.5) | 5,033 (1,455.5) | - |
|  |  |  | Age at Index (years, SD) | 0.833 ± 0.791 | 0.555 ± 0.775 | 0.3553 | 0.834 ± 0.791 | 0.835 ± 0.792 | 0.0014 |
|  |  |  | Female (%) | 47.543 | 49.164 | 0.0324 | 47.484 | 47.537 | 0.0011 |
|  |  |  | Male (%) | 52.457 | 50.769 | 0.0338 | 52.516 | 52.463 | 0.0011 |
|  |  |  | Black or African American | 0 | 0 | - | 0 | 0 | - |
|  |  |  | White (%) | 100 | 100 | - | 100 | 100 | - |
|  |  |  | Hispanic or Latino (%) | 20.406 | 8.242 | 0.3526 | 20.289 | 20.289 | <0.0001 |
|  |  |  | Persons with potential health hazards related to socioeconomic and psychosocial circumstances (Z55-Z66, %) | 0.534 | 0.188 | 0.0577 | 0.535 | 0.535 | <0.0001 |
|  |  |  | Overweight and obesity (E66, %) | 0.534 | 0.025 | 0.0965 | 0.535 | 0.535 | <0.0001 |
|  |  |  | Family history of ischemic heart disease and other diseases of the circulatory system (Z82.4, %) | 0.534 | 0.023 | 0.0971 | 0.535 | 0.535 | <0.0001 |
|  |  |  | Nicotine dependence (F17, %) | 0.534 | 0.023 | 0.0971 | 0.535 | 0 | 0.1038 |
|  |  |  | Diabetes mellitus (E08-E13, %) | 0.534 | 0.023 | 0.0971 | 0.535 | 0.535 | <0.0001 |
|  |  |  | Disorders of lipoprotein metabolism and other lipidemias (E78, %) | 0.534 | 0.11 | 0.0749 | 0.535 | 0.535 | <0.0001 |
| **Transient (cases) versus no AD (controls)** | **Cardiovascular diseases** | **Primary** | Number of participants | 25,173 | 186,263 | - | 17,992 | 17,992 | - |
|  |  |  | Follow-up (days) median (interquartile range) | 4,934 (1,307) | 4,812 (1,110) | - | 4,934 (1,307) | 4,781 (1,134) | - |
|  |  |  | Age at Index (years, SD) | 0.668 ± 0.795 | 0.597 ± 0.796 | 0.0904 | 0.667 ± 0.774 | 0.668 ± 0.775 | 0.0009 |
|  |  |  | Female (%) | 44.64 | 48.867 | 0.0848 | 44.609 | 44.581 | 0.0006 |
|  |  |  | Male (%) | 55.355 | 51.092 | 0.0855 | 55.386 | 55.414 | 0.0006 |
|  |  |  | Black or African American | 30.913 | 21.963 | 0.2040 | 30.841 | 30.83 | 0.0002 |
|  |  |  | White (%) | 42.865 | 53.743 | 0.2190 | 42.886 | 42.914 | 0.0006 |
|  |  |  | Hispanic or Latino (%) | 18.557 | 22.228 | 0.0912 | 18.547 | 18.536 | 0.0003 |
|  |  |  | Persons with potential health hazards related to socioeconomic and psychosocial circumstances (Z55-Z66, %) | 0.749 | 0.319 | 0.0590 | 0.745 | 0.734 | 0.0013 |
|  |  |  | Overweight and obesity (E66, %) | 0.483 | 0.055 | 0.0826 | 0.306 | 0.3 | 0.0010 |
|  |  |  | Family history of ischemic heart disease and other diseases of the circulatory system (Z82.4, %) | 0.205 | 0.118 | 0.0217 | 0.206 | 0.2 | 0.0012 |
|  |  |  | Nicotine dependence (F17, %) | 0.055 | 0.007 | 0.0276 | 0 | 0.056 | 0.0334 |
|  |  |  | Diabetes mellitus (E08-E13, %) | 0.055 | 0.025 | 0.0152 | 0.056 | 0.056 | <0.0001 |
|  |  |  | Disorders of lipoprotein metabolism and other lipidemias (E78, %) | 0.139 | 0.099 | 0.0115 | 0.139 | 0.145 | 0.0015 |
|  |  | **S1** | Number of participants | 18,235 | 145,905 | - | 12,322 | 12,322 | - |
|  |  |  | Follow-up (days) median (interquartile range) | 4,956 (1,335) | 4,979 (1,225) | - | 4.956 (1,334) | 4,929.5 (1,250) | - |
|  |  |  | Age at Index (years, SD) | 0.649 ± 0.767 | 0.588 ± 0.794 | 0.0784 | 0.648 ± 0.767 | 0.648 ± 0.767 | 0.0006 |
|  |  |  | Female (%) | 44.604 | 49.156 | 0.0913 | 44.546 | 44.514 | 0.0001 |
|  |  |  | Male (%) | 55.388 | 50.792 | 0.0922 | 55.446 | 55.478 | 0.0007 |
|  |  |  | Black or African American | 27.342 | 21.213 | 0.1433 | 27.285 | 27.285 | <0.0001 |
|  |  |  | White (%) | 47.85 | 58.199 | 0.2085 | 47.882 | 47.874 | 0.0002 |
|  |  |  | Hispanic or Latino (%) | 10.582 | 10.257 | 0.0106 | 10.566 | 10.55 | 0.0005 |
|  |  |  | Persons with potential health hazards related to socioeconomic and psychosocial circumstances (Z55-Z66, %) | 0.575 | 0.212 | 0.0579 | 0.568 | 0.568 | <0.0001 |
|  |  |  | Overweight and obesity (E66, %) | 0.389 | 0.039 | 0.0758 | 0.211 | 0.211 | <0.0001 |
|  |  |  | Family history of ischemic heart disease and other diseases of the circulatory system (Z82.4, %) | 0.081 | 0.018 | 0.0280 | 0.081 | 0.081 | <0.0001 |
|  |  |  | Nicotine dependence (F17, %) | 0.081 | 0.009 | 0.0341 | 0 | 0 | - |
|  |  |  | Diabetes mellitus (E08-E13, %) | 0.081 | 0.013 | 0.0312 | 0.081 | 0.081 | <0.0001 |
|  |  |  | Disorders of lipoprotein metabolism and other lipidemias (E78, %) | 0.113 | 0.063 | 0.168 | 0.114 | 0.106 | 0.0025 |
|  |  | **S2** | Number of participants | 12,884 | 105,672 | - | 11,708 | 11,708 | - |
|  |  |  | Follow-up (days) median (interquartile range) | 4,980 (1,364) | 5,115 (1,324) | - | 4,980.5 (1,364.5) | 5,070 (1,372) | - |
|  |  |  | Age at Index (years, SD) | 0.645 ± 0.765 | 0.542 ± 0.776 | 0.1340 | 0.644 ± 0.765 | 0.644 ± 0.765 | 0.0001 |
|  |  |  | Female (%) | 44.602 | 49.025 | 0.0887 | 44.542 | 44.482 | 0.0012 |
|  |  |  | Male (%) | 55.39 | 50.915 | 0.0898 | 55.449 | 55.509 | 0.0012 |
|  |  |  | Black or African American | 27.576 | 19.631 | 0.1879 | 27.537 | 27.52 | 0.0004 |
|  |  |  | White (%) | 47.15 | 57.919 | 0.2169 | 47.181 | 47.181 | <0.0001 |
|  |  |  | Hispanic or Latino (%) | 10.618 | 11.083 | 0.0150 | 10.591 | 10.591 | <0.0001 |
|  |  |  | Persons with potential health hazards related to socioeconomic and psychosocial circumstances (Z55-Z66, %) | 0.605 | 0.247 | 0.0551 | 0.598 | 0.598 | <0.0001 |
|  |  |  | Overweight and obesity (E66, %) | 0.418 | 0.042 | 0.0786 | 0.231 | 0.231 | <0.0001 |
|  |  |  | Family history of ischemic heart disease and other diseases of the circulatory system (Z82.4, %) | 0.085 | 0.015 | 0.0316 | 0.085 | 0.085 | <0.0001 |
|  |  |  | Nicotine dependence (F17, %) | 0.085 | 0.01 | 0.0342 | 0.085 | 0.085 | <0.0001 |
|  |  |  | Diabetes mellitus (E08-E13, %) | 0.085 | 0.015 | 0.0316 | 0.085 | 0.085 | <0.0001 |
|  |  |  | Disorders of lipoprotein metabolism and other lipidemias (E78, %) | 0.119 | 0.07 | 0.0160 | 0.12 | 0.102 | 0.0051 |
|  |  | **Female** | Number of participants | 5,771 | 51,896 | - | 5,208 | 5,208 | - |
|  |  |  | Follow-up (days) median (interquartile range) | 4,985.5 (1,410) | 5,134 (1,355) | - | 4,987.5 (1,411.5) | 5,068.5 (1,382.5) | - |
|  |  |  | Age at Index (years, SD) | 0.706 ± 0.784 | 0.544 ± 0.778 | 0.2074 | 0.702 ± 0.782 | 0.702 ± 0.782 | 0.0007 |
|  |  |  | Female (%) | 100 | 100 | - | 100 | 100 | - |
|  |  |  | Male (%) | 0 | 0 | - | 0 | 0 | - |
|  |  |  | Black or African American | 29.175 | 19.652 | 0.2231 | 29.147 | 29.09 | 0.0013 |
|  |  |  | White (%) | 45.166 | 57.915 | 0.2572 | 45.219 | 45.219 | <0.0001 |
|  |  |  | Hispanic or Latino (%) | 10.566 | 11.085 | 0.0167 | 10.522 | 10.522 | <0.0001 |
|  |  |  | Persons with potential health hazards related to socioeconomic and psychosocial circumstances (Z55-Z66, %) | 0.63 | 0.226 | 0.1109 | 0.269 | 0.269 | 0.0024 |
|  |  |  | Overweight and obesity (E66, %) | 0.707 | 0.034 | 0.1109 | 0.269 | 0.269 | <0.0001 |
|  |  |  | Family history of ischemic heart disease and other diseases of the circulatory system (Z82.4, %) | 0.191 | 0.021 | 0.0521 | 0.192 | 0.192 | <0.0001 |
|  |  |  | Nicotine dependence (F17, %) | 0.191 | 0.021 | 0.0521 | 0 | 0 | - |
|  |  |  | Diabetes mellitus (E08-E13, %) | 0 | 0 | 0.0521 | 0 | 0 | - |
|  |  |  | Disorders of lipoprotein metabolism and other lipidemias (E78, %) | 0.191 | 0.066 | 0.0349 | 0.192 | 0.192 | <0.0001 |
|  |  | **Male** | Number of participants | 7,112 | 53,717 | - | 6,493 | 6,493 | - |
|  |  |  | Follow-up (days) median (interquartile range) | 4,977 (1,337) | 5,099 (1,295) | - | 4,977 (1.337) | 5,064 (1,337) | - |
|  |  |  | Age at Index (years, SD) | 0.596 ± 0.747 | 0.539 ± 0.774 | 0.0751 | 0.596 ± 0.747 | 0.596 ± 0.747 | 0.0002 |
|  |  |  | Female (%) | 0 | 0 | - | 0 | 0 | - |
|  |  |  | Male (%) | 100 | 100 | - | 100 | 100 | - |
|  |  |  | Black or African American | 26.292 | 19.614 | 0.1593 | 26.244 | 26.244 | <0.0001 |
|  |  |  | White (%) | 48.738 | 57.927 | 0.1850 | 48.76 | 48.745 | 0.0003 |
|  |  |  | Hispanic or Latino (%) | 10.662 | 11.092 | 0.0138 | 10.642 | 10.642 | 0.0005 |
|  |  |  | Persons with potential health hazards related to socioeconomic and psychosocial circumstances (Z55-Z66, %) | 0.585 | 0.267 | 0.0488 | 0.57 | 0.57 | <0.0001 |
|  |  |  | Overweight and obesity (E66, %) | 0.185 | 0.049 | 0.0396 | 0.154 | 0.154 | <0.0001 |
|  |  |  | Family history of ischemic heart disease and other diseases of the circulatory system (Z82.4, %) | 0 | 0.021 | 0.0203 | 0 | 0 | - |
|  |  |  | Nicotine dependence (F17, %) | 0.154 | 0 | 0.0555 | 0 | 0 | - |
|  |  |  | Diabetes mellitus (E08-E13, %) | 0.154 | 0.021 | 0.0452 | 0.154 | 0.154 | <0.0001 |
|  |  |  | Disorders of lipoprotein metabolism and other lipidemias (E78, %) | 0.154 | 0.074 | 0.0237 | 0.154 | 0.154 | <0.0001 |
|  |  | **Black or African American** | Number of participants | 3,567 | 21,877 | - | 3,221 | 3,221 | - |
|  |  |  | Follow-up (days) median (interquartile range) | 5,019.5 (1,464.5) | 5,033 (1,484) | - | 5,019 (1,465) | 4,941 (1,469) | - |
|  |  |  | Age at Index (years, SD) | 0.593 ± 0.756 | 0.525 ± 0.784 | 0.0892 | 0.591 ± 0.756 | 0.591 ± 0.756 | 0.0004 |
|  |  |  | Female (%) | 47.188 | 49.076 | 0.0378 | 47.097 | 47.006 | 0.0006 |
|  |  |  | Male (%) | 52.812 | 50.87 | 0.0389 | 52.903 | 52.934 | 0.0006 |
|  |  |  | Black or African American | 100 | 100 | - | 100 | 100 | - |
|  |  |  | White (%) | 0 | 0 | - | 0 | 0 | - |
|  |  |  | Hispanic or Latino (%) | 2.936 | 3.535 | 0.0339 | 2.887 | 2.918 | 0.0018 |
|  |  |  | Persons with potential health hazards related to socioeconomic and psychosocial circumstances (Z55-Z66, %) | 1.051 | 0.458 | 0.0686 | 1.025 | 1.025 | <0.0001 |
|  |  |  | Overweight and obesity (E66, %) | 0.587 | 0.053 | 0.0946 | 0.31 | 0.31 | <0.0001 |
|  |  |  | Family history of ischemic heart disease and other diseases of the circulatory system (Z82.4, %) | 0 | 0.053 | 0.0326 | 0 | 0 | - |
|  |  |  | Nicotine dependence (F17, %) | 0.309 | 0 | 0.0787 | 0 | 0 | - |
|  |  |  | Diabetes mellitus (E08-E13, %) | 0 | 0.053 | 0.0326 | 0 | 0 | - |
|  |  |  | Disorders of lipoprotein metabolism and other lipidemias (E78, %) | 0.309 | 0.053 | 0.0602 | 0.31 | 0.31 | <0.0001 |
|  |  | **Hispanic** | Number of participants | 1,394 | 11,749 | - | 1,253 | 1,253 | - |
|  |  |  | Follow-up (days) median (interquartile range) | 4,852 (1,290) | 4,950 (1,354) | - | 4,854 (1,290) | 4,926 (1,392) | - |
|  |  |  | Age at Index (years, SD) | 0.724 ± 0.784 | 0.636 ± 0.806 | 0.1108 | 0.718 ± 0.782 | 0.718 ± 0.782 | <0.0001 |
|  |  |  | Female (%) | 44.506 | 48.907 | 0.0883 | 44.453 | 44.374 | 0.0016 |
|  |  |  | Male (%) | 55.494 | 51.084 | 0.0885 | 55.547 | 55.626 | 0.0016 |
|  |  |  | Black or African American | 7.51 | 6.209 | 0.0515 | 7.422 | 7.422 | <0.0001 |
|  |  |  | White (%) | 29.723 | 34.018 | 0.0923 | 29.449 | 29.529 | 0.0018 |
|  |  |  | Hispanic or Latino (%) | 100 | 100 | - | 100 | 100 | - |
|  |  |  | Persons with potential health hazards related to socioeconomic and psychosocial circumstances (Z55-Z66, %) | 0.791 | 0.104 | 0.1030 | 0.798 | 0.798 | <0.0001 |
|  |  |  | Overweight and obesity (E66, %) | 0.87 | 0.104 | 0.1030 | 0.798 | 0.798 | <0.0001 |
|  |  |  | Family history of ischemic heart disease and other diseases of the circulatory system (Z82.4, %) | 0 | 0.095 | 0.435 | 0 | 0 | - |
|  |  |  | Nicotine dependence (F17, %) | 0.791 | 0 | 0.1262 | 0 | 0 | - |
|  |  |  | Diabetes mellitus (E08-E13, %) | 0.791 | 0.095 | 0.1050 | 0 | 0 | - |
|  |  |  | Disorders of lipoprotein metabolism and other lipidemias (E78, %) | 0.791 | 0.123 | 0.0991 | 0.798 | 0.798 | <0.0001 |
|  |  | **White** | Number of participants | 6,104 | 60,201 | - | 5,522 | 5,522 | - |
|  |  |  | Follow-up (days) median (interquartile range) | 4,997 (1,381) | 5,157 (1,243) | - | 4,999 (1,381) | 5,115 (1,249) | - |
|  |  |  | Age at Index (years, SD) | 0.691 ± 0.767 | 0.539 ± 0.768 | 0.1985 | 0.689 ± 0.767 | 0.689 ± 0.766 | 0.0005 |
|  |  |  | Female (%) | 42.725 | 49.022 | 0.1266 | 42.666 | 42.629 | 0.0007 |
|  |  |  | Male (%) | 57.256 | 50.922 | 0.1274 | 57.316 | 57.352 | 0.0007 |
|  |  |  | Black or African American | 0 | 0 | - | 0 | 0 | - |
|  |  |  | White (%) | 100 | 100 | - | 100 | 100 | - |
|  |  |  | Hispanic or Latino (%) | 6.904 | 6.967 | 0.0025 | 6.845 | 6.827 | 0.0007 |
|  |  |  | Persons with potential health hazards related to socioeconomic and psychosocial circumstances (Z55-Z66, %) | 0.416 | 0.197 | 0.0396 | 0.417 | 0.417 | <0.0001 |
|  |  |  | Overweight and obesity (E66, %) | 0.253 | 0.029 | 0.0598 | 0.181 | 0.181 | <0.0001 |
|  |  |  | Family history of ischemic heart disease and other diseases of the circulatory system (Z82.4, %) | 0.181 | 0.018 | 0.0516 | 0.181 | 0.181 | <0.0001 |
|  |  |  | Nicotine dependence (F17, %) | 0.181 | 0.018 | 0.0516 | 0 | 0 | - |
|  |  |  | Diabetes mellitus (E08-E13, %) | 0.181 | 0.018 | 0.0516 | 0.181 | 0.181 | <0.0001 |
|  |  |  | Disorders of lipoprotein metabolism and other lipidemias (E78, %) | 0.181 | 0.087 | 0.0258 | 0.181 | 0.181 | <0.0001 |

**Supplement Table 3**. Baseline characteristics before and after propensity-score matching for all analyses relating to the risk of cardiovascular diseases in patients with persistent, transient or no childhood atopic dermatitis (AD).

| **Outcome** | **Analysis** | **Persistent AD**  **% \| total n** | **No AD**  **% \| total n** | **Hazard ratio (CI 95%)** | **p** | **Chi-square** |
| --- | --- | --- | --- | --- | --- | --- |
| Asthma (J45) | Primary | 53.36 \| 5,250 | 25.77 \| 5,250 | 2.419 (2.267, 2.581) | <0.0001 | 45.47 |
|  | S1 | 52.00 \| 5,156 | 23.84 \| 5,218 | 2.538 (2.373, 2.715) | <0.0001 | 45.464 |
|  | S2 | 53.26 \| 5,310 | 24.99 \| 5,310 | 2.49 (2.333, 2.659) | <0.0001 | 61.6 |
|  | Female | 47.35 \| 2,758 | 22.48 \| 2,758 | 2.38 (2.163, 2.619) | <0.0001 | 16.973 |
|  | Male | 59.64 \| 2,547 | 27.60 \| 2,547 | 2.651 (2.423, 2.899) | <0.0001 | 35.888 |
|  | Black or African American | 59.56 \| 2,438 | 29.53 \| 2,438 | 2.437 (2.228, 2.665) | <0.0001 | 12.881 |
|  | Hispanic or Latino | 48.14 \| 912 | 25 \| 912 | 2.172 (1.85, 2.549) | <0.0001 | 2.12 |
|  | White | 49.39 \| 1,802 | 21.87 \| 1,802 | 2.639 (2.343, 2.972) | <0.0001 | 5.437 |
| Allergic rhinitis (J30) | Primary | 66.78 \| 5,250 | 35.31 \| 5,250 | 2.384 (2.254, 2.523) | <0.0001 | 15.144 |
|  | S1 | 66.53 \| 5,207 | 34.87 \| 5,257 | 2.359 (2.229, 2.496) | <0.0001 | 49.804 |
|  | S2 | 67.12 \| 5,310 | 36.65 \| 5,310 | 2.257 (2.136, 2.386) | <0.0001 | 45.11 |
|  | Female | 63.62 \| 2,758 | 32.81 \| 2,758 | 2.367 (2.184, 2.565) | <0.0001 | 11.091 |
|  | Male | 70.91 \| 2,547 | 36.95 \| 2,547 | 2.469 (2.281, 2.672) | <0.0001 | 33.314 |
|  | Black or African American | 71.74 \| 2,438 | 42.45 \| 2,438 | 2.062 (1.909, 2.228) | <0.0001 | 24.876 |
|  | Hispanic or Latino | 64.70 \| 912 | 35.31 \| 912 | 2.182 (1.904, 2.5) | <0.0001 | 13.642 |
|  | White | 65.10 \| 1,802 | 33.57 \| 1,802 | 2.407 (2.181, 2.656) | <0.0001 | 16.8 |
| Chronic sinusitis (J32) | Primary | 11.94 \| 5,250 | 10.04 \| 5,250 | 1.146 (1.021, 1.287) | <0.0001 | 21.802 |
|  | S1 | 10.87 \| 5,289 | 7.73 \| 5,294 | 1.357 (1.195, 1.541) | <0.0001 | 11.898 |
|  | S2 | 11.15 \| 5,310 | 8.10 \| 5,310 | 1.331 (1.175, 1.507) | <0.0001 | 5.442 |
|  | Female | 10.37 \| 2,758 | 7.03 \| 2,758 | 1.407 (1.172, 1.69) | 0.0002 | 0.893 |
|  | Male | 11.98 \| 2,547 | 7.34 \| 2,547 | 1.613 (1.344, 1.935) | <0.0001 | 3.123 |
|  | Black or African American | 8.37 \| 2,438 | 5.95 \| 2,438 | 1.34 (1.082, 1.659) | 0.0070 | 7.216 |
|  | Hispanic or Latino | 8.89 \| 912 | 5.48 \| 912 | 1.549 (1.087, 2.206) | 0.0146 | 1.352 |
|  | White | 16.54 \| 1,802 | 11.38 \| 1,802 | 1.425 (1.192, 1.702) | <0.0001 | 2.397 |
| Eosinophilic esophagitis (K20.0) | Primary | 1.94 \| 5,250 | 0.21 \| 5,250 | 8.722 (4.68, 16.252) | <0.0001 | 0.267 |
|  | S1 | 1.96 \| 5,299 | 0.34 \| 5,304 | 5.326 (3.227, 8.79) | <0.0001 | 5.198 |
|  | S2 | 2.07 \| 5,310 | 0.19 \| 5,310 | 11.341 (5.745, 22.387) | <0.0001 | 8.395 |
|  | Female | 1.49 \| 2,758 | 0.36 \| 2,758 | 37.699 (5.183, 274.206) | <0.0001 | 1.557 |
|  | Male | 2.71 \| 2,547 | 0.43 \| 2,547 | 6.006 (3.177, 11.354) | <0.0001 | 0.421 |
|  | Black or African American | 1.93 \| 2,438 | 0.41 \| 2,438 | 42.424 (5.848, 307.748) | <0.0001 | 3.318 |
|  | Hispanic or Latino | 1.21 \| 912 | 1.10 \| 912 | 5.247 (1.167, 23.826) | 0.0157 | 0.825 |
|  | White | 2.39 \| 1,802 | 0.56 \| 1,802 | 5.773 (2.596, 12.841) | <0.0001 | 1.216 |
| Food allergy status (Z91.01) or anaphylactic reaction due to food (T78.0) | Primary | 32.19 \| 5,250 | 3.96 \| 5,250 | 9.18 (7.948, 10.602) | <0.0001 | 0.261 |
|  | S1 | 31.08 \| 5,241 | 4.13 \| 5,298 | 8.334 (7.237, 9.598) | <0.0001 | 0.027 |
|  | S2 | 31.90 \| 5,310 | 3.82 \| 5,310 | 9.369 (8.099, 10.838) | <0.0001 | 1.402 |
|  | Female | 29.12 \| 2,758 | 3.34 \| 2,758 | 9.534 (7.683, 11.831) | <0.0001 | 4.366 |
|  | Male | 35.02 \| 2,547 | 4.08 \| 2,547 | 9.951 (8.121, 12.193) | <0.0001 | 0.072 |
|  | Black or African American | 31.13 \| 2,438 | 3.86 \| 2,438 | 8.826 (7.122, 10.937) | <0.0001 | 0.011 |
|  | Hispanic or Latino | 25.88 \| 912 | 2.41 \| 912 | 11.646 (7.521, 18.032) | <0.0001 | 0.255 |
|  | White | 34.41 \| 1,802 | 3.89 \| 1,802 | 10.236 (7.994, 13.107) | <0.0001 | 3.752 |
| Any T2ID | Primary | 80.46 \| 5,250 | 50.91 \| 5,250 | 2.109 (2.008, 2.214) | <0.0001 | 43.861 |
|  | S1 | 79.27 \| 5,013 | 47.44 \| 5,158 | 2.214 (2.105, 2.329) | <0.0001 | 69.324 |
|  | S2 | 80.38 \| 5,310 | 50.09 \| 5,310 | 2.14 (2.038, 2.246) | <0.0001 | 57.151 |
|  | Female | 77.63 \| 2,758 | 45.36 \| 2,758 | 2.275 (2.121, 2.44) | <0.0001 | 18.464 |
|  | Male | 83.27 \| 2,547 | 50.22 \| 2,547 | 2.327 (2.17, 2.496) | <0.0001 | 41.571 |
|  | Black or African American | 84.37 \| 2,438 | 54.02 \| 2,438 | 2.108 (1.967, 2.26) | <0.0001 | 33.302 |
|  | Hispanic or Latino | 75.77 \| 912 | 46.82 \| 912 | 2.042 (1.809, 2.305) | <0.0001 | 8.542 |
|  | White | 78.14 \| 1,802 | 47.50 \| 1,802 | 2.215 (2.034, 2.413) | <0.0001 | 8.827 |
| Autoimmune diseases | Primary | 4.22 \| 4,856 | 2.30 \| 4,861 | 1.679 (1.333, 2.116) | <0.0001 | 0.329 |
|  | S1 | 4.22 \| 5,470 | 2.25 \| 5,477 | 1.783 (1.432, 2.219) | <0.0001 | 0.053 |
|  | S2 | 4.27 \| 5,476 | 1.95 \| 5,481 | 2.094 (1.666, 2.633) | <0.0001 | 0.141 |
|  | Female | 4.62 \| 2,837 | 2.36 \| 2,843 | 1.847 (1.375, 2.48) | <0.0001 | 0.153 |
|  | Male | 3.88 \| 2,631 | 1.82 \| 2,632 | 2.03 (1.44, 2.862) | <0.0001 | 0.051 |
|  | Black or African American | 3.73 \| 2,437 | 1.44 \| 2,437 | 2.449 (1.657, 3.618) | <0.0001 | 0.097 |
|  | Hispanic or Latino | 5.92 \| 844 | 2.13 \| 847 | 2.744 (1,601, 4.703) | 0.0001 | 0.839 |
|  | White | 4.48 \| 1,851 | 2.70 \| 1,854 | 1.633 (1.15, 2.32) | 0.0057 | 0.164 |
| VTE | Primary | 0.28 \| 4,961 | 0.20 \| 4,961 | 1.374 (0.592, 3.187) | 0.4577 | 0.355 |
|  | S1 | 0.24 \| 5,503 | 0.18 \| 5,506 | 2.008 (0.763, 5.285) | 0.1498 | 0.523 |
|  | S2 | 0.25 \| 5,510 | 0.18 \| 5,511 | 2.178 (0.836, 5.617) | 0.1022 | 0.131 |
|  | Female | 0.35 \| 2,850 | 0.35 \| 2,850 | 3.664 (0.777, 17.27) | 0.0786 | 0.549 |
|  | Male | 0.38 \| 2,650 | 0 \| 2,650 | - | 0.0192 | - |
|  | Black or African American | 0.41 \| 2,440 | 0.41 \| 2,441 | 4.427 (0.516, 37.995) | 0.1379 | 0.959 |
|  | Hispanic or Latino | 1.17 \| 855 | 1.17 \| 855 | 1.217 (0.203, 7.304) | 0.8298 | 0.102 |
|  | White | 0.54 \| 1,868 | 0 \| 1,868 | - | 0.0328 | - |
| MACE | Primary | 0.53 \| 4,938 | 0.77 \| 4,942 | 0.633 (0.384, 1.044) | 0.0705 | 3.334 |
|  | S1 | 0.46 \| 5,475 | 0.38 \| 5,488 | 1.128 (0.631, 2.105) | 0.6851 | 0.542 |
|  | S2 | 0.49 \| 5,483 | 0.40 \| 5,493 | 1.165 (0.663, 2.046) | 0.5949 | 0.397 |
|  | Female | 0.39 \| 2,837 | 0.39 \| 2,845 | 0.961 (0.461, 2.218) | 0.9257 | 0.941 |
|  | Male | 0.61 \| 2,635 | 0.72 \| 2,635 | 0.78 (0.401, 1.517) | 0.4626 | 0.337 |
|  | Black or African American | 0.58 \| 2,427 | 0.66 \| 2,432 | 0.795 (0.387, 1.631) | 0.5302 | 0.344 |
|  | Hispanic or Latino | 1.18 \| 848 | 1.17 \| 854 | 1.272 (0.284, 5.961) | 0.7523 | 2.074 |
|  | White | 0.54 \| 1,860 | 0.54 \| 1,863 | 1.133 (0.38, 3.371) | 0.8228 | 0.365 |
| Cardiovascular risk factors | Primary | 12.06 \| 4,942 | 7.09 \| 4,942 | 1.511 (1.323, 1.725) | <0.0001 | 0.149 |
|  | S1 | 11.53 \| 5,481 | 8.35 \| 5,483 | 1.315 (1.166, 1.483) | <0.0001 | 0 |
|  | S2 | 11.57 \| 5,489 | 8.81 \| 5,494 | 1.252 (1.113, 1.41) | 0.0002 | 0.379 |
|  | Female | 11.19 \| 2,842 | 8.76 \| 2,843 | 1.191 (1.009, 1.406) | 0.0389 | 4.925 |
|  | Male | 12.02 \| 2,637 | 8.21 \| 2,642 | 1.387 (1.167, 1.649) | 0.0002 | 0.137 |
|  | Black or African American | 12.25 \| 2,432 | 9.59 \| 2,439 | 1.157 (0.975, 1.373) | 0.0955 | 0.549 |
|  | Hispanic or Latino | 15.04 \| 851 | 14.19 \| 853 | 0.978 (0.762, 1.254) | 0.8599 | 0 |
|  | White | 10.37 \| 1,861 | 7.09 \| 1,861 | 1.418 (1.136, 1.77) | 0.0019 | 3.017 |

**Supplement Table 4**. Risks of other type 2 inflammatory diseases (T2IDs), autoimmune disease, and cardiovascular diseases contrasted amongst patients with persistent versus no childhood atopic dermatitis (AD). To counter the bias introduced by multiple testing, a Bonferroni correction was applied, adjusting the significance threshold based on the number of outcomes tested within each disease category: type-2 inflammatory diseases (T2IDs, n=6; adjusted α=0.0083), autoimmune diseases (n=1; adjusted α=0.05), and cardiovascular outcomes (n=3; adjusted α=0.0167). “-“ indicates value cannot	be calculated die to zero outcomes in at least one group. *Abbreviations:* ***VTE****: Venous thromboembolism,* ***MACE****: Major adverse cardiac events.*

| **Outcome** | **Analysis** | **Persistent AD**  **% \| total n** | **Transient AD**  **% \| total n** | **Hazard ratio (CI 95%)** | **p** | **Chi-square** |
| --- | --- | --- | --- | --- | --- | --- |
| Asthma (J45) | Primary | 53.24 \| 5,302 | 29.56 \| 5,302 | 2.094 (1.968, 2.227) | <0.0001 | 12.849 |
|  | S1 | 51.48 \| 4,705 | 28.22 \| 4,766 | 2.112 (1.976, 2.258) | <0.0001 | 11.246 |
|  | S2 | 53.10 \| 4,989 | 29.71 \| 4,989 | 2.066 (1.939, 2.202) | <0.0001 | 21.124 |
|  | Female | 47.90 \| 2,499 | 27.57 \| 2,499 | 1.942 (1.769, 2.134) | <0.0001 | 4.404 |
|  | Male | 60.50 \| 2,352 | 33.16 \| 2,352 | 2.245 (2.057, 2.45) | <0.0001 | 1.818 |
|  | Black or African American | 59.97 \| 2,091 | 35.49 \| 2,091 | 2.013 (1.838, 2.205) | <0.0001 | 6.847 |
|  | Hispanic or Latino | 48.29 \| 905 | 24.42 \| 905 | 2.268 (1.928, 2.666) | <0.0001 | 0.967 |
|  | White | 49.48 \| 1,837 | 26.40 \| 1,837 | 2.176 (1.949, 2.43) | <0.0001 | 0.785 |
| Allergic rhinitis (J30) | Primary | 66.60 \| 5,302 | 36.01 \| 5,302 | 2.33 (2.204, 2.464) | <0.0001 | 13.846 |
|  | S1 | 66.51 \| 4,760 | 37.54 \| 4,811 | 2.218 (2.093, 2.35) | <0.0001 | 17.617 |
|  | S2 | 67.53 \| 4,989 | 38.49 \| 4,989 | 2.199 (2.079, 2.326) | <0.0001 | 22.11 |
|  | Female | 62.95 \| 2,499 | 36.38 \| 2,499 | 2.064 (1.902, 2.24) | <0.0001 | 8.625 |
|  | Male | 71.30 \| 2,352 | 40.94 \| 2,352 | 2.256 (2.083, 2.443) | <0.0001 | 8.386 |
|  | Black or African American | 71.07 \| 2,091 | 41.42 \| 2,901 | 2.215 (2.036, 2.409) | <0.0001 | 1.662 |
|  | Hispanic or Latino | 64.53 \| 905 | 35.91 \| 905 | 2.162 (1.887, 2.477) | <0.0001 | 6.92 |
|  | White | 64.94 \| 1,837 | 36.75 \| 1,837 | 2.2 (2.002, 2.419) | <0.0001 | 5.581 |
| Chronic sinusitis (J32) | Primary | 11.94 \| 5,302 | 7.92 \| 5,302 | 1.483 (1.311, 1.678) | <0.0001 | 6.259 |
|  | S1 | 12.50 \| 4,826 | 8.67\| 4,834 | 1.428 (1.26, 1.617) | <0.0001 | 3.626 |
|  | S2 | 12.71 \| 4,989 | 9.16 \| 4,989 | 1.37 (1.214, 1.545) | <0.0001 | 4.315 |
|  | Female | 11.12 \| 2,499 | 7.64 \| 2,499 | 1.419 (1.18, 1.707) | 0.0002 | 2.593 |
|  | Male | 12.59 \| 2,352 | 8.76 \| 2,352 | 1.414 (1.183, 1.689) | 0.0001 | 1.263 |
|  | Black or African American | 7.7 \| 2,091 | 4.36 \| 2,091 | 1.743 (1.348, 2.255) | <0.0001 | 4.29 |
|  | Hispanic or Latino | 8.95 \| 905 | 4.31 \| 905 | 1.986 (1.354, 2.912) | 0.0003 | 0.207 |
|  | White | 16.93 \| 1,837 | 10.78 \| 1,837 | 1.572 (1.316, 1.879) | <0.0001 | 0.749 |
| Eosinophilic esophagitis (K20.0) | Primary | 1.92 \| 5,302 | 0.43 \| 5,302 | 4.227 (2.687, 6.647) | <0.0001 | 0.8 |
|  | S1 | 1.88 \| 4.841 | 0.62 \| 4.846 | 2.886 (1.91, 4.362) | <0.0001 | 4.87 |
|  | S2 | 2.09 \| 4,989 | 0.42 \| 4,989 | 4.771 (2.985, 7.626) | <0.0001 | 0.412 |
|  | Female | 1.48 \| 2,499 | 0.4 \| 2,499 | 4.301 (2.001, 9.243) | <0.0001 | 0.439 |
|  | Male | 2.59 \| 2,352 | 0.68 \| 2,352 | 3.664 (2.112, 6.356) | <0.0001 | 3.644 |
|  | Black or African American | 1.91 \| 2,091 | 0.48 \| 2,091 | 7.693 (3.035, 19.499) | <0.0001 | 1.234 |
|  | Hispanic or Latino | 1.22 \| 905 | 1.11 \|905 | 3.531 (0.984, 12.672) | 0.0389 | 0.222 |
|  | White | 2.29 \| 1,837 | 1.47 \| 1,837 | 1.484 (0.915, 2,407) | 0.1077 | 2.187 |
| Food allergy status (Z91.01) or anaphylactic reaction due to food (T78.0) | Primary | 32.27 \| 5,302 | 6.70 \| 5,302 | 5.445 (4.857, 6.105) | <0.0001 | 6.451 |
|  | S1 | 32.83 \| 4,782 | 8.16 \| 4.827 | 4.552 (4.076, 5.084) | <0.0001 | 3.173 |
|  | S2 | 33.61 \| 4,989 | 8.46 \| 4,989 | 4.519 (4.061, 5.028) | <0.0001 | 10.506 |
|  | Female | 29.17 \| 2,499 | 7.96 \| 2,499 | 3.968 (3.392, 4.641) | <0.0001 | 0.054 |
|  | Male | 35.97 \| 2,352 | 8.67 \| 2,352 | 4.783 (4.104, 5.573) | <0.0001 | 2.839 |
|  | Black or African American | 29.94 \| 2,091 | 7.41 \| 2,091 | 4.474 (3.753, 5.5335) | <0.0001 | 0.132 |
|  | Hispanic or Latino | 25.75 \| 905 | 4.64 \| 905 | 5.955 (4.286, 8.273) | <0.0001 | 0.014 |
|  | White | 34.62 \| 1,837 | 10.72 \| 1,837 | 3.648 (3.109, 4.281) | <0.0001 | 0.561 |
| Any T2ID | Primary | 80.39 \| 5,302 | 52.47 \| 5,302 | 2.076 (1.978, 2.178) | <0.0001 | 17.368 |
|  | S1 | 79.51 \| 4,568 | 52.80 \| 4,708 | 2 (1.9, 2.105) | <0.0001 | 19.254 |
|  | S2 | 80.64 \| 4,989 | 54.14 \| 4,989 | 1.986 (1.891, 2.086) | <0.0001 | 24.62 |
|  | Female | 77.31 \| 2,499 | 52.10 \| 2,499 | 1.884 (1.756, 2.022) | <0.0001 | 9.577 |
|  | Male | 83.67 \| 2,352 | 58.29 \| 2,352 | 2.003 (1.868, 2.147) | <0.0001 | 4.165 |
|  | Black or African American | 84.27 \| 2,091 | 57.58 \| 2,091 | 1.989 (1.848, 2.142) | <0.0001 | 14.886 |
|  | Hispanic or Latino | 75.69 \| 905 | 48.18 \| 905 | 2.041 (1.809, 2.303) | <0.0001 | 2.524 |
|  | White | 78.06 \| 1,837 | 53.78 \| 1,837 | 1.917 (1.767, 2.079) | <0.0001 | 1.936 |
| Autoimmune diseases | Primary | 4.28 \| 5,146 | 2.67 \| 5,141 | 1.185 (1.132, 1.24) | <0.0001 | 38.089 |
|  | S1 | 4.30 \| 5,342 | 2.16 \| 5,333 | 1.938 (1.549, 2.424) | <0.0001 | 0.285 |
|  | S2 | 4.34 \| 5,254 | 2.69 \| 5,244 | 1.571 (1.274, 1.939) | <0.0001 | 2.575 |
|  | Female | 4.67 \| 2,744 | 2.66 \| 2,740 | 1.692 (1.269, 2.255) | 0.0003 | 0.368 |
|  | Male | 3.92 \| 2,651 | 2.42 \| 2,650 | 1.57 (1.15, 2.144) | 0.0042 | 0.361 |
|  | Black or African American | 3.74 \| 2,379 | 2.36 \| 2,372 | 1.548 (1.108, 2.162) | 0.0098 | 0.0241 |
|  | Hispanic or Latino | 5.90 \| 848 | 1.89 \| 849 | 3.048 (1.735, 5.353) | <0.0001 | 0.008 |
|  | White | 4.49 \| 1,869 | 2.52 \| 1,865 | 1.772 (1.24, 2.532) | 0.0015 | 0.008 |
| VTE | Primary | 0.26 \| 5,302 | 0.19 \| 5,302 | 3.096 (1.017, 9.424) | 0.0360 | 1.414 |
|  | S1 | 0.24 \| 5,445 | 0.18 \| 5,445 | 2.915 (9.949, 8.953) | 0.0502 | 0.01 |
|  | S2 | 0.26 \| 5,450 | 0.18 \| 5,451 | 3.151 (1,036, 9.585) | 0.0328 | 0.676 |
|  | Female | 0.36 \| 2,808 | 0.36 \| 2,808 | 7.377 (0.921, 59.061) | 0.0271 | 1.164 |
|  | Male | 0.38 \| 2,634 | 0.38 \| 3,635 | 1.846 (0.461, 7.39) | 0.3792 | 0.136 |
|  | Black or African American | 0.42 \| 2,404 | 0 \| 2,405 | - | 0.0305 | - |
|  | Hispanic or Latino | 1.20 \| 834 | 1.2 \| 833 | 2.407 (0.249, 23.265) | 0.4336 | 0.6188 |
|  | White | 0.54 \| 1,839 | 0.54 \| 1,839 | 2.019 (0.39, 10.443) | 0.3925 | 0.013 |
| MACE | Primary | 0.49 \| 5,276 | 0.68 \| 5,285 | 0.662 (0.399, 1.098) | 0.1075 | 0.063 |
|  | S1 | 0.48 \| 5,417 | 0.33 \| 5,430 | 1.336 (0.732, 2.439) | 0.3431 | 0 |
|  | S2 | 0.50 \| 5,424 | 0.48 \| 5,437 | 0.97 (0.565, 1.663) | 0.9107 | 0.266 |
|  | Female | 0.36 \| 2,796 | 0.57 \| 2,800 | 0.59 (0.268, 1.302) | 0.1864 | 0.409 |
|  | Male | 0.65 \| 2,620 | 0.38 \| 2,628 | 1.782 (0.794, 4) | 0.1559 | 1.374 |
|  | Black or African American | 0.58 \| 2,391 | 0.42 \| 2,398 | 1.868 (0.753, 4.633) | 0.1706 | 0.143 |
|  | Hispanic or Latino | 1.21 \| 827 | 1.2 \| 832 | 1.879 (0.344, 10.277) | 0.4596 | 1.981 |
|  | White | 0.55 \| 1,831 | 0.55 \| 1,836 | 0.856 (0.31, 2.361) | 0.7636 | 1.58 |
| Cardiovascular risk factors | Primary | 11.94 \| 5,283 | 7.91 \| 5,295 | 1.382 (1.221, 1.564) | <0.0001 | 2.127 |
|  | S1 | 11.65 \| 5,423 | 8.11 \| 5,429 | 1.332 (1.179, 1.504) | <0.0001 | 0.005 |
|  | S2 | 11.70 \| 5,429 | 8.37 \| 5,438 | 1.302 (1.155, 1.469) | <0.0001 | 1.465 |
|  | Female | 11.18 \| 2,800 | 7.75 \| 2,799 | 1.328 (1.117, 1.58) | <0.0013 | 0.13 |
|  | Male | 12.09 \| 2,622 | 7.80 \| 2,628 | 1.461 (1.225, 1.742) | <0.0001 | 1.039 |
|  | Black or African American | 12.23 \| 2,396 | 9.30 \| 2,397 | 1.234 (1.036, 1.469) | 0.0179 | 0.559 |
|  | Hispanic or Latino | 15.30 \| 830 | 10.48 \| 830 | 1.364 (1.038, 1.792) | 0.0255 | 0.045 |
|  | White | 10.15 \| 1,832 | 6.76 \| 1,834 | 1.373 (1.093, 1.724) | 0.0062 | 5.096 |

**Supplement Table 5**. Risks of other type 2 inflammatory diseases (T2IDs), autoimmune disease, and cardiovascular diseases contrasted amongst patients with persistent versus transient childhood atopic dermatitis (AD). To counter the bias introduced by multiple testing, a Bonferroni correction was applied, adjusting the significance threshold based on the number of outcomes tested within each disease category: type-2 inflammatory diseases (T2IDs, n=6; adjusted α=0.0083), autoimmune diseases (n=1; adjusted α=0.05), and cardiovascular outcomes (n=3; adjusted α=0.0167). *Abbreviations:* ***VTE****: Venous thromboembolism,* ***MACE****: Major adverse cardiac events.*

| **Outcome** | **Analysis** | **Transient AD**  **% \| total n** | **No AD**  **% \| total n** | **Hazard ratio (CI 95%)** | **p** | **Chi-square** |
| --- | --- | --- | --- | --- | --- | --- |
| Asthma (J45) | Primary | 29.03 \|13,950 | 24.56 \| 13,950 | 1.185 (1.132, 1.24) | <0.0001 | 38.089 |
|  | S1 | 29.26 \| 15,953 | 23.59 \| 16,008 | 1.268 (1.215, 1.324) | <0.0001 | 84.797 |
|  | S2 | 30.51 \| 15,325 | 24.74 \| 15,325 | 1.266 (1.213, 1.322) | <0.0001 | 71.033 |
|  | Female | 26.39 \| 6,840 | 21.17 \| 6,840 | 1.289 (1.203, 1.382) | <0.0001 | 10.075 |
|  | Male | 33.38 \| 7,184 | 26.13 \| 7,184 | 1.328 (1.25, 1.411) | <0.0001 | 48.977 |
|  | Black or African American | 37.57 \| 3,785 | 29.78 \| 3,785 | 1.318 (1.219, 1.426) | <0.0001 | 8.759 |
|  | Hispanic or Latino | 26.67 \| 1,815 | 21.49 \| 1,815 | 1.281 (1.121, 1.463) | 0.0003 | 0.205 |
|  | White | 18.06 \| 5,437 | 20.53 \| 5,437 | 0.859 (0.789, 0.936) | 0.0005 | 1.375 |
| Allergic rhinitis (J30) | Primary | 29.03 \| 13,950 | 24.56 \| 13,950 | 1.185 (1.132, 1.24) | <0.0001 | 38.089 |
|  | S1 | 37.24 \| 16,064 | 33.6 \| 16,101 | 1.138 (1.097, 1.18) | <0.0001 | 3.935 |
|  | S2 | 38.41 \| 15,325 | 34.11 \| 15,325 | 1.164 (1.122, 1.209) | <0.0001 | 5.308 |
|  | Female | 35.18 \| 6,840 | 32.19 \| 6,840 | 1.12 (1.057, 1.187) | 0.0001 | 4.473 |
|  | Male | 40.48 \| 7,184 | 34.35 \| 7,184 | 1.262 (1.197, 1.332) | <0.0001 | 8.125 |
|  | Black or African American | 44.31 \| 3,785 | 40.21 \| 3,785 | 1.133 (1.057, 1.214) | 0.0004 | 1.028 |
|  | Hispanic or Latino | 31.80 \| 1,815 | 27.49 \| 1,815 | 1.184 (1.05, 1.335) | 0.0056 | 0.565 |
|  | White | 36.56 \| 5,437 | 33.42 \| 5,437 | 1.11 (1.041, 1.183) | 0.0013 | 0.003 |
| Chronic sinusitis (J32) | Primary | 9.56 \| 13,950 | 9.94 \| 13,950 | 0.94 (0.872, 1.014) | 0.1081 | 6.635 |
|  | S1 | 7.48 \| 16,195 | 7.94 \| 16,175 | 0.94 (0.869, 1.017) | 0.1235 | 13.77 |
|  | S2 | 7.99 \| 15,325 | 8.13 \| 15,325 | 0.985 (0.911, 1.066) | 0.7143 | 15.172 |
|  | Female | 7.91 \| 6,840 | 8.19 \| 6,840 | 0.972 (0.864, 1.094) | 0.6360 | 3.834 |
|  | Male | 8.69 \| 7,184 | 8.95 \| 7,184 | 0.981 (0.879, 1.095) | 0.7354 | 2.455 |
|  | Black or African American | 4.65 \| 3,785 | 5.76 \| 3,785 | 0.807 (0.661, 0.984) | 0.0338 | 1.72 |
|  | Hispanic or Latino | 5.95 \| 1,815 | 4.19 \| 1,815 | 1.423 (1.061, 1.909) | 0.0178 | 1.223 |
|  | White | 12.95 \| 5,437 | 10.80 \| 5,437 | 1.202 (1.077, 1.341) | 0.0010 | 3.862 |
| Eosinophilic esophagitis (K20.0) | Primary | 0.53 \| 13,950 | 0.24 \| 13,950 | 2.09 (1.392, 3.137) | 0.0003 | 0.407 |
|  | S1 | 0.68 \| 16,229 | 0.34 \| 16,226 | 2.022 (1,463, 2.795) | <0.0001 | 0.757 |
|  | S2 | 0.67 \| 15,325 | 0.3 \| 15,325 | 2.302 (1.626, 3.26) | <0.0001 | 3.73 |
|  | Female | 0.34 \| 6,840 | 0.15 \| 6,840 | 6.025 (2.082, 17.429) | 0.0002 | 0.004 |
|  | Male | 0.92 \| 7,184 | 0.38 \| 7,184 | 2.543 (1.625, 3.981) | <0.0001 | 0.588 |
|  | Black or African American | 0.29 \| 3,785 | 0.26 \| 3,785 | 2.825 (0.898, 8.882) | 0.0633 | 0.011 |
|  | Hispanic or Latino | 0.55 \| 1,815 | 0.55 \| 1,815 | 2.516 (0.789, 8.022) | 0.1062 | 3.153 |
|  | White | 0.74 \| 5,437 | 0.24 \| 5,437 | 3.051 (1.631, 5.706) | 0.0002 | 0.07 |
| Food allergy status (Z91.01) or anaphylactic reaction due to food (T78.0) | Primary | 7.53 \| 13,950 | 4.26 \| 13,950 | 1.759 (1.591, 1.946) | <0.0001 | 0.19 |
|  | S1 | 8.87 \| 16,159 | 3.80 \| 16,199 | 2.411 (2.194, 2.65) | <0.0001 | 0.2 |
|  | S2 | 9.49 \| 15,325 | 3.63 \| 15,325 | 2.731 (2.476, 3.011) | <0.0001 | 1.621 |
|  | Female | 8,47 \| 6,840 | 3.41 \| 6,840 | 2.592 (2.226, 3.018) | <0.0001 | 0.332 |
|  | Male | 10.26 \| 7,184 | 4.00 \| 7,184 | 2.707 (2.3362, 3.103) | <0.0001 | 0.086 |
|  | Black or African American | 8.61 \| 3,785 | 5.44 \| 3,785 | 1.611 (1.353, 1.918) | <0.0001 | 0.753 |
|  | Hispanic or Latino | 4.63 \| 1,815 | 2.92 \| 1,815 | 1.572 (1.115, 2.218) | 0.0093 | 0.604 |
|  | White | 5.22 \| 5,437 | 3.16 \| 5,437 | 1.66 (1.374, 2.006) | <0.0001 | 3.852 |
| Any T2ID | Primary | 53.33 \| 13,950 | 48.27 \| 13,950 | 1.121 (1.085, 1.159) | <0.0001 | 19.713 |
|  | S1 | 52.69 \| 15,713 | 47.04 \| 15,811 | 1.161 (1.125, 1.198) | <0.0001 | 38.121 |
|  | S2 | 54.70 \| 15,325 | 48.20 \| 15,325 | 1.19 (1.153, 1.227) | <0.0001 | 35.683 |
|  | Female | 50.72 \| 6,840 | 45.47 \| 6,840 | 1.164 (1.109, 1.221) | <0.0001 | 9.384 |
|  | Male | 57.70 \| 7,184 | 50.13 \| 7,184 | 1.244 (1.189, 1.3) | <0.0001 | 2.725 |
|  | Black or African American | 60.56 \| 3,785 | 53.84 \| 3,785 | 1.184 (1.115, 1.257) | <0.0001 | 6.445 |
|  | Hispanic or Latino | 47.22 \| 1,815 | 39.17 \| 1,815 | 1.281 (1.16, 1.416) | <0.0001 | 1.317 |
|  | White | 49.95 \| 5,437 | 46.44 \| 5,437 | 1.095 (1.038, 1.156) | 0.0010 | 0.116 |
| Autoimmune diseases | Primary | 2.62 \| 18,044 | 2.25 \| 18,076 | 1.134 (0.993, 1.295) | 0.0637 | 2.62 \|18,044 |
|  | S1 | 2.31 \| 12,027 | 2.02 \| 12,058 | 1.124 (0.946, 1.335) | 0.1835 | 0.983 |
|  | S2 | 2.36 \| 12,055 | 2.11 \| 12.086 | 1.092 (0.923, 1.294) | 0.3054 | 1.154 |
|  | Female | 2.56 \| 4,928 | 2.43 \| 4,934 | 1.063 (0.828, 1.365) | 0.06305 | 0.7.769 |
|  | Male | 2.29 \| 6,154 | 2.14 \| 6,167 | 1.074 (0.847, 1.361) | 0.5579 | 0.306 |
|  | Black or African American | 2.11 \| 2,937 | 1.97 \| 2,934 | 1.063 (0.743, 1.521) | 0.7385 | 1.31 |
|  | Hispanic or Latino | 2.25 \| 1,158 | 1.16 \| 1,161 | 1.548 (0.84, 2.852) | 0.1582 | 1.788 |
|  | White | 2.79 \| 5,524 | 2.66 \| 5,532 | 1.069 (0.853, 1.34) | 0.5618 | 1.205 |
| VTE | Primary | 0.07 \| 17,985 | 0.07 \| 17,988 | 0.953 (0.442, 2.057) | 0.9026 | 1.138 |
|  | S1 | 0.08 \| 12,318 | 0.09 \| 12,320 | 0.635 (0.246, 1.638) | 0.3435 | 0.126 |
|  | S2 | 0.09 \| 11,705 | 0.09 \| 11,708 | 1.174 (0.426, 3.237) | 0.7567 | 0.061 |
|  | Female | 0.19 \| 5,207 | 0.19 \| 5,208 | 0.408 (0.079, 2.103) | 0.2681 | 3.9 |
|  | Male | 0.15 \| 6,491 | 0.15 \| 6,493 | 1.658 (0.466, 5.898) | 0.4304 | 5.481 |
|  | Black or African American | 0.31 \| 3,221 | 0.31 \| 3,221 | 0.329 (0.034, 3.161) | 0.3107 | 0.249 |
|  | Hispanic or Latino | 0.80 \| 1,252 | 0.80 \| 1,253 | 0.532 (0.048, 5.872) | 0.6008 | 2.133 |
|  | White | 0.18 \| 5,520 | 0.18 \| 5,522 | 3.281 (0.661, 16.29) | 0.1237 | 0.283 |
| MACE | Primary | 0.66 \| 17,925 | 0.67 \| 17,919 | 0.955 (0.741, 1.232) | 0.7245 | 0.078 |
|  | S1 | 0.39 \| 12,287 | 0.39 \| 12,278 | 0.994 (0.666, 1.483) | 0.9776 | 0.6633 |
|  | S2 | 0.45 \| 11,680 | 0.57 \| 11,682 | 0.787 (0.548, 1.131) | 0.1939 | 3.553 |
|  | Female | 0.42 \| 5,195 | 0.42 \| 5,196 | 1.011 (0.56, 1.826) | 0.9709 | 0.431 |
|  | Male | 0.45 \| 6,478 | 0.71 \| 6,478 | 0.644 (0.404, 1.025) | 0.0612 | 2.058 |
|  | Black or African American | 0.31 \| 3,212 | 0.81 \| 3,205 | 0.343 (0.161, 0.733) | 0.0038 | 2.733 |
|  | Hispanic or Latino | 0.8 \| 1,250 | 0.80 \| 1,251 | 0.333 (0.035, 3.205) | 0.3173 | 0.266 |
|  | White | 0.53 \| 5,511 | 0.45 \| 5,502 | 1.185 (0.694, 2.023) | 0.5344 | 0.027 |
| Cardiovascular risk factors | Primary | 7.94 \| 17,937 | 7.94 \| 17,921 | 0.958 (0.89, 1.031) | 0.2555 | 3.444 |
|  | S1 | 7.31 \| 12.277 | 6.83 \| 12,290 | 1.069 (0.973, 1.175) | 0.1619 | 0.445 |
|  | S2 | 7.46 \| 11.675 | 7.04 \| 11,684 | 1.1 (1, 1.21) | 0.0498 | 6.589 |
|  | Female | 7.35 \| 5,195 | 7.66 \| 5,198 | 0.984 (0.856, 1.133) | 0.8270 | 2.134 |
|  | Male | 7.60 \| 6,474 | 6.75 \| 6,477 | 1.164 (1.023, 1.324) | 0.0212 | 1.894 |
|  | Black or African American | 9.37 \| 3,211 | 9.04 \| 3,220 | 1.035 (0.88, 1.215) | 0.6800 | 6.471 |
|  | Hispanic or Latino | 11.29 \| 1,249 | 13.05 \| 1,249 | 0.872 (0.696, 1.093) | 0.2352 | 0.046 |
|  | White | 5.92 \| 5,506 | 6.62 \| 5,498 | 0.918 (0.791, 1.067) | 0.2648 | 1.897 |

**Supplement Table 6**. Risks of other type 2 inflammatory diseases (T2IDs), autoimmune disease, and cardiovascular diseases contrasted amongst patients with transient versus no childhood atopic dermatitis (AD). To counter the bias introduced by multiple testing, a Bonferroni correction was applied, adjusting the significance threshold based on the number of outcomes tested within each disease category: type-2 inflammatory diseases (T2IDs, n=6; adjusted α=0.0083), autoimmune diseases (n=1; adjusted α=0.05), and cardiovascular outcomes (n=3; adjusted α=0.0167). *Abbreviations:* ***VTE****: Venous thromboembolism,* ***MACE****: Major adverse cardiac events.*
